# Genome-wide association study of Alzheimer’s disease brain imaging biomarkers and neuropsychological phenotypes in the EMIF-AD Multimodal Biomarker Discovery dataset

**DOI:** 10.1101/2021.12.22.21268168

**Authors:** Jan Homann, Tim Osburg, Olena Ohlei, Valerija Dobricic, Laura Deecke, Isabelle Bos, Rik Vandenberghe, Silvy Gabel, Philip Scheltens, Charlotte E. Teunissen, Sebastiaan Engelborghs, Giovanni Frisoni, Olivier Blin, Jill C. Richardson, Regis Bordet, Alberto Lleó, Daniel Alcolea, Julius Popp, Christopher Clark, Gwendoline Peyratout, Pablo Martinez-Lage, Mikel Tainta, Richard J. B. Dobson, Cristina Legido-Quigley, Kristel Sleegers, Christine Van Broeckhoven, Michael Wittig, Andre Franke, Christina M. Lill, Kaj Blennow, Henrik Zetterberg, Simon Lovestone, Johannes Streffer, Mara ten Kate, Stephanie J. B. Vos, Frederik Barkhof, Pieter Jelle Visser, Lars Bertram

## Abstract

Alzheimer’s disease (AD) is the most frequent neurodegenerative disease with an increasing prevalence in industrialized, ageing populations. AD susceptibility has an established genetic basis which has been the focus of a large number of genome-wide association studies (GWAS) published over the last decade. Most of these GWAS used dichotomized clinical diagnostic status, i.e. case vs. control classification, as outcome phenotypes, without the use of biomarkers. An alternative and potentially more powerful study design is afforded by using quantitative AD-related phenotypes as GWAS outcome traits, an analysis paradigm that we followed in this work. Specifically, we utilized genotype and phenotype data from n=931 individuals collected under the auspices of the European Medical Information Framework for Alzheimer’s Disease Multimodal Biomarker Discovery (EMIF-AD MBD) study to perform a total of 19 separate GWAS analyses. As outcomes we used five magnetic resonance imaging (MRI) traits and seven cognitive performance traits. For the latter, longitudinal data from at least two timepoints were available in addition to cross-sectional assessments at baseline. Our GWAS analyses revealed several genome-wide significant associations for the neuropsychological performance measures, in particular those assayed longitudinally. Among the most noteworthy signals were associations in or near *EHBP1* (EH domain binding protein 1; on chromosome 2p15) and *CEP112* (centrosomal protein 112; 17q24.1) with delayed recall in a memory performance test. On the X chromosome, which is often excluded in other GWAS, we identified a genome-wide significant signal near *IL1RAPL1* (interleukin 1 receptor accessory protein like 1; Xp21.3). While polygenic score (PGS) analyses showed the expected strong associations with SNPs highlighted in relevant previous GWAS on hippocampal volume and cognitive function, they did not show noteworthy associations with recent AD risk GWAS findings. In summary, our study highlights the power of using quantitative endophenotypes as outcome traits in AD-related GWAS analyses and nominates several new loci not previously implicated in cognitive decline.

## 1 Introduction

Alzheimer’s disease is the most common neurodegenerative disease in humans and the most common form of dementia. In 2018, estimates were published that 50 million dementia patients exist worldwide, about two-third of whom were diagnosed with AD (Patterson, 2018). Pathologically, AD is characterized by the accumulation of extracellular amyloid β (Aβ) peptide deposits (“plaques”) and intracellular hyperphosphorylated tau protein aggregates (“tangles”) in the brain, leading to synaptic dysfunction, neuroinflammation, neuronal loss, and, ultimately, onset of cognitive decline (Sperling et al, 2014; Mattsson et al., 2015). Genetically, AD is a heterogeneous disorder with both monogenic and polygenic forms. The former is caused by highly penetrant but rare mutations in three genes encoding the amyloid beta precursor protein (*APP*) and presenilins 1 and 2 (*PSEN1*/*PSEN2*), which only make up a small fraction (<<5%) of all AD cases (Cacace et al., 2016). Most patients, however, suffer from “polygenic AD”, which is determined by the action (and interaction) of numerous independent genomic variants, likely in concert with nongenetic factors, such as environmental exposures (e.g., head trauma) and lifestyle choices (e.g., alcohol consumption and cigarette smoking) (Bertram and Tanzi, 2020). Based on results from the currently most recent and largest genome-wide association study (GWAS) performed in AD, there are now 38 independent loci showing genome-wide significant association with disease risk (Wightman et al., 2021). The most strongly and most consistently associated AD risk gene is *APOE*, which encodes apolipoprotein E, a cholesterol transport protein that has been implicated in numerous amyloid-specific pathways, including amyloid trafficking, as well as plaque clearance (Holtzman et al., 2012). Although the heritability of polygenic AD is estimated to be around 60-80% (Gatz et al., 2016), *APOE* and the other currently known 37 independent risk loci explain only part of the disease’s phenotypic variance (Wightman et al., 2021). While most AD GWAS only consider clinically diagnosed “probable AD” cases and cognitively unimpaired controls, involving a risk for mis-diagnosis of patients and inclusion of preclinical AD cases as controls, additional information about the genetic architecture of AD and additional statistical power is also afforded by using “endophenotypes” related to AD, ideally measured on a quantitative scale such as biomarker data, imaging, or neurocognitive performance (Gottesman and Gould, 2003; MacRae and Vasan, 2011; Zhang et al., 2020).

In our study, we expand earlier work from our group (Hong et al., 2020; Hong et al., 2021) derived from European Medical Information Framework Alzheimer’s Disease Multimodal Biomarker Discovery (EMIF-AD MBD) sample (Bos et al., 2018). Specifically, in two previous GWAS we set out to identify variants underlying variation in several cerebrospinal fluid (CSF) phenotypes, such as levels of CSF Aβ and tau protein (Hong et al., 2020), or neurofilament light (NfL) chain, chitinase-3-like protein 1 (YKL-40), and neurogranin (Ng), which reflect axonal damage, astroglial activation, and synaptic degeneration, respectively (Hong et al., 2021). However, the EMIF-AD MBD dataset features several other quantitative phenotypes, including cross-sectional MRI measurements and cross sectional and longitudinal neuropsychological tests, which are used as outcome traits in the current study. Specifically, we performed GWAS and polygenic score (PGS) analyses on seven neuropsychological (using both cross-sectional and longitudinal data) and five brain imaging phenotypes (using cross-sectional data from MRI scans). In the 19 performed GWAS scans (which also included the X chromosome), we identified a total of 13 genome-wide significant loci highlighting several novel genes showing association with the analyzed traits. While we do not see a noteworthy overlap in the genetic architectures underlying our “endophenotypes” and AD by polygenic score (PGS) analysis, we did observe significant correlations in PGS constructed from earlier GWAS on hippocampal volume (Hibar et al., 2017) and general cognitive function (Davies et al., 2018) with the respective phenotypes in EMIF-AD MBD. Taken together, our novel results pinpoint several new genetic loci potentially involved in AD-related pathophysiology.

## 2 Materials and Methods

### 2.1 Sample description

Analyses were based on the EMIF-AD MBD dataset which was collected across eleven different European study centers (Bos et al., 2018). In total, this dataset included 1221 (563 [46%] female; mean age = 67.9 years, SD=8.3) individuals from three diagnostic stages: normal controls (NC), subjects with mild cognitive impairment (MCI) and subjects with a clinical diagnosis of AD. An overview of the quantitative phenotypes investigated in this study is provided in Table 1. Due to partially missing phenotype data (in the neurocognitive domain), the effective sample sizes vary for the different GWAS analyses (see Table 1). The local medical ethical review boards in each participating recruitment center had approved the study prior to commencement. Furthermore, all subjects had provided written informed consent at the time of inclusion in the cohort for use of data, samples and scans (Bos et al., 2018).

**Table 1:** Description of EMID-AD MBD datasets analyzed per phenotype. “MAF filter” denotes the applied MAF filter for each GWAS. For cross-sectional MMSE we used an MAF threshold of 0.02 due to residual inflation of the GWAS test statistics. Information on tests used for generating baseline and longitudinal phenotypes can be found in the Supplementary Material. MMSE = Mini Mental State Examination. “n.a.” = not available.

| Category | Phenotype | Baseline |  | Longitudinal |  |
| --- | --- | --- | --- | --- | --- |
|  |  | Sample size | MAF filter | Sample size | MAF filter |
| Neuropsychological | MMSE | 867 | 0.02 | 520 | 0.02 |
|  | Attention | 806 | 0.01 | 402 | 0.02 |
|  | Executive functioning | 686 | 0.01 | 234 | 0.02 |
|  | Language | 849 | 0.01 | 409 | 0.02 |
|  | Memory Delayed | 729 | 0.01 | 337 | 0.02 |
|  | Memory Immediate | 797 | 0.01 | 345 | 0.02 |
|  | Visuoconstruction | 429 | 0.02 | 149 | 0.04 |
| MRI | Fazekas score | 606 | 0.01 | n.a. | n.a. |
|  | Cortical thickness | 560 | 0.01 | n.a. | n.a. |
|  | Left Hippocampus volume | 605 | 0.01 | n.a. | n.a. |
|  | Right Hippocampus volume | 605 | 0.01 | n.a. | n.a. |
|  | Summed Hippocampus volume | 605 | 0.01 | n.a. | n.a. |

### 2.2 MRI phenotypes description

The five MRI phenotypes were collected for 862 subjects. Brain MRIs were used to assess hippocampal volume (mm^3^, left and right hemisphere, and sum of both; all adjusted for intracranial volume), whole brain cortical thickness (in mm), and white matter lesions (WML; using the Fazekas scale) (Ten Kate et al., 2018). The Fazekas scale categorizes WMLs into 4 categories: Level 0 (no or almost no lesion), level 1 (multiple punctate lesions), level 2 (early confluent WML), and level 3 (presence of large confluent WML). Details on the scanning procedures and data harmonization across centers can be found in Bos et al. (2018) and Ten Kate et al. (2018).

### 2.3 Neuropsychological phenotypes description

Cross-sectional (and follow-up) data were available for the following seven neuropsychological domains within the EMIF-AD MBD dataset: global cognition (Mini Mental State Examination, MMSE), attention, executive function, language, memory (immediate and delayed) and visuoconstruction. For each cognitive domain, a primary test was selected by Bos et al. (2018). If the preferred test were not available, an alternative priority test from the same cognitive domain was chosen. More details on the neuropsychological tests used for generating these phenotypes can be found in Bos et al. (2018). Raw data on these tests were normalized with the help of a *z*-transformation, so that the data were comparable within a cognitive domain despite representing partially different tests across centers. For the cross-sectional GWAS analyses, the *z*-scores derived from baseline data were used. The number of subjects used for each test can be found in the Supplementary Material. For all seven neuropsychological domains, follow-up data from at least one additional time point were available for each individual and used to construct a longitudinal phenotype using the following formula (which estimates the relative change in cognitive performance per time interval [here: years]):

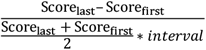

When calculating longitudinal phenotypes, this formula was applied separately for each neuropsychological test. Outlying scores were determined using false discovery rate (FDR) <0.05 estimations and were excluded from all subsequent analyses. Only the most frequently used tests per cognitive domain were included in the final phenotypes. For more information, see Supplementary Material. Both baseline and longitudinal phenotypes were adjusted for age at baseline.

### 2.4 DNA extraction, genotype imputation and quality control

A detailed description of the genotyping procedures, quality control (QC) and subsequent data processing can be found in Hong et al. (2020) and in the Supplementary Material. Here, the same genotype data were used for the GWAS analyses. Briefly, 936 DNA samples were subjected to genome-wide SNP genotyping using the Infinium Global Screening Array (GSA) with Shared Custom Content (Illumina Inc.). Imputation was then performed using Minimac3 (Das et al., 2016). Extensive post-imputation QC resulted in 7,464,105 autosomal SNPs with a minor allele frequency (MAF) ≥0.01 in 888 individuals of European ancestry. More details can be found in the Supplementary Material.

For the X chromosome, QC was performed separately for male and female subjects for non-pseudoautosomal regions, using slightly different criteria compared to the autosomes (see Supplementary Material). In contrast, pseudoautosomal regions (PAR1 and PAR2) were treated analogously to the autosomal SNPs. After QC, imputations were performed on the Sanger Institute imputation server (https://imputation.sanger.ac.uk/) using the extended HRC reference panel (McCarthy et al., 2016). After imputation, we used the same QC criteria as for the autosomal SNPs but performed these separately for female and male data sets, except the HWE test (*P*<1.0E-4) which was performed on all samples combined as recommended previously (Graffelmann and Weir, 2016) and implemented in PLINK2. For males, markers were coded as 0 vs. 2 (instead of 0 vs. 1), to adjust for the missing second X chromosome (as recommended in Smith et al. [2021]).

### 2.5 GWAS and post-GWAS analyses

SNP-based association analyses were performed assuming an additive linear model (command: --glm) using allele dosages (to account for imputation uncertainty) in PLINK2 (Purcell et al., 2007). The covariates included in the analyses were sex, diagnostic status and the first three principal components from a principal component analysis (PCA) to adjust for population-specific differences. Generally, we excluded SNPs from the GWAS analyses with MAF<0.01. However, due to differences in the effective sample sizes across phenotypes this threshold was adapted upward (up to 0.04) to prevent inflation of test statistics owing to low frequency SNPs (see Table 1 for more details). Diagnostic status was coded with two dummy variables as follows: NC = (0,0), MCI = (0,1), AD = (1,1). For four longitudinal cognitive phenotypes an additional dummy variable was introduced to code for the neuropsychological test used, in cases where two different tests were used for generating these phenotypes. Details can be found in the Supplementary Material.

To explore associations on the X chromosome that were potentially driven by genetic sex, we additionally conducted the analyses separately in females and males. We then combined these two additional sets of results in a meta-analysis using Stouffer’s method as implemented in METAL (Willer et al., 2010). As we found no noteworthy differences in the results using Stouffer’s method, only the results from the linear regression analysis in the combined sample are shown.

The FUMA platform (http://fuma.ctglab.nl/; Watanabe et al., 2017) was used for post-GWAS analyses, including gene-based association analyses (via MAGMA (de Leeuw et al., 2015)) and to annotate and visualize the GWAS results. To this end, we defined genome-wide significance at α<5.0E-08 for the SNP-based analyses while genome-wide suggestive evidence was set at α<1.0E-05. For the gene-based analyses, we adjusted for the number of protein-coding genes examined (19,485) using the Bonferroni method, resulting in a threshold of α<2.566E-06.

In FUMA, both the SNP annotation and the Combined Annotation Dependent Depletion (CADD) score (Rentzsch et al., 2021) are provided. The main GWAS results are reported only for “independent significant” SNPs, as defined by FUMA. These represent SNPs that are not highly correlated with one another using a threshold of *r*^*2*^<0.6 (using reference data from the 1000 Genomes Project).

Subsequently, the top SNPs, i.e., those with the smallest *P* values per respective phenotype, were examined in more detail using additional tools. First, the Variant Effect Predictor on Ensembl (VEP, http://grch37.ensembl.org/Tools/VEP; McLaren et al., 2016) was used to determine a possibly functional effect due to changes in the coding sequence, e.g. missense variants. Second, SNPs were examined using data from the RegulomeDB database (https://regulomedb.org/regulome-search; Boyle et al., 2012) to assess possible effects on gene expression. Third, we used data from the Genotype-Tissue Expression (GTEx, V8) project portal (https://www.gtexportal.org/home/; Lonsdale et al., 2013) to assess whether SNPs represent expression / splicing quantitative trait loci (eQTLs/sQTLs). While GTEx provides data on gene expression in 54 tissues, we laid particular emphasis in genes expressed in brain. Lastly, we interrogated the GWAS catalogue (https://www.ebi.ac.uk/gwas/home; Buniello et al., 2019) to assess whether any of the top SNPs were previously reported to show association with other phenotypes by GWAS. To this end, we considered genes and loci within a 1 Mb region (± 500,000 bp) around the SNP of interest. In case SNPs not identical to our “top SNP” were reported to show association with an AD-relevant phenotype (brain imaging, cognition, etc.), the LDlink platform (Machiela and Chanock, 2015) was used to determine pairwise LD to top SNPs (https://ldlink.nci.nih.gov/?tab=ldpair). In this context we defined relevant LD using a threshold of *r*^*2*^>0.6.

### 2.6 Polygenic score (PGS) analysis

In addition to the primary GWAS analyses described above, we also calculated polygenic scores (PGS) to estimate the extent of genetic correlation with the GWAS results for three other phenotypes. To this end, we used the summary statistics of a GWAS on AD risk (Jansen et al., 2019) as comparison to both phenotypic domains (MRI and neurocognitive performance) of our study, and the GWAS on general cognitive function (Davies et al., 2018) as comparison to the GWAS on neuropsychological phenotypes. Finally, the GWAS on hippocampal volume (Hibar et al., 2018) served as comparison to our GWAS analyses on MRI phenotypes. PGS calculations were performed using PRSice-2 software (Choi and O’Riley, 2019). Statistical analyses fitted general linear regression models with PGS as predictor adjusting for the same covariates as in the primary GWAS analyses: sex, diagnostic status, and PC1-3 (and type of cognitive test, where applicable). To adjust for multiple testing of this arm of our study, we used a conservative threshold based on Bonferroni adjustment (α<5.0E-03 = 0.05 / 5*2 for the MRI phenotypes, and α<1.8E-03 = 0.05 / 14*2 for the neuropsychological phenotypes). However, given the (at least partial) correlation between phenotypes, we note that the true threshold is likely somewhere between 0.05 and these Bonferroni-adjusted values.

## 3 Results

### 3.1 GWAS on MRI phenotypes

The genomic inflation factor λ ranged between 1.004 and 1.012 in all five SNP-based analyses, indicating that the results of the MRI GWAS analyses were not affected by substantial inflation of the test statistics. In the actual association analyses of the five quantitative MRI phenotypes, we identified no genome-wide significant (*P*<5.0E-08) signals but observed 385 variants with at least suggestively significant (*P*<1.0E-05) evidence of association (Supplementary Tables 15-19). The lowest *P* value was observed with SNP rs16829761 for the Fazekas phenotype (*P*=5.08E-08; Supplementary Figure 31), which only fell slightly above the genome-wide significance threshold. According to VEP (McLaren et al., 2016), this variant is located in an intron of the genes *IQCJ* (protein: IQ motif containing J) and *SCHIP1* (protein: schwannomin-interacting protein 1). In the GTEx database (Lonsdale et al., 2013), the lead-SNP identified here (rs16829761) is not listed as eQTL or sQTL, which may be due to the comparatively low MAF (0.01). The CADD score, i.e., the *in silico* predicted deleteriousness, of rs16829761 is also low at approximately 0.074. In addition, none of the gene-based GWAS analyses using MAGMA revealed any genome-wide significant signals (*P*<2.566E-06) using the MRI traits analyzed. The genomic inflation factor λ ranged between 0.984 and 1.060 in these five gene-based analyses.

### 3.2 GWAS on neuropsychological phenotypes

Across the 14 GWAS performed on cross-sectional and longitudinal neuropsychological phenotypes available in EMIF-AD MBD, there were a total of 13 genome-wide significant loci, two of which were identified via the gene-based analyses using MAGMA (de Leeuw et al., 2015). Three of the genome-wide significant signals were observed in the analyses of cross-sectional phenotype data and ten with longitudinal outcomes. Overall, none of the sets of GWAS results in this arm of our study appeared to be strongly affected by inflation of the genome-wide test statistics as evidenced by genomic inflation factors near 1 (range: 0.969-1.012 in the SNP-based analyses and 0.922-1.036 in the gene-based analyses). Table 2 provides a detailed summary of these genome-wide significant loci, and Figure 1 shows multi-trait Manhattan (MH) plots of the SNP-based GWAS results for cross-sectional (Figure 1A) and longitudinal (Figure 1B) analyses (for corresponding QQ plots: see Supplementary Figures 1 and 2). The following two paragraphs highlight the most interesting results in either the analyses of cross-sectional or longitudinal neuropsychological traits.

**Table 2:**
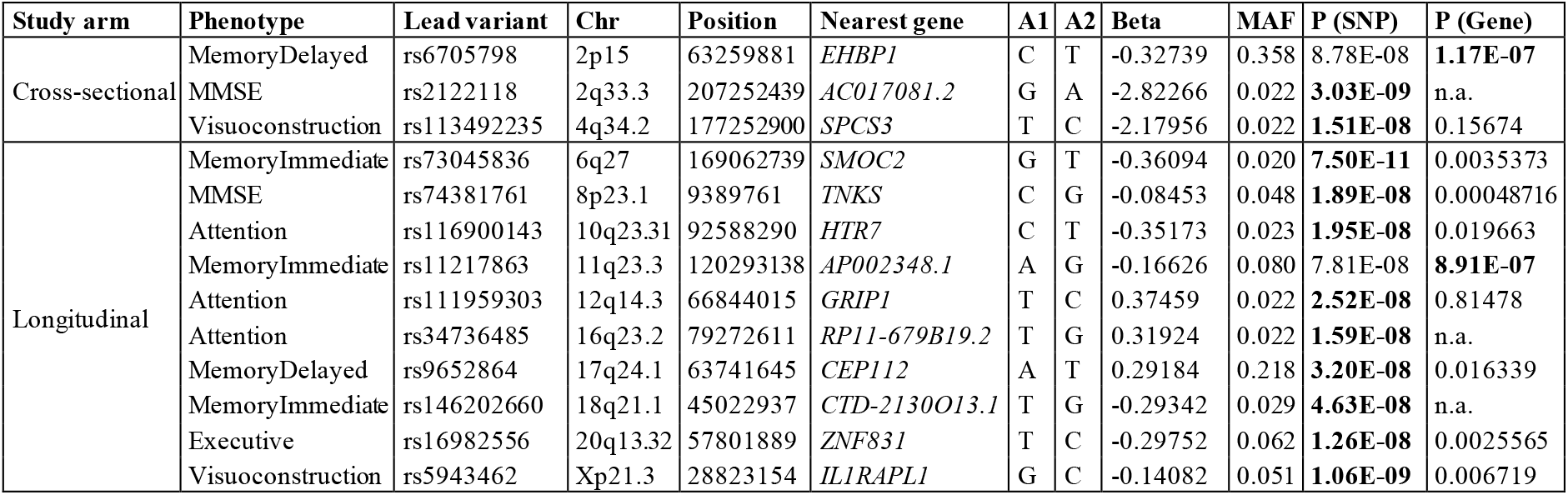
Genome-wide significant associations observed in GWAS of cognitive phenotypes. **Bold font** indicates genome-wide significant (on SNP- or gene-level) results (see Methods section for details). “Chr” and “Position” according to GRCh37/hg19. “A1” denotes the effect allele. “P (SNP)” is the *P* value of the lead SNP at this locus. “P (Gene)” is the *P* value belonging to “Nearest gene”. Top results from these GWAS analyses can be found in the Supplementary Tables. MMSE = Mini Mental State Examination. “n.a.” = not available.

**Figure 1:**
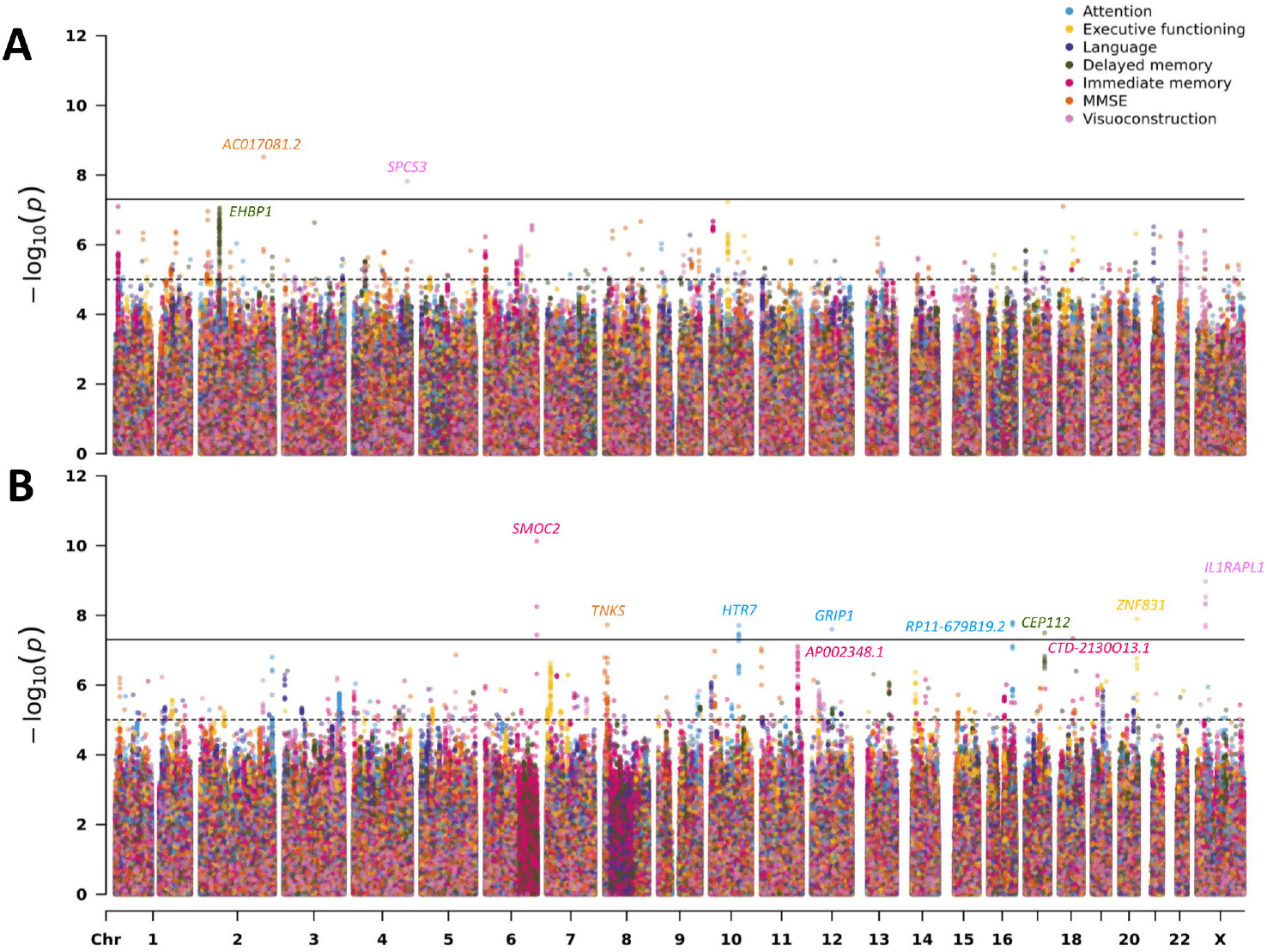
Multi-trait Manhattan plots for the SNP-based GWAS results on neuropsychological phenotypes (A: cross-sectional; B: longitudinal). For details on the analyzed traits see Methods and Supplementary Material.

#### Analyses of cross-sectional data

The most interesting finding in this domain was elicited by markers in *EHBP1* which showed genome-wide significant evidence of association with the delayed recall memory phenotype in the gene-based analysis (*P*=1.17E-07; Table 2; Supplementary Figure 20). The lead SNP (rs6705798) in this region only missed the genome-wide significance threshold by a small margin (*P*=8.78E-08; Table 2; Figure 1A). *EHBP1* is located on chromosome 2p15 and encodes EH domain binding protein 1.

#### Analyses of longitudinal data

The strongest signal in the longitudinal analyses was elicited by a locus on chromosome 6q27 (rs73045836; *P*=7.50E-11; Table 2; Figure 1B; Supplementary Figure 25) in the analysis using an immediate memory recall paradigm. This SNP is located in an intron of *SMOC2* coding for secreted modular calcium-binding protein 2, which, among other functions, promotes extracellular matrix assembly (Gao et al., 2019). It needs to be noted that with an MAF ∼2% this SNP is rather infrequent which may increase the possibility of representing a false-positive finding. Perhaps more interesting is the association signal observed near SNP rs5943462 (MAF ∼0.05) and the visuoconstruction phenotype on the X chromosome (*P*=1.06E-09; Table 2; Figure 1B; Supplementary Figure 29). This SNP is an intronic variant located in *IL1RAPL1* encoding interleukin 1 receptor accessory protein-like 1, which belongs to a class of molecules that regulate synapse formation (Montani et al., 2019). The third highlighted signal in this domain relates to the genome-wide significant variant rs74381761 (MAF ∼0.05) on chromosome 8p23.1 (*P*=1.89E-08; Table 2; Figure 1B; Supplementary Figure 5) which shows association with the longitudinal MMSE phenotype. The lead SNP is located in an intergenic region near *TNKS* (gene-based *P*=4.87E-04; Table 2). This gene encodes the protein tankyrase, which belongs to a class of poly (ADP-ribose) polymerases and is involved in various processes in the body, such as telomere length regulation, the Wnt/β-catenin signaling pathway, or glucose transport (Damale et al., 2020). The last featured signal relates to the association observed near SNP rs9652864 (MAF ∼0.22) on chromosome 17q24.1 (*P*=3.20E-08; Table 2; Figure 1B; Supplementary Figure 21) and the delayed recall test. This variant is located in an intron of *CEP112*, which encodes centrosomal protein 112. Overall, there were eight correlated SNPs in this locus all showing strongly association (Supplementary Table 10).

#### Comparison of cross-sectional vs. longitudinal GWAS results

After completion of the separate GWAS on cross-sectional and longitudinal outcomes, we assessed whether the results of these two analysis arms showed any overlap. To this end, we followed two approaches: First, we performed a look-up of top results from one paradigm in the equivalent other. Specifically, we checked whether a genome-wide significant SNP from the cross-sectional analyses also had a low *P* value in the corresponding longitudinal GWAS and vice versa.The lowest corresponding *P* value was 0.015 (at baseline) for rs73045828, which attained *P*=5.65E-09 in the longitudinal GWAS for immediate memory (Supplementary Table 21). No further signal overlaps were observed across corresponding cross-sectional and longitudinal phenotypes. Second, we took a more comprehensive approach by comparing a larger set of SNPs across both phenotypic domains. To this end, we constructed PGS from the summary statistics of the cross-sectional GWAS (as an approximate measure of “aggregated SNP effects”) and used these PGS as independent variables in a linear model predicting longitudinal outcomes. Effectively, this allowed us to determine how much phenotypic variance in the longitudinal data can be explained by top SNPs of the matching cross-sectional GWAS. Overall, these analyses did not reveal a substantial correlation in genetic results for corresponding phenotypes (Supplementary Table 22), in agreement with the look up of individual SNPs (see above). The best model fit was observed with the PGS for executive function and visuoconstruction, where the GWAS top SNPs from the cross-sectional data used in the PGS explained 4-9% of the phenotypic variance of the corresponding longitudinal outcomes, respectively (Supplementary Table 22). We note, however, that the PGS method was not designed for computing genetic correlations of non-independent samples (as is the case here), so this analysis must be considered “exploratory “, and the reported results represent no more than “upper bounds” of the potential genetic correlations.

### 3.3 Role *APOE* in GWAS on MRI and neuropsychological performance

Given the substantial role that variants in *APOE* play in the genetic architecture of AD, we present findings for this locus separately, i.e. the results for SNP rs429358 (which defines the ε4 allele) and rs7412 (which defines the ε2 allele). In relation to the common genotype ε3/ε3, the risk to develop AD is increased by a factor of ∼3.2 for genotype ε3/ε4, while two ε4 alleles (genotype ε4/ε4) show ORs around 10-12 when compared to normal controls (Neu et al., 2017). The minor allele at rs429358 (ε4) is overrepresented in the EMIF-AD MBD dataset with an MAF ∼29% (the MAF in the general Northern European population is ∼16%), which is due to the special design of participant recruitment (see Bos et al. (2018)). For the neuropsychological phenotypes, the *P* values of rs429358 are unremarkable except for the domain “delayed memory”, where *P* values of 0.0005 and 0.0042 were observed for the baseline and longitudinal analyses, respectively (Supplementary Table 20). In the MRI analyses, the only association signal observed with rs429358 was with hippocampal volume (Supplementary Table 20). Interestingly, this was driven by an association with the volume of the left (*P*=0.0002) hippocampus, while no association was observed with the corresponding data of the right hemisphere (*P*=0.2956). We note that for both traits, i.e. delayed memory and left hippocampal volume, the effect direction the corresponding *β* coefficient is consistent with the deleterious effect of the minor (T/ε4) allele at rs429358 known from the literature (Neu et al., 2017). For the minor allele at rs7412 (ε2) we observed no noteworthy association signals in any of the analyses performed in this study (Supplementary Table 20), although power for this variant was much reduced owing to its lower MAF (4.6% here, 7.5% in the general western European control population).

### 3.4 Polygenic score (PGS) analyses using published GWAS results

In these analyses we aimed to estimate the degree of genetic overlap between the MRI and neuropsychological outcomes available in EMIF-AD MBD and other relevant traits from the literature, such as AD risk, using published GWAS summary statistics.

#### PGS analyses with MRI phenotypes

As expected, the strongest overlap was observed with a prior GWAS also using MRI outcomes. Specifically, we used GWAS results by the ENIGMA group (Hibar et al., 2017) who studied 26 imaging traits in n=33,536 individuals. Here, the best overlap was seen with each of the three hippocampal MRI traits (up to 2.7% variance explained, *P*=6.0E-06; Table 3; Supplementary Table 24). In contrast, in PGS analyses using SNPs associated with AD risk (Jansen et al., 2019), we found only one moderate correlation with white matter damage (measured by the Fazekas score). For this trait the AD SNPs explained 1.4% variance (*P*=3.7E-03; Table 3; Supplementary Table 24).

**Table 3:** Summary of PGS results significant after multiple testing correction. “Threshold” refers to *P* value cut-off used for PGS construction in prior GWAS summary statistics and “Number of SNPs” refers to the LD-pruned SNPs passing this threshold that are included in PGS calculations. “R2” denotes the phenotypic variance explained by the SNPs of the prior GWAS in the EMIF-AD MBD dataset. A full listing of results from these PRS analyses can be found in Supplementary Material. MMSE = Mini Mental State Examination.

| Prior GWAS | Phenotype | Threshold | Number of SNPs | R2 | P value |
| --- | --- | --- | --- | --- | --- |
| Hibar et al. (2017) | Hippocampus volume sum | 0.0001 | 127 | 0.027 | 6.06E-06 |
|  | Hippocampus volume left | 0.0001 | 127 | 0.026 | 9.98E-06 |
|  | Hippocampus volume right | 0.0001 | 127 | 0.024 | 2.48E-05 |
| Jansen et al. (2019) | Fazekas | 0.17075 | 22269 | 0.014 | 3.72E-03 |
| Davies et al. (2018) | Baseline MMSE | 0.248 | 63792 | 0.016 | 1.66E-06 |
|  | Baseline executive functioning | 0.0014 | 4469 | 0.018 | 1.98E-05 |
|  | Baseline language | 0.0061 | 9031 | 0.010 | 1.25E-03 |
|  | Longitudinal attention | 5.0E-08 | 163 | 0.023 | 1.79E-03 |

#### PGS analyses with neuropsychological phenotypes

As for the MRI data, the best fit in the PGS analyses with the neuropsychological phenotypes was observed with a GWAS that also used neurocognitive performance as outcome (Davies et al., 2018). Specifically, this study defined a PCA-derived factor for “general cognitive function” which was analyzed in >300,000 individuals. In EMIF-AD MBD, associations with four of the 14 calculated PGS fell below the multiple testing threshold of 1.8E-03 (Table 3). The strongest association was observed with the longitudinal attention function for which the GWAS results from Davies et al. (2018) explained 2.3% of the phenotypic variance (*P*=1.79E-03, Table 3; Supplementary Table 23). The next best associations were seen with longitudinal executive functioning (*r*^*2*^=0.028; *P*=9.79E-03; Supplementary Table 23) and visuoconstructional abilities (*r*^*2*^=0.058; *P*=3.08E-03; Supplementary Table 23). However, these latter two associations do not survive multiple testing correction (Table 3). Interestingly and similar to the MRI-based results, we did not find strong evidence for a genetic overlap between the neurocognitive outcomes tested here and AD risk based on Jansen et al. (2019) (Supplementary Table 23). This included the various phenotypes measuring components of “memory” performance, regardless of whether or not they were ascertained cross-sectionally or longitudinally.

## 4 Discussion

This study extends previous GWAS analyses from our group utilizing phenotypic data from the EMIF-AD MBD study (Hong et al., 2020; Hong et al., 2021). The overarching goal of this work was to decipher the genetic architecture of AD-related MRI and neuropsychological (endo)phenotypes to better understand AD pathophysiology. Both previous EMIF-AD MBD GWAS focused on AD biomarkers measured in CSF and, among other findings, identified variants in *TMEM106B* as *trans*-pQTLs of CSF neurofilament light (NfL) levels (Hong et al., 2021). Interestingly, the same locus was subsequently highlighted as a novel AD risk locus in a GWAS on >1.1 million individuals (Wightman et al. 2021), showcasing the power of the quantitative biomarker GWAS approach that was also followed in this study. In the current work, we focused on biomarkers / phenotypes derived from brain imaging and neuropsychological testing in the same EMIF-AD MBD individuals. Overall, we performed 19 individual GWAS and identified a total of 13 genome-wide significant loci highlighting several novel genes that are potentially involved in contributing to AD pathophysiology. Our study represents one of few GWAS in the literature to also include the X chromosome, where we identified a genome-wide significant association between markers near *IL1RAPL1* and longitudinal visuoconstructive ability. Interestingly, neither *APOE* nor the other recently described AD GWAS loci appear to have a major impact on the traits analyzed in our study. In summary, our extensive genome-wide analyses nominate several novel loci potentially involved in neurocognitive functioning. Some of these may prove informative to better understand the genetic forces underlying AD and related phenotypes.

In the remainder of this section, we discuss the potential role of five loci, which we consider the most interesting findings of our study. The strongest GWAS signal was elicited by SNP rs73045836 (*P*=7.50E-11; Table 2; Figure 1B; Supplementary Figure 25) showing genome-wide significant association with the longitudinal data of the immediate recall memory phenotype. The gene annotated to the associated region on chromosome 6q27, *SMOC2*, encodes secreted modular calcium-binding protein 2. SMOC2 is an extracellular matrix protein from the secreted protein, acidic and rich in cysteine (SPARC) family (Gao et al., 2019) recently linked to age-dependent bone loss in humans (Morkmued et al., 2020). Despite performing careful literature and database searches, we could not pinpoint any obvious mechanistic connection of this locus to cognitive functioning or other AD-relevant phenotypes.

The second strongest association signal was observed near SNP rs5943462 (MAF ∼0.05) on the X chromosome (*P*=1.06E-09; Table 2; Figure 1B; Supplementary Figure 29) with the longitudinal data of the visuoconstruction phenotype. The SNP is located in an intron of *IL1RAPL1*. This gene encodes interleukin 1 receptor accessory protein-like 1, which belongs to a class of molecules that regulate synapse formation. *IL1RAPL1* is mostly expressed in brain areas that are involved in memory development, such as hippocampus, dentate gyrus, and entorhinal cortex, suggesting that the protein may have a specialized role in physiological processes underlying memory and learning abilities (Montani et al., 2019). Even small changes in the expression and function of these proteins can provoke major alterations in synaptic connectivity, resulting in cognitive damage (Montani et al., 2019). Moreover, *IL1RAPL1* was nominated as a candidate gene for X-linked mental retardation (Raymond, 2006). Although the GWAS on longitudinal visuoconstruction included only 149 individuals, we believe this signal to be plausible and very interesting because of the well-established role of *IL1RAPL1* on human brain function.

The third highlighted signal relates to the association between variant rs74381761 (MAF ∼0.05) on chromosome 8p23.1 (*P*=1.89E-08; Table 2; Figure 1B; Supplementary Figure 5) and longitudinal MMSE measurements. This SNP is located in an intergenic region near *TNKS*. This gene encodes the protein tankyrase, which belongs to a class of poly (ADP-ribose) polymerases and is involved in various processes in the body, such as telomere regulation, Wnt/β-catenin signaling pathway or glucose transport (Damale et al., 2020). According to GTEx (Lonsdale et al., 2013), *TNKS* is highly expressed in brain (mostly in cerebellum). Moreover, SNPs annotated to *TNKS* were associated with brain white matter hyperintensity (WMH) measurements (Armstrong et al., 2020; Sargurupremraj et al., 2020; Zhao et al., 2021) and cortical surface area measurements (Grasby et al., 2020) according to the GWAS catalog (Buniello et al., 2019). With a gene-based *P* value of 4.87E-04 and the strong functional link to brain function, we consider the signal around *TNKS* as plausible and very interesting.

The last highlighted finding from the longitudinal analyses relates to the genome-wide significant association observed between SNP rs9652864 and the delayed recall memory phenotype (*P*=3.20E-08; Table 2; Figure 1B; Supplementary Figure 21). This variant (MAF=0.218) attained a *P* value of 6.73E-04 in the GWAS of Davies (2018) on cross-sectional cognitive performance, lending additional support to our finding. The SNPs is located in an intron of *CEP112* which encodes centrosomal protein 112. Centrosomal proteins are known as the components of the centrosome involved in centriole biogenesis, cell cycle progression, and spindle-kinetochore assembly control (Mazaheri Moghaddam et al., 2021). Despite showing only low levels of expression in the central nervous system (CNS) according to GTEx, SNPs in this gene have been associated with cortical surface area by neuroimaging in two independent GWAS (Grasby et al., 2020; van der Meer et al., 2020) according to the GWAS catalog (Buniello et al., 2019). However, none of these neuroimaging SNPs is in relevant LD (*r*^*2*^>0.6) to the lead variant identified here. Notwithstanding, given that variants in this gene have shown genetic links to both cognitive function and structural brain imaging, we consider this finding as plausible and highly interesting.

In the GWAS analyses of the cross-sectional neurocognitive phenotypes, we observed three genome-wide significant signals, of which we consider the gene-based association with *EHBP1* as the most interesting finding (*P*=1.17E-07; Table 2; Figure 1A; Supplementary Figure 20). This protein interacts with Eps15-homology domain-containing protein 1/2 (EHD1/2) that plays a central role in GLUT4 transport and couples endocytic vesicles to the actin cytoskeleton (Rai et al., 2020). It is highly expressed in many tissues, including the brain, according to GTEx (Lonsdale et al., 2013). While there does not appear to be an obvious link between *EHBP1* and brain function in the literature (e.g. in the GWAS catalog [Buniello et al., 2019]), we note that this gene is located within 5kb of *OTX1* (orthodenticle homeobox 1; gene-based *P*=1.19E-05), which acts as transcription factor and plays a role in brain and sensory organ development in Drosophila and vertebrates, including humans (Omodei et al., 2009). Our lead SNP in this region, i.e. rs6705798, falls just short of attaining genome-wide significance (*P=*8.78E-08; Table 2; Figure 1A) and is reported to represent an eQTL of both *OTX1* and *EHBP1* in various human tissues according to GTEx (Lonsdale et al., 2013).

In addition to searching for novel genetic determinants of the neuroimaging and neurocognitive traits analyzed in this study, we also investigated the overlap with known GWAS findings. First and foremost, this relates to two commonly studied alleles in the *APOE* gene, which appear to only play a minor role in this setting. Specifically, SNP rs429358, which defines the ε4 allele in *APOE*, does not even reach genome-wide suggestive significance (*P<*1.0E-05) in any of the 19 GWAS investigated here. The strongest associations with this allele were seen with MRI-based hippocampus volume (left volume *P*=0.0002, summed volume *P*=0.0005; Supplementary Table 20) and with the delayed recall memory test (baseline *P*=0.0005, longitudinal *P*=0.0042; Supplementary Table 20). The effect directions of these associations are consistent with the deleterious influence of rs429358 on AD (Neu et al., 2017). These at best marginal associations are in line with the literature: In the GWAS on cognitive function by Davies (2018), *APOE* ε4 also only showed marginal association (*P*=2.2E-04), while it was not reported to show any evidence of association with hippocampal volume in the GWAS by Hibar (2017). These findings are different from our earlier GWAS in the EMIF-AD MBD dataset, where the ε4 allele showed very pronounced evidence of association in CSF and imaging markers related to Aβ42 (Hong et al., 2020). Extending the comparison to additional genetic variants associated with AD risk in the GWAS by Jansen et al. (2019) did also not show any noteworthy or consistent overlap with the GWAS results generated in this study. In contrast, highly significant overlaps by PGS analysis were observed upon using GWAS results from Davies et al. (2018) for the neuropsychological and Hibar et al. (2017) for the MRI phenotypes, which is not surprising given that very similar neuropsychological and neuroimaging traits were used as outcomes in these studies. Collectively, the PGS results of this and previous work show that there is only very limited overlap in the genetic architecture (at least when studying common SNPs) between AD on the one and neuropsychological performance or structural brain imaging on the other hand. We note that this does not preclude the possibility that certain molecular pathways targeted by the genes highlighted in this GWAS may be shared with AD pathophysiology.

While our study has several noteworthy strengths (e.g. the use of highly standardized procedures in generating and harmonizing both the genotype and phenotype data of our study, use of both cross-sectional and longitudinal neurocognitive performance data, inclusion of the X chromosome in the GWAS), it may also have been negatively affected by some limitations. First and foremost, we note that the sample size used for the present analyses is comparatively small for “GWAS standards” and was well under 1,000 in some instances (Table 1). Accordingly, the statistical power of these analyses was low. This limitation is at least partially countered by the quantitative nature of nearly all analyzed phenotypes: it is well established that quantitative trait association analyses are more powerful than those using binary phenotypes, e.g. in a case-control setting (Bush and Moore, 2012). Second, in addition to resulting in low power, small sample sizes also increase the possibility of false-positive findings, especially for infrequent variants (i.e. those with an MAF <5%). In this context we note that eight of our thirteen genome-wide significant signals were elicited by such variants. Thus, independent replication – ideally in larger datasets – is needed to confirm the main findings of our GWAS before any further-reaching conclusions can be reached. Third, we note that the phenotype data used as outcome traits in our GWAS analyses were collected at different participating centers at times using different types of examinations (e.g. different tests to study the same overarching neuropsychological domain). To alleviate potential bias resulting from this inherent phenotypic heterogeneity, all clinical data were processed, quality-controlled and harmonized (e.g. by normalizing most variables within centers) centrally by an experienced team of researchers (see Bos et al. (2018) for more details). We emphasize that this potential heterogeneity does not apply to the genetic data as these were generated in one laboratory experiment and subsequently processed jointly in one analytical framework, minimizing the emergence of potential batch effects. Last but not least, we emphasize that owing to its particular ascertainment design (Bos et al., 2018) the EMIF-AD MBD dataset does not (attempt to) constitute a representative sample from the “general population”. Accordingly, the results presented here cannot be generalized to the general population. We note that the same is true for many GWAS in this and other fields, which typically use clinic-based ascertainment which is not representative of the population as a whole.

In conclusion, our study delivers an entirely novel set of GWAS results from participants of the EMIF-AD MBD dataset. We nominate several novel and functionally interesting genetic association signals with phenotypes related to neurocognitive function and structural brain imaging. Even though independent replication is still needed, our results may prove informative to better understand the genetic forces underlying AD and related phenotypes.

## Data Availability

GWAS summary statistics for the top (*P* value <1.0E-05) results are listed in the Supplementary Tables. Full GWAS summary statistics are available from the authors upon request. Clinical data and genome-wide genotyping data are stored on an online data platform using the “tranSMART” data warehouse framework. Access to the genome-wide genotyping data can be requested from the corresponding author of this study who will forward each request to the EMIF-AD data access team. All scripts used to generate the primary GWAS and PGS analyses are available from the authors upon request.

## Author Contributions

JH and TO performed all the analyses and together with LB interpreted the data and wrote the manuscript. OO performed X chromosomal analyses. VD was responsible for EMIF-AD MBD DNA sample preparation and DNA extraction. LD contributed to the interpretation of the data and visualization of the results. IB, SV, and PJV coordinated the collection and harmonization of phenotypes and biosamples in EMIF-AD MBD. MW and AF supervised the genotyping experiments. RV, SG, JS, PS, CT, SE, GF, OB, JR, RB, AL, DA, JP, CC, GP, PML, MT, RD, CLQ, KS, CVB, CL, KB, HZ, MTK, and FB contributed to sample and phenotype data collection. JS, PJV and SL are the lead PIs for the EMIF-AD MBD study as a whole and were in charge of designing and managing the platform. LB designed and supervised the genomics portion of the EMIF-AD MBD project and co-wrote all drafts of the manuscript. All authors critically revised all manuscripts drafts, read and approved the final manuscript.

## Acknowledgments

The authors acknowledge the assistance of Ellen De Roeck, Naomi De Roeck, and Hanne Struyfs (UAntwerp) with data collection. We thank Mrs. Tanja Wesse and Mrs. Sanaz Sedghpour Sabet at the Institute of Clinical Molecular Biology, Christian-Albrechts-University of Kiel, Kiel, Germany for technical assistance with the GSA genotyping. We thank Dr. Fabian Kilpert for his assistance with the QC and genotype imputations and Marcel Schilling for his assistance on visualizing the results. The LIGA team acknowledges computational support from the OMICS compute cluster at the University of Lübeck.

## References

Armstrong, N. J., Mather, K. A., Sargurupremraj, M., Knol, M. J., Malik, R., Satizabal, C. L., et al. (2020). Common Genetic Variation Indicates Separate Causes for Periventricular and Deep White Matter Hyperintensities. Stroke, 51:7, 2111–2121. doi:10.1161/STROKEAHA.119.027544

Bertram, L., and Tanzi, R. E. (2020). Genomic mechanisms in Alzheimer’s disease. Brain pathology (Zurich, Switzerland), 30:5, 966–977. doi:10.1111/bpa.12882

Bos, I., Vos, S., Vandenberghe, R., Scheltens, P., Engelborghs, S., Frisoni, G., et al. (2018). The EMIF-AD Multimodal Biomarker Discovery study: design, methods and cohort characteristics. Alzheimer’s research and therapy, 10:1, 64. doi:10.1186/s13195-018-0396-5

Boyle, A. P., Hong, E. L., Hariharan, M., Cheng, Y., Schaub, M. A., Kasowski, M., et al. (2012). Annotation of functional variation in personal genomes using RegulomeDB. Genome research, 22:9, 1790–1797. doi:10.1101/gr.137323.112

Buniello, A., MacArthur, J., Cerezo, M., Harris, L. W., Hayhurst, J., Malangone, C., et al. (2019). The NHGRI-EBI GWAS Catalog of published genome-wide association studies, targeted arrays and summary statistics 2019. Nucleic acids research, 47:D1, D1005–D1012. doi:10.1093/nar/gky1120

Bush, W. S., and Moore, J. H. (2012). Chapter 11: Genome-wide association studies. PLoS computational biology, 8:12, e1002822. doi:10.1371/journal.pcbi.1002822

Cacace, R., Sleegers, K., and Van Broeckhoven, C. (2016). Molecular genetics of early-onset Alzheimer’s disease revisited. Alzheimer’s & dementia: the journal of the Alzheimer’s Association, 12:6, 733–748. doi:10.1016/j.jalz.2016.01.012

Choi, S. W., and O’Reilly, P. F. (2019). PRSice-2: Polygenic Risk Score software for biobank-scale data. GigaScience, 8(7), giz082. doi:10.1093/gigascience/giz082

Damale, M. G., Pathan, S. K., Shinde, D. B., Patil, R. H., Arote, R. B., and Sangshetti, J. N. (2020). Insights of tankyrases: A novel target for drug discovery. European journal of medicinal chemistry, 207, 112712. doi:10.1016/j.ejmech.2020.112712

Das, S., Forer, L., Schönherr, S., Sidore, C., Locke, A. E., Kwong, A., et al. (2016). Next-generation genotype imputation service and methods. Nature genetics, 48:10, 1284–1287. doi:10.1038/ng.3656

Davies, G., Lam, M., Harris, S. E., Trampush, J. W., Luciano, M., Hill, W. D., et al. (2018). Study of 300,486 individuals identifies 148 independent genetic loci influencing general cognitive function. Nature communications, 9:1, 2098. doi:10.1038/s41467-018-04362-x

de Leeuw, C. A., Mooij, J. M., Heskes, T., and Posthuma, D. (2015). MAGMA: generalized gene-set analysis of GWAS data. PLoS computational biology, 11:4, e1004219. doi:10.1371/journal.pcbi.1004219

Gao, Q., Mok, H. P., and Zhuang, J. (2019). Secreted modular calcium-binding proteins in pathophysiological processes and embryonic development. Chinese medical journal, 132:20, 2476– 2484. doi:10.1097/CM9.0000000000000472

Gatz, M., Reynolds, C. A., Fratiglioni, L., Johansson, B., Mortimer, J. A., Berg, S., et al. (2006). Role of genes and environments for explaining Alzheimer disease. Archives of general psychiatry, 63:2, 168–174. doi:10.1001/archpsyc.63.2.168

Gottesman, I. I., and Gould, T. D. (2003). The endophenotype concept in psychiatry: etymology and strategic intentions. The American journal of psychiatry, 160:4, 636–645. doi:10.1176/appi.ajp.160.4.636

Graffelman, J., and Weir, B. S. (2016). Testing for Hardy-Weinberg equilibrium at biallelic genetic markers on the X chromosome. Heredity, 116:6, 558–568. doi:10.1038/hdy.2016.20

Grasby, K. L., Jahanshad, N., Painter, J. N., Colodro-Conde, L., Bralten, J., Hibar, D. P., et al. (2020). The genetic architecture of the human cerebral cortex. Science (New York, N.Y.), 367:6484, eaay6690. doi:10.1126/science.aay6690

Hibar, D. P., Adams, H., Jahanshad, N., Chauhan, G., Stein, J. L., Hofer, M. A., et al. (2017). Novel genetic loci associated with hippocampal volume. Nature communications, 8, 13624. doi:10.1038/ncomms13624

Holtzman, D. M., Herz, J., and Bu, G. (2012). Apolipoprotein E and apolipoprotein E receptors: normal biology and roles in Alzheimer disease. Cold Spring Harbor perspectives in medicine, 2:3, a006312. doi:10.1101/cshperspect.a006312

Hong, S., Prokopenko, D., Dobricic, V., Kilpert, F., Bos, I., Vos, S., et al. (2020). Genome-wide association study of Alzheimer’s disease CSF biomarkers in the EMIF-AD Multimodal Biomarker Discovery dataset. Translational psychiatry, 10:1, 403. doi:10.1038/s41398-020-01074-z

Hong, S., Dobricic, V., Ohlei, O., Bos, I., Vos, S., Prokopenko, D., et al. (2021). TMEM106B and CPOX are genetic determinants of cerebrospinal fluid Alzheimer’s disease biomarker levels. Alzheimer’s and dementia: the journal of the Alzheimer’s Association, 17:10, 1628–1640. doi:10.1002/alz.12330

Jansen, I. E., Savage, J. E., Watanabe, K., Bryois, J., Williams, D. M., Steinberg, S., et al. (2019). Genome-wide meta-analysis identifies new loci and functional pathways influencing Alzheimer’s disease risk. Nature genetics, 51:3, 404–413. doi:10.1038/s41588-018-0311-9

Lonsdale, J., Thomas, J., Salvatore, M., Phillips, R., Lo, E., Shad, S., et al. (2013). The Genotype-Tissue Expression (GTEx) project. Nature genetics, 45:6, 580–585. doi:10.1038/ng.2653

Machiela, M. J., and Chanock, S. J. (2015). LDlink: a web-based application for exploring population-specific haplotype structure and linking correlated alleles of possible functional variants. Bioinformatics (Oxford, England), 31:21, 3555–3557. doi:10.1093/bioinformatics/btv402

MacRae, C. A., and Vasan, R. S. (2011). Next-generation genome-wide association studies: time to focus on phenotype?. Circulation. Cardiovascular genetics, 4:4, 334–336. doi:10.1161/CIRCGENETICS.111.960765

Martin, P. M., Carnaud, M., Garcia del Caño, G., Irondelle, M., Irinopoulou, T., Girault, J. A., et al. (2008). Schwannomin-interacting protein-1 isoform IQCJ-SCHIP-1 is a late component of nodes of Ranvier and axon initial segments. The Journal of neuroscience: the official journal of the Society for Neuroscience, 28:24, 6111–6117. doi:10.1523/JNEUROSCI.1044-08.2008

Mattsson, N., Carrillo, M. C., Dean, R. A., Devous, M. D., Sr, Nikolcheva, T., Pesini, P., et al. (2015). Revolutionizing Alzheimer’s disease and clinical trials through biomarkers. Alzheimer’s & dementia (Amsterdam, Netherlands), 1:4, 412–419. doi:10.1016/j.dadm.2015.09.001

Martin, P. M., Cifuentes-Diaz, C., Devaux, J., Garcia, M., Bureau, J., Thomasseau, S., et al. (2017). Schwannomin-interacting Protein 1 Isoform IQCJ-SCHIP1 Is a Multipartner Ankyrin- and Spectrin-binding Protein Involved in the Organization of Nodes of Ranvier. The Journal of biological chemistry, 292:6, 2441–2456. doi:10.1074/jbc.M116.758029

Mazaheri Moghaddam, M., Mazaheri Moghaddam, M., Hamzeiy, H., Baghbanzadeh, A., Pashazadeh, F., and Sakhinia, E. (2021). Genetic basis of acephalic spermatozoa syndrome, and intracytoplasmic sperm injection outcomes in infertile men: a systematic scoping review. Journal of assisted reproduction and genetics, 38:3, 573–586. doi:10.1007/s10815-020-02008-w

McCarthy, S., Das, S., Kretzschmar, W., Delaneau, O., Wood, A. R., Teumer, A., et al. (2016). A reference panel of 64,976 haplotypes for genotype imputation. Nature genetics, 48:10, 1279–1283. doi:10.1038/ng.3643

McLaren, W., Gil, L., Hunt, S. E., Riat, H. S., Ritchie, G. R., Thormann, A., et al. (2016). The Ensembl Variant Effect Predictor. Genome biology, 17:1, 122. doi:10.1186/s13059-016-0974-4

Montani, C., Gritti, L., Beretta, S., Verpelli, C., and Sala, C. (2019). The Synaptic and Neuronal Functions of the X-Linked Intellectual Disability Protein Interleukin-1 Receptor Accessory Protein Like 1 (IL1RAPL1). Developmental neurobiology, 79:1, 85–95. doi:10.1002/dneu.22657

Morkmued, S., Clauss, F., Schuhbaur, B., Fraulob, V., Mathieu, E., Hemmerlé, J., et al. (2020). Deficiency of the SMOC2 matricellular protein impairs bone healing and produces age-dependent bone loss. Scientific reports, 10:1, 14817. doi:10.1038/s41598-020-71749-6

Neu, S. C., Pa, J., Kukull, W., Beekly, D., Kuzma, A., Gangadharan, P., et al. (2017). Apolipoprotein E Genotype and Sex Risk Factors for Alzheimer Disease: A Meta-analysis. JAMA neurology, 74:10, 1178–1189. doi:10.1001/jamaneurol.2017.2188

Omodei, D., Acampora, D., Russo, F., De Filippi, R., Severino, V., Di Francia, R., et al. (2009). Expression of the brain transcription factor OTX1 occurs in a subset of normal germinal-center B cells and in aggressive Non-Hodgkin Lymphoma. The American journal of pathology, 175:6, 2609– 2617. doi:10.2353/ajpath.2009.090542

Patterson, C. (2018) World Alzheimer Report 2018. The State of the Art of Dementia Research: New Frontiers. An Analysis of Prevalence, Incidence, Cost and Trends. Alzheimer’s Disease International.

Purcell, S., Neale, B., Todd-Brown, K., Thomas, L., Ferreira, M. A., Bender, D., et al. (2007). PLINK: a tool set for whole-genome association and population-based linkage analyses. American journal of human genetics, 81:3, 559–575. doi:10.1086/519795

Quintino-Santos, S., Diniz, B. S., Firmo, J. O., Moriguchi, E. H., Lima-Costa, M. F., and Castro-Costa, E. (2015). APOE ε4 allele is associated with worse performance in memory dimensions of the mini-mental state examination: the Bambuì Cohort Study of Aging. International journal of geriatric psychiatry, 30:6, 573–579. doi:10.1002/gps.4186

Rai, A., Bleimling, N., Vetter, I. R., and Goody, R. S. (2020). The mechanism of activation of the actin binding protein EHBP1 by Rab8 family members. Nature communications, 11:1, 4187. doi:10.1038/s41467-020-17792-3

Raymond F. L. (2006). X linked mental retardation: a clinical guide. Journal of medical genetics, 43:3, 193–200. doi:10.1136/jmg.2005.033043

Rentzsch, P., Schubach, M., Shendure, J., and Kircher, M. (2021). CADD-Splice-improving genome-wide variant effect prediction using deep learning-derived splice scores. Genome medicine, 13:1, 31. doi:10.1186/s13073-021-00835-9

Sargurupremraj, M., Suzuki, H., Jian, X., Sarnowski, C., Evans, T. E., Bis, J. C., et al. (2020). Cerebral small vessel disease genomics and its implications across the lifespan. Nature communications, 11:1, 6285. doi:10.1038/s41467-020-19111-2

Scheufele, E., Aronzon, D., Coopersmith, R., McDuffie, M. T., Kapoor, M., Uhrich, C. A., et al. (2014). tranSMART: An Open Source Knowledge Management and High Content Data Analytics Platform. AMIA Joint Summits on Translational Science proceedings. AMIA Joint Summits on Translational Science, 2014, 96–101.

Smith, S. M., Douaud, G., Chen, W., Hanayik, T., Alfaro-Almagro, F., Sharp, K., et al. (2021). An expanded set of genome-wide association studies of brain imaging phenotypes in UK Biobank. Nature neuroscience, 24:5, 737–745. doi:10.1038/s41593-021-00826-4

Sperling, R., Mormino, E., and Johnson, K. (2014). The evolution of preclinical Alzheimer’s disease: implications for prevention trials. Neuron, 84:3, 608–622. doi: 10.1016/j.neuron.2014.10.038

Ten Kate, M., Redolfi, A., Peira, E., Bos, I., Vos, S. J., Vandenberghe, R., et al. (2018). MRI predictors of amyloid pathology: results from the EMIF-AD Multimodal Biomarker Discovery study. Alzheimer’s research & therapy, 10:1, 100. doi:10.1186/s13195-018-0428-1

van der Meer, D., Frei, O., Kaufmann, T., Shadrin, A. A., Devor, A., Smeland, O. B., et al. (2020). Understanding the genetic determinants of the brain with MOSTest. Nature communications, 11:1, 3512. doi:10.1038/s41467-020-17368-1

Watanabe, K., Taskesen, E., van Bochoven, A., and Posthuma, D. (2017). Functional mapping and annotation of genetic associations with FUMA. Nature communications, 8:1, 1826. doi:10.1038/s41467-017-01261-5

Wightman, D. P., Jansen, I. E., Savage, J. E., Shadrin, A. A., Bahrami, S., Holland, D., et al. (2021). A genome-wide association study with 1,126,563 individuals identifies new risk loci for Alzheimer’s disease. Nature genetics, 53:9, 1276–1282. doi:10.1038/s41588-021-00921-z

Willer, C. J., Li, Y., and Abecasis, G. R. (2010). METAL: fast and efficient meta-analysis of genomewide association scans. Bioinformatics (Oxford, England), 26:17, 2190–2191. doi:10.1093/bioinformatics/btq340

Zhang, Q., Cai, Z., Lhomme, M., Sahana, G., Lesnik, P., Guerin, M., et al. (2020). Inclusion of endophenotypes in a standard GWAS facilitate a detailed mechanistic understanding of genetic elements that control blood lipid levels. Scientific reports, 10:1, 18434. doi:10.1038/s41598-020-75612-6

Zhao, B., Zhang, J., Ibrahim, J. G., Luo, T., Santelli, R. C., Li, Y., et al. (2021). Large-scale GWAS reveals genetic architecture of brain white matter microstructure and genetic overlap with cognitive and mental health traits (n = 17,706). Molecular psychiatry, 26:8, 3943–3955. doi:10.1038/s41380-019-0569-z

