## Supplementary Material for "Genome-wide association study of Alzheimer’s disease brain imaging biomarkers and neuropsychological phenotypes in the EMIF-AD Multimodal Biomarker Discovery dataset"

#### Supplementary information

*NB: Some text in the methods description of this document was taken from our previous EMIF-AD MDB GWAS manuscripts Hong et al. (2020) [1] and Hong et al. (2021) [2] without explicit referencing.*

##### Genotype data handling, QC and imputation procedures

Genotyping was performed at the UKSH NGS facility located at the Institute for Clinical Molecular Biology (IKMB) located in Kiel, Germany. All post-genotyping data processing and handling was performed at LIGA located at UKSH campus Lübeck / University of Lübeck.

Raw data processing, i.e. clustering and genotype calling from raw intensity data (idat format) was performed in GenomeStudio software (v2.0.4; Illumina, Inc.) using the genotyping module (version 2.0.2). Samples with call rate <0.95 and p50GC <0.7 were excluded at this stage. We then used PLINK software **v1.9** [3] to perform additional QC filtering, i.e. sex checks (--check-sex 0.25 0.75), strand check (--flip), missing genotype rate (--geno 0.02; --mind 0.05), Hardy-Weinberg equilibrium (HWE) tests (--hwe 0.000005), and minor allele frequency (MAF) filtering (--maf 0.01). For determining pairwise allele sharing (to identify cryptic relatedness), we used an LD-pruned set of markers (--indep-pairwise 1500 150 0.2). Overall, this procedure led to 498,589 QC-filtered SNPs in 931 samples suitable for imputation.

Before imputation, the QC'ed genotype data were then subjected to bcftools (v1.9) [4] for removing ambiguous SNPs and flipping and swapping alleles to align to GRCh37/hg19. This was followed by haplotype phasing using SHAPEIT2 [5] and imputation of unobserved genotypes using Minimac3 [6] using a precompiled Haplotype Reference Consortium (HRC) reference panel (EGAD00001002729 including 39,131,578 SNPs from 11K individuals).

We performed a slightly different QC than Hong et al. (2020) [1] did. Post-imputation of the 39,131,578 SNPs in the 931 samples, we only retained autosomal SNPs with minimac3 $R^2 \geq 0.7$. We then used PLINK software **v2.0** [3] to perform additional QC filtering, i.e. missing genotype rate (--geno 0.02; --mind 0.05), Hardy-Weinberg equilibrium (HWE) tests (--hwe 0.000005), and minor allele frequency (MAF) filtering (--maf 0.01), leaving a total of 7,464,105 SNPs for statistical analyses.

We then performed QC on an individual level with the use of a LD pruned dataset (--indep-pairwise 1500 150 0.2) that includes 198,957 SNPs. It was used for determining pairwise allele-sharing IBD/IBS using (--genome --min 0.1). Furthermore, we excluded all individuals that deviated $>$ 3 SD from the mean of heterozygosity ($\sim$18% in this sample). The LD pruned dataset was also used for principal component analysis (PCA; using PLINK command "–pca") along with the reference dataset of the 1000 Genome Project Consortium Phase 3 (The 1000 Genomes Project Consortium, 2015) to assign ethnic descent groups using the five 1000G super-populations by k-nearest neighbor (k-NN; k=9) classification (using R package "class" [7] in R 2.3.2). This resulted in assigning a "European descent" to 888 out of all 931 samples; only these n=888 samples were used in the subsequent statistical analyses. For neuropsychological phenotypes, n=868 individuals were used because 20 individuals developed a different form of dementia (other than AD) or mental illness during the study and were therefore excluded for these analyses.

#### X Chromosome

In preparation for the analysis of the X chromosomal data, a slightly different approach was followed. After determining the genotypes on the X chromosome, QC of the markers was performed separately on male and female subjects for non-pseudoautosomal regions, using slightly different criteria compared to the autosomes. SNPs were excluded with MAF below 1% in the female group, markers with the genotyping rate below 98%, deviations from HWE in individuals ($P$ <1.0E-04), differential genotyping efficiency between women and men ($P$ <1.0E-04), differential allele frequency between women and men ($P$ <1.0E-04) and for ambiguous allele combinations (A/T and C/G). For the pseudoautosomal regions (PAR1 and PAR2) the same QC was applied as for autosomal SNPs. We aligned the alleles of the remaining SNPs to the reference genome "GRCh37/hg19" before imputation. Imputation of female and male participants were subsequently performed on the Sanger Institute imputation server (https://imputation.sanger.ac.uk/) using the extended HRC reference panel available at that site [8]. After imputation, we used the same QC criteria as for the autosomal SNPs but performed these separately for female and male data sets. For males, markers were coded as 0 vs. 2 (instead of 0 vs. 1), to adjust for the missing second X chromosome (as recommended in Smith et al. (2021) [9]).

To compute the SNP-level association statistics on the X chromosome, we used linear regression models in PLINK2 (command: --glm), including the same covariates as for the analyses of the autosomal SNPs. To explore associations on the X chromosome that are driven by genetic sex, we conducted two additional analyses restricted to just genetic females and (separately) just genetic males. We then combined these two additional GWAS in a meta-analysis using Stouffer's method implemented in METAL [10]. Results from X chromosome analyses using linear regression were compared to a meta-analysis using Stouffer's method. We found no additional genome-wide significant signals using Stouffer's method, so only the results from linear regression using PLINK2 are shown in this study.

#### Neuropsychological phenotypes

For cross-sectional analyses, $z$-scores derived from baseline data were used. In the "MMSE" domain all samples were tested with the "Mini Mental State Examination" (MMSE). For the trait "attention" the tests "Trail Making Test A" (726 samples), "Repeatable Battery for the Assessment of Neuropsychological Status (RBANS): index concentration" (46), "Hasegawa Dementia Scale (HDS) concentration index" (27), and "Wechsler Memory Scale" (WMS, 33) were used. For the trait "executive functioning" the tests "Trail Making Test B" (546), and "Stroop part 3" (1) were used. For the trait "language" the tests "Animals Fluency 1 min" (590), "Category Fluency Sum of 3" (125), "Boston Naming Test" (BNT, 102), "RBANS: index language" (28), and "Animals Fluency 2 min" (4) were used. For the trait "delayed memory" the tests "Auditory Verbal Learning Test" (AVLT, 446), "Consortium to Establish a Registry for AD (CERAD) Wordlist" (150), "Free and Cued Selective Reminding Test" (FCSRT, 102), "RI-48" (22), and "MMSE Memory" (17) were used. For the trait "immediate memory" the tests "Auditory Verbal Learning Test" (AVLT, 446), "CERAD Wordlist" (156), "FCSRT" (96), "WMS" (51), "HDS: recent memory item 20" (27), and "RI-48" (22) were used. For the trait "visuoconstruction" the tests "Complex Figure Test Copy" (204), "CERAD Figures" (189), "Rbans: index visuoconstruction" (48), "HDS: drawing item 15" (30), and "MMSE Figure" (2) were used. More details on the neuropsychological testing performed at each site and procedures to harmonize data across sites can be found in Bos et al. (2018) [11]. For all seven neuropsychological traits follow-up data from at least one additional time point were available for each individual and used to construct a longitudinal phenotype using the following formula:

$$\frac{(Score_{last} - Score_{first})}{(interval \times \frac{(Score_{last}+Score_{first})}{2}))} \quad (1)$$

When calculating longitudinal phenotypes, this formula was applied separately for each neuropsychological test. Outlying scores were determined using false discovery rate (FDR) <0.05 estimations and were excluded from all subsequent analyses. Only the most frequently used tests per cognitive domain were included in the final phenotypes. For the trait "MMSE" all 520 individuals were tested with the test "MMSE". For the trait "attention" all 402 individuals were tested with "Trail Making Test A". For the trait "executive functioning" all 234 individuals were tested with "Trail Making Test B". For the trait "language" 297 samples were tested with "Animals Fluency 1 min" and 112 with "Category Fluency Sum of 3". For the trait "delayed memory" 251 samples were tested with "AVLT" and 86 with "CERAD Wordlist". For the trait "immediate memory" 255 samples were tested with "AVLT" and 90 with "CERAD Wordlist". For the trait "visuoconstruction" 104 samples were tested with "CERAD Figures" and 45 with "Complex Figure Test Copy". Both baseline and longitudinal phenotypes were adjusted for age at baseline. More details on the collected neurocognitive phenotypes can be found in Bos et al. (2018) [11].

#### Supplementary References


1. Hong, S., Prokopenko, D., Dobricic, V., Kilpert, F., Bos, I., Vos, Set al. (2020). Genome-wide association study of Alzheimer's disease CSF biomarkers in the EMIF-AD Multimodal Biomarker Discovery dataset. *Translational psychiatry*, 10:1, 403. doi:10.1038/s41398-020-01074-z

2. Hong, S., Dobricic, V., Ohlei, O., Bos, I., Vos, S., Prokopenko et al. (2021). TMEM106B and CPOX are genetic determinants of cerebrospinal fluid Alzheimer's disease biomarker levels. *Alzheimer's and dementia: the journal of the Alzheimer's Association*, 17:10, 1628–1640. doi:10.1002/alz.12330

3. Purcell, S., Neale, B., Todd-Brown, K., Thomas, L., Ferreira, M. A., Bender, D et al. (2007). PLINK: a tool set for whole-genome association and population-based linkage analyses. *American journal of human genetics*, 81:3, 559–575. doi:10.1086/519795

4. Narasimhan, V., Danecek, P., Scally, A., Xue, Y., Tyler-Smith, C. and Durbin, R. (2016). BCFtools/RoH: a hidden Markov model approach for detecting autozygosity from next-generation sequencing data. *Bioinformatics (Oxford, England)*, 32:11, 1749–1751. doi:10.1093/bioinformatics/btw044

5. Delaneau, O., Marchini, J. and Zagury, J. F. (2011). A linear complexity phasing method for thousands of genomes. *Nature methods*, 9:2, 179–181. doi:10.1038/nmeth.1785

6. Das, S., Forer, L., Schönherr, S., Sidore, C., Locke, A. E., Kwong, A. et al. (2016). Next-generation genotype imputation service and methods. *Nature genetics*, 48:10, 1284–1287. doi:10.1038/ng.3656

7. Venables WN, Springer BDR. Modern Applied Statistics with S Fourth edition. http://www.insightful.com. (accessed May 24, 2019)

8. McCarthy, S., Das, S., Kretzschmar, W., Delaneau, O., Wood, A. R., Teumer, A. et al. (2016). A reference panel of 64,976 haplotypes for genotype imputation. *Nature genetics*, 48:10, 1279–1283. doi:10.1038/ng.3643

9. Smith, S. M., Douaud, G., Chen, W., Hanayik, T., Alfaro-Almagro, F., Sharp, K. et al. (2021). An expanded set of genome-wide association studies of brain imaging phenotypes in UK Biobank. *Nature neuroscience*, 24:5, 737–745. doi:10.1038/s41593-021-00826-4

10. Willer, C. J., Li, Y., and Abecasis, G. R. (2010). METAL: fast and efficient meta-analysis of genomewide association scans. *Bioinformatics (Oxford, England)*, 26:17, 2190–2191. doi:10.1093/bioinformatics/btq340

11. Bos, I., Vos, S., Vandenberghe, R., Scheltens, P., Engelborghs, S., Frisoni, G. et al. (2018). The EMIF-AD Multimodal Biomarker Discovery study: design, methods and cohort characteristics. *Alzheimer's research and therapy*, 10:1, 64. doi:10.1186/s13195-018-0396-5

**Supplementary Tables**

Supplementary Tables 1-24 can be found in the MS-Excel file ”Supplementary_Tables.xls”.

**Supplementary Figures**

Supplementary Figures with descriptions can be found on the following pages.

### 1 GWAS - neuropsychological phenotypes

#### 1.1 Aggregated Q-Q plots

##### 1.1.1 Cross-sectional

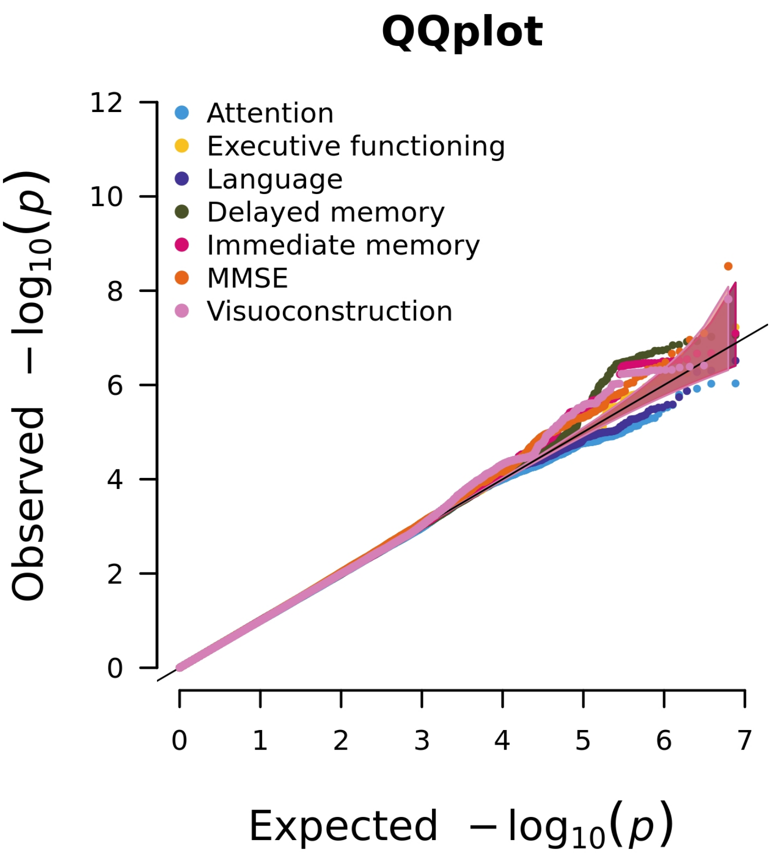


Figure 1: Aggregated Q-Q plot for the analyses of the seven cross-sectional neuropsychological phenotypes

##### 1.1.2 Longitudinal

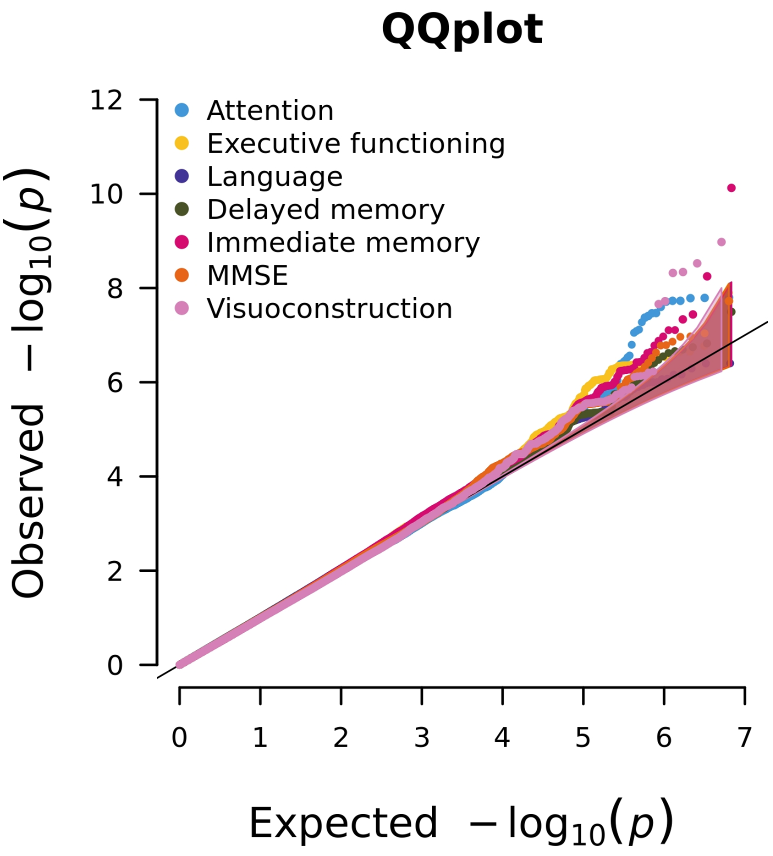


Figure 2: Aggregated Q-Q plot for the analyses of the seven longitudinal neuropsychological phenotypes

#### 1.2 MMSE

##### 1.2.1 Cross-sectional

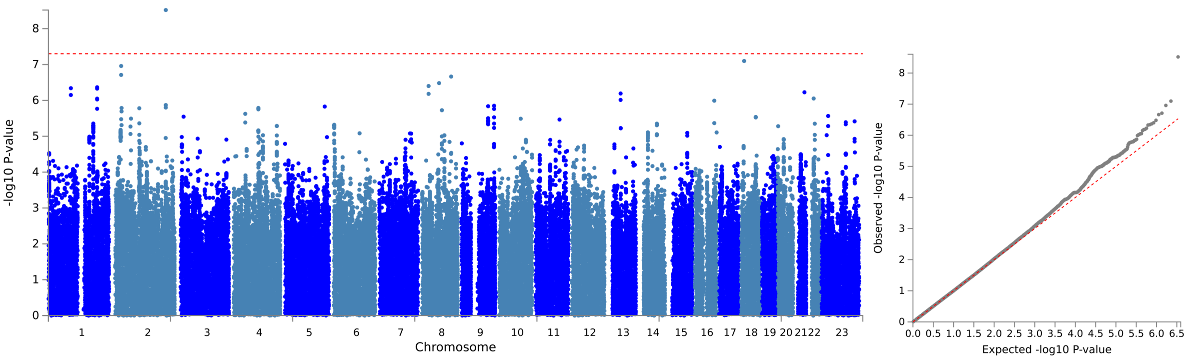


(a) Manhattan plot (b) Q-Q plot

Figure 3: Visualisation of the GWAS results with the cross-sectional data for the cognitive phenotype MMSE

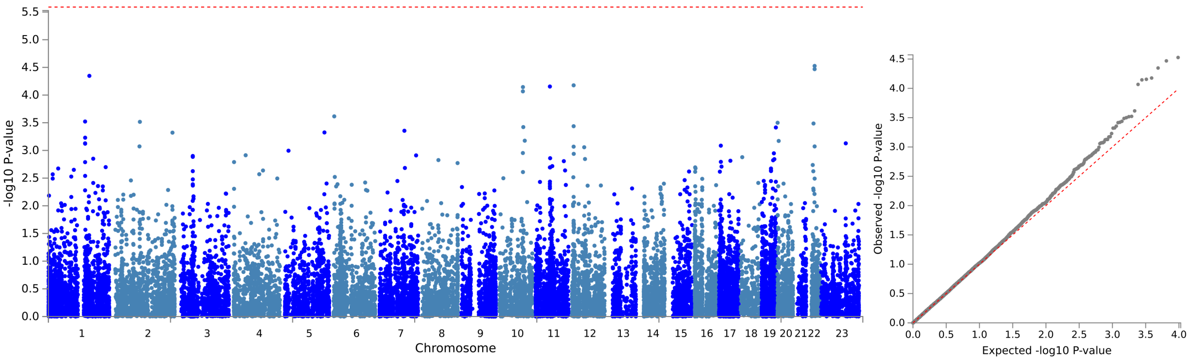


(a) Manhattan plot (b) Q-Q plot

Figure 4: Visualisation of the results of the gene-based analyses with the cross-sectional data for the cognitive phenotype MMSE

##### 1.2.2 Longitudinal

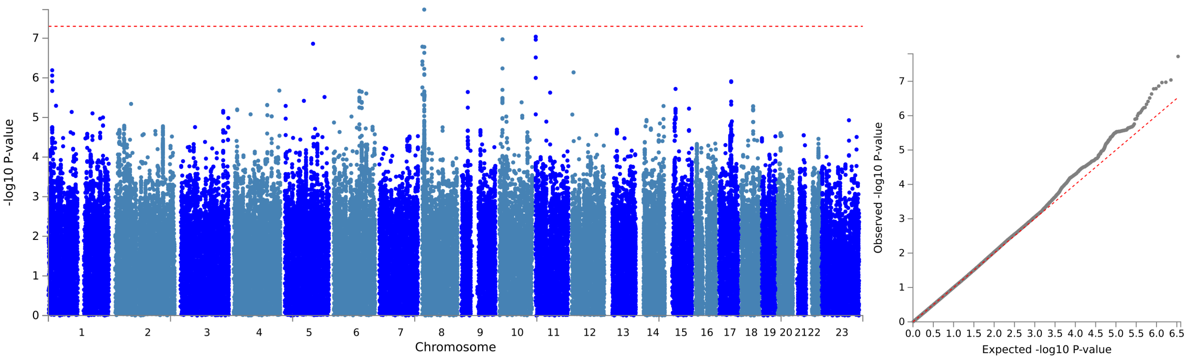


(a) Manhattan plot (b) Q-Q plot

Figure 5: Visualisation of the GWAS results with the longitudinal data for the cognitive phenotype MMSE

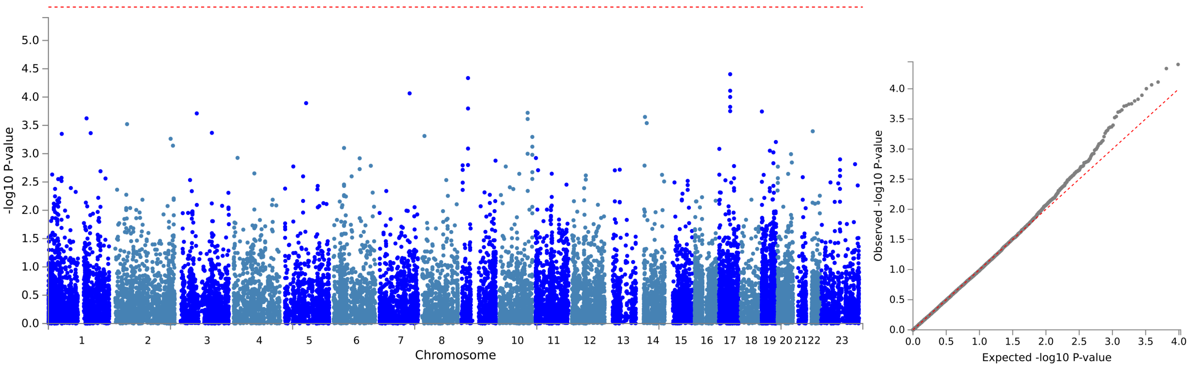


(a) Manhattan plot (b) Q-Q plot

Figure 6: Visualisation of the GWAS results with the longitudinal data for the cognitive phenotype MMSE

#### 1.3 Attention

##### 1.3.1 Cross-sectional

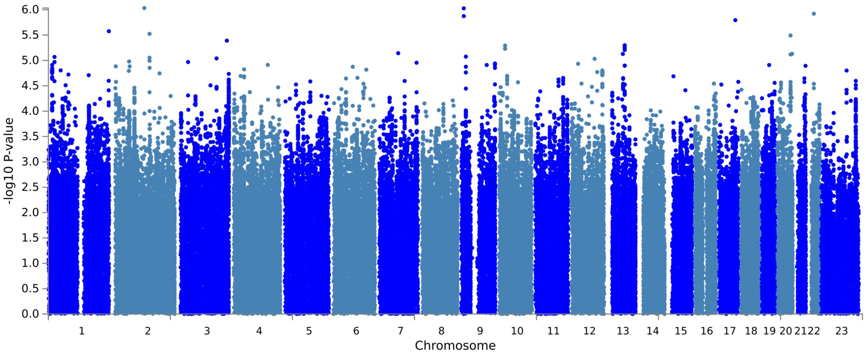


(a) Manhattan plot

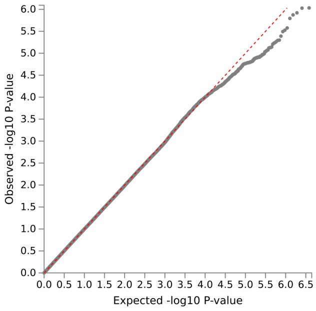


(b) Q-Q plot

Figure 7: Visualisation of the GWAS results with the cross-sectional data for the cognitive phenotype attention

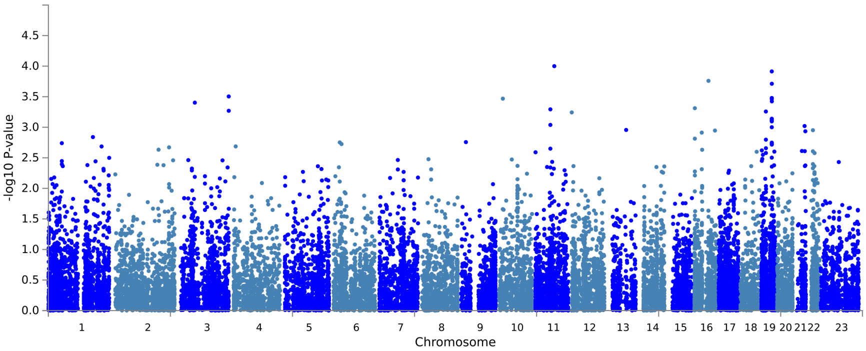


(a) Manhattan plot

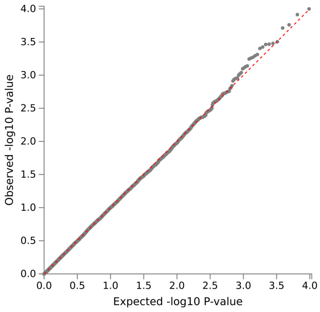


(b) Q-Q plot

Figure 8: Visualisation of the results of the gene-based analyses with the cross-sectional data for the cognitive phenotype attention

##### 1.3.2 Longitudinal

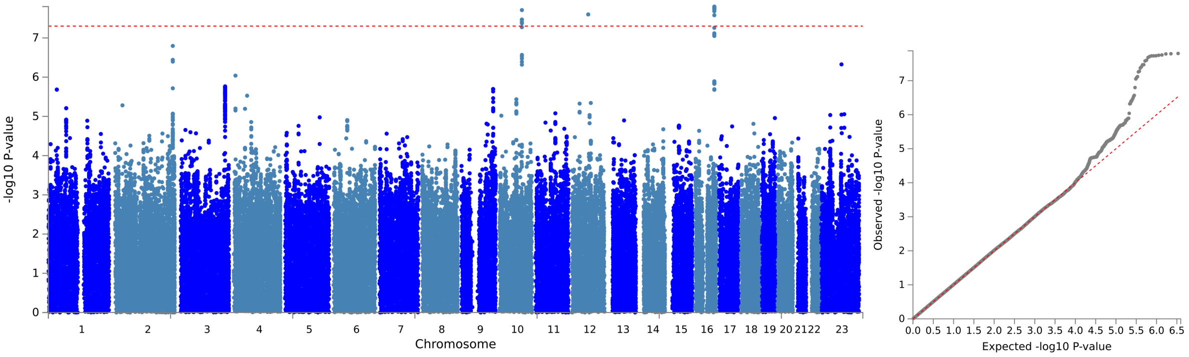


(a) Manhattan plot (b) Q-Q plot

Figure 9: Visualisation of the GWAS results with the longitudinal data for the cognitive phenotype attention

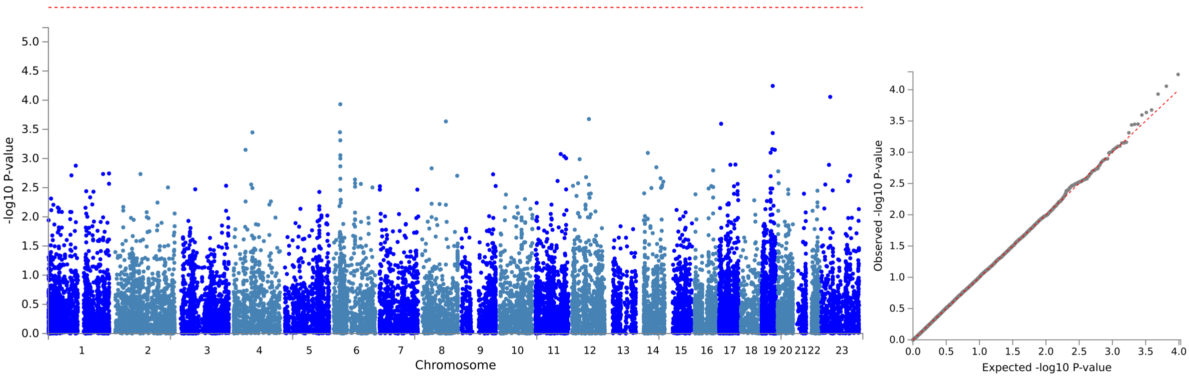


(a) Manhattan plot (b) Q-Q plot

Figure 10: Visualisation of the GWAS results with the longitudinal data for the cognitive phenotype attention

#### 1.4 Executive functioning

##### 1.4.1 Cross-sectional

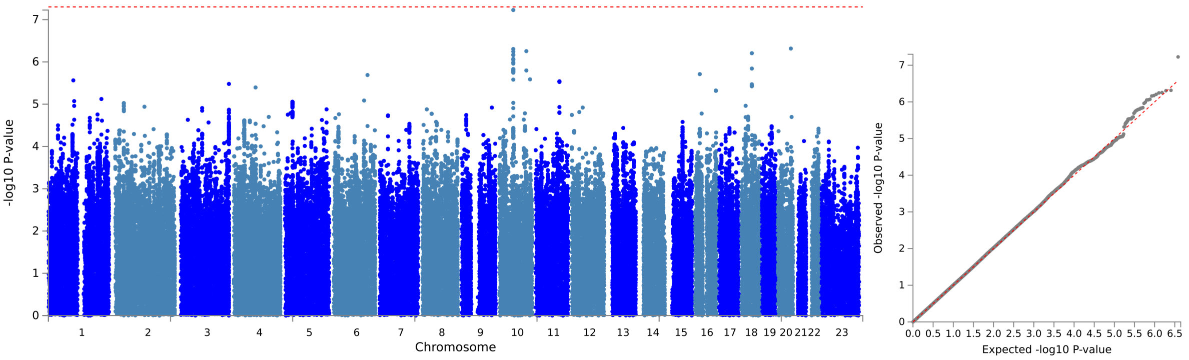


(a) Manhattan plot (b) Q-Q plot

Figure 11: Visualisation of the GWAS results with the cross-sectional data for the cognitive phenotype for the cognitive domain executive functioning

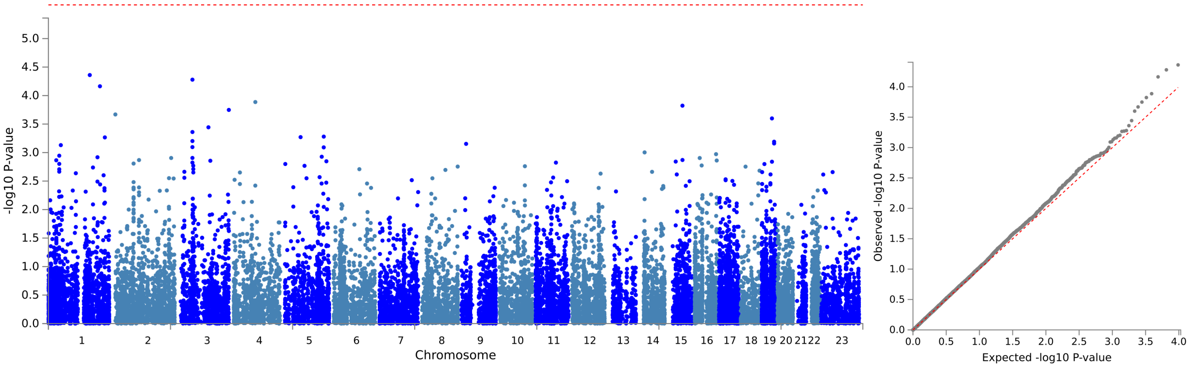


(a) Manhattan plot (b) Q-Q plot

Figure 12: Visualisation of the results of the gene-based analyses with the cross-sectional data for the cognitive phenotype executive functioning

##### 1.4.2 Longitudinal

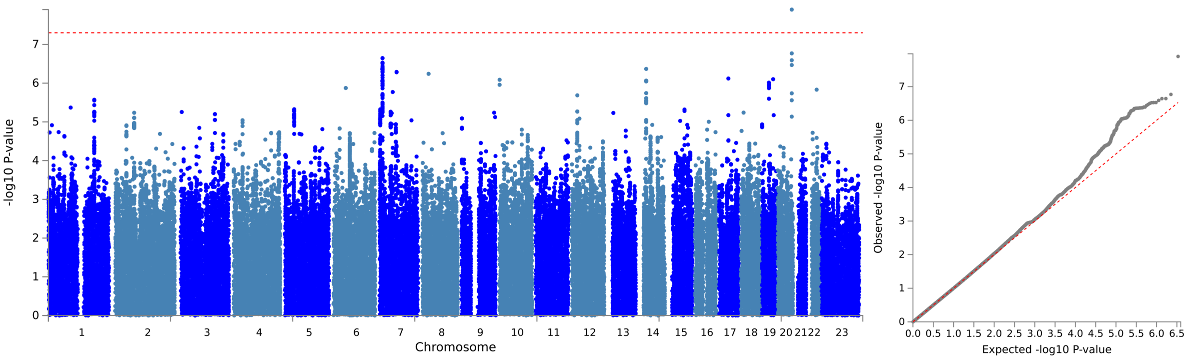


(a) Manhattan plot (b) Q-Q plot

Figure 13: Visualisation of the GWAS results with the longitudinal data for the cognitive phenotype executive functioning

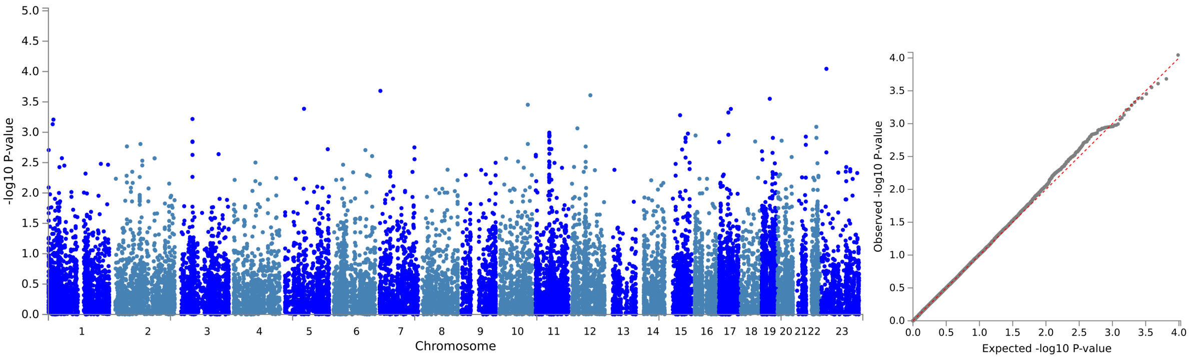


(a) Manhattan plot (b) Q-Q plot

Figure 14: Visualisation of the GWAS results with the longitudinal data for the cognitive phenotype executive functioning

#### 1.5 Language

##### 1.5.1 Cross-sectional

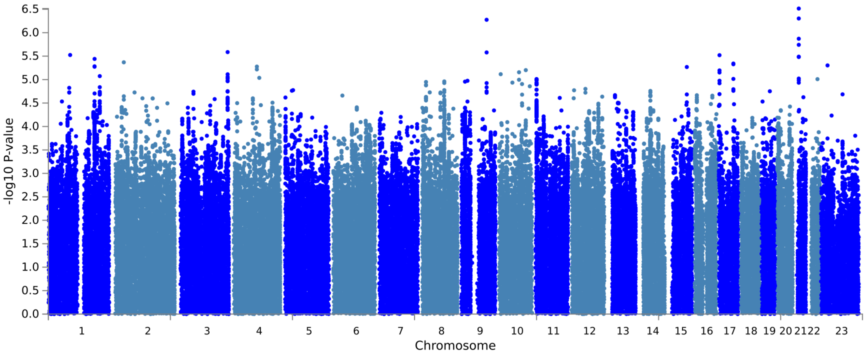


(a) Manhattan plot

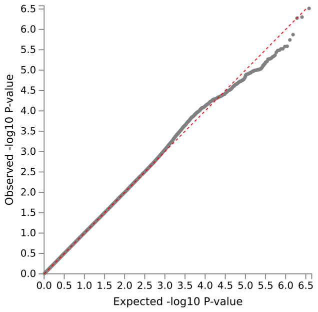


(b) Q-Q plot

Figure 15: Visualisation of the GWAS results with the cross-sectional data for the cognitive phenotype for the cognitive domain language

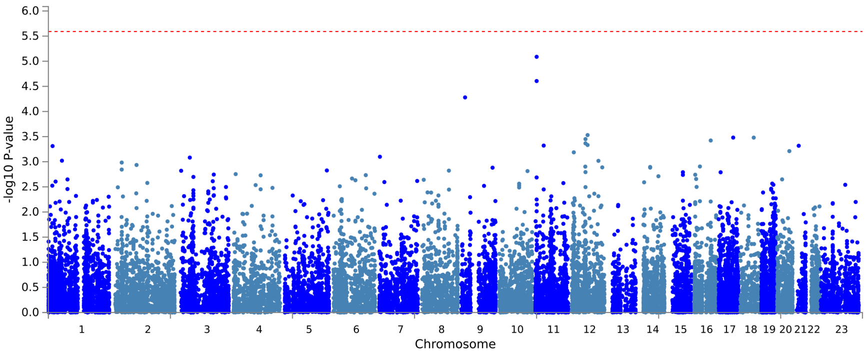


(a) Manhattan plot

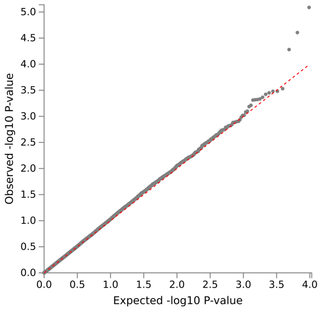


(b) Q-Q plot

Figure 16: Visualisation of the results of the gene-based analyses with the cross-sectional data for the cognitive phenotype language

##### 1.5.2 Longitudinal

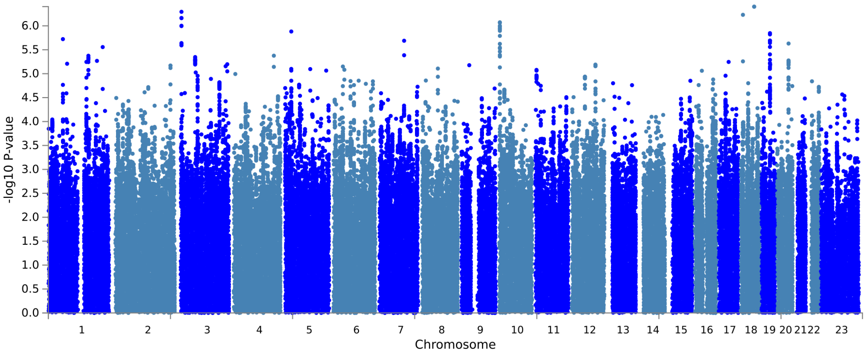


(a) Manhattan plot

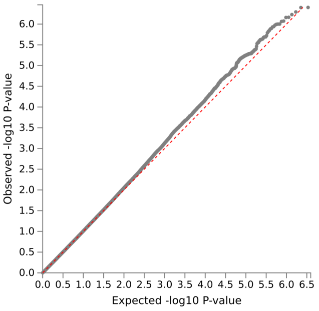


(b) Q-Q plot

Figure 17: Visualisation of the GWAS results with the longitudinal data for the cognitive phenotype language

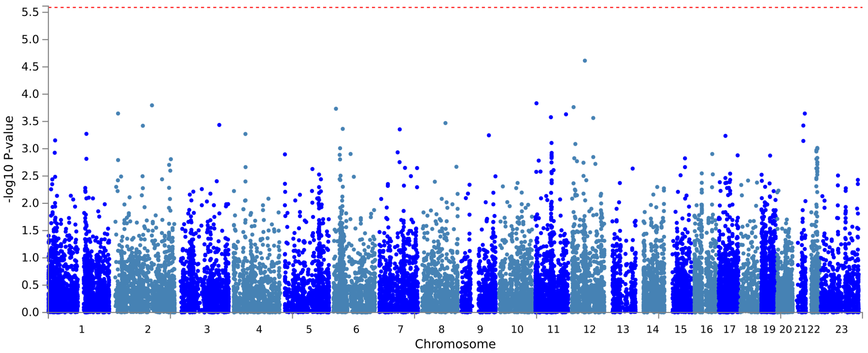


(a) Manhattan plot

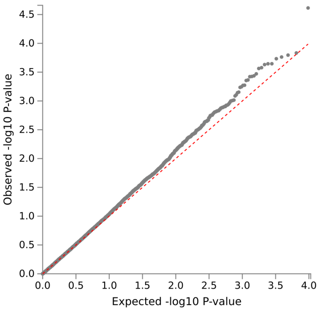


(b) Q-Q plot

Figure 18: Visualisation of the GWAS results with the longitudinal data for the cognitive phenotype language

#### 1.6 Delayed memory

##### 1.6.1 Cross-sectional

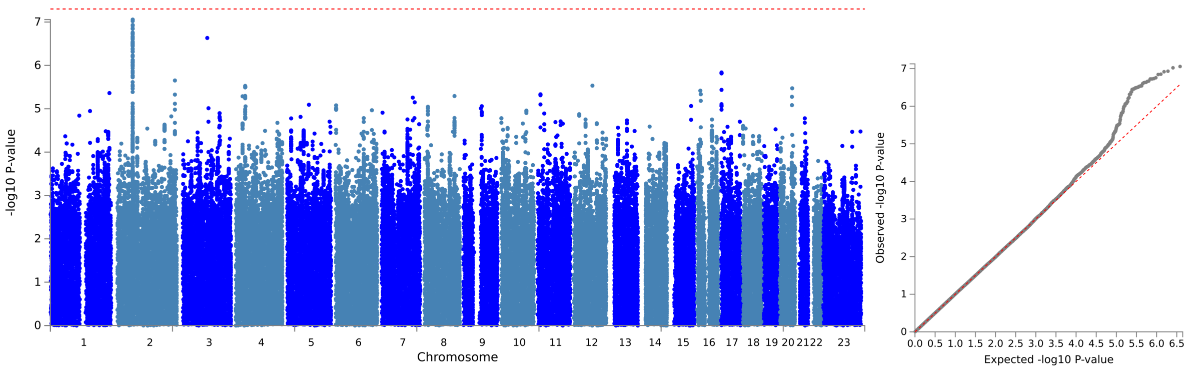


(a) Manhattan plot (b) Q-Q plot

Figure 19: Visualisation of the GWAS results with the cross-sectional data for the cognitive phenotype for the cognitive domain delayed memory

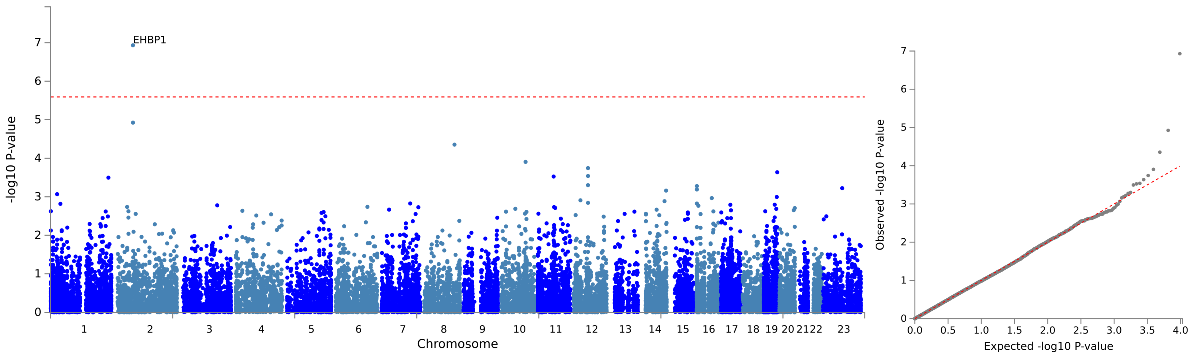


(a) Manhattan plot (b) Q-Q plot

Figure 20: Visualisation of the results of the gene-based analyses with the cross-sectional data for the cognitive phenotype delayed memory

##### 1.6.2 Longitudinal

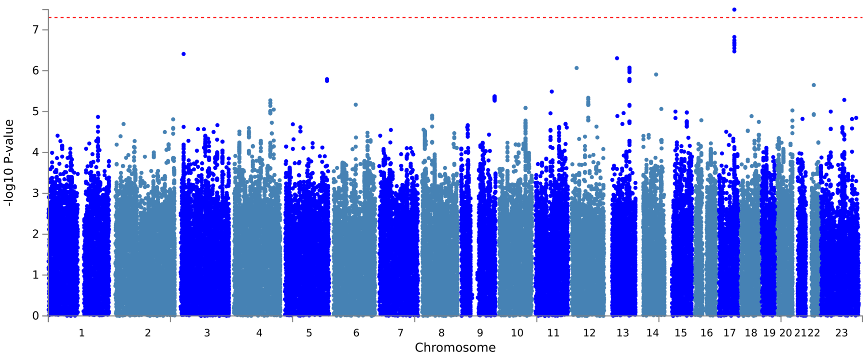


(a) Manhattan plot

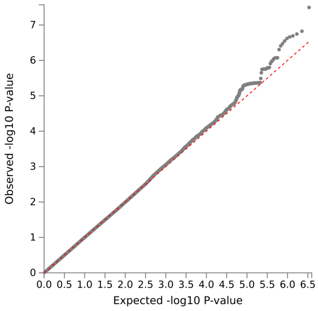


(b) Q-Q plot

Figure 21: Visualisation of the GWAS results with the longitudinal data for the cognitive phenotype delayed memory

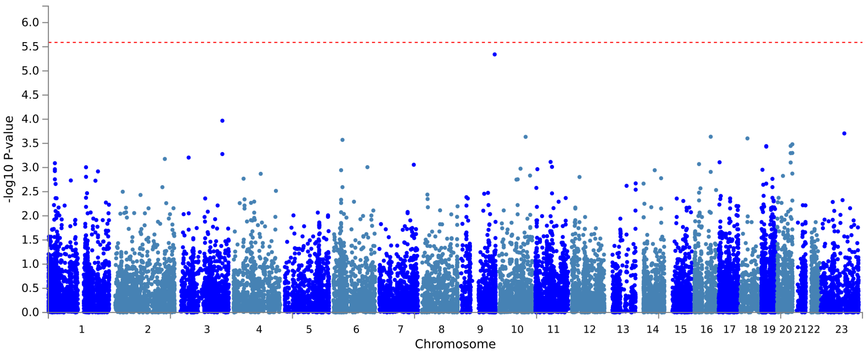


(a) Manhattan plot

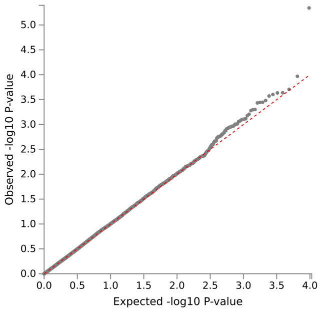


(b) Q-Q plot

Figure 22: Visualisation of the GWAS results with the longitudinal data for the cognitive phenotype delayed memory

#### 1.7 Immediate memory

##### 1.7.1 Cross-sectional

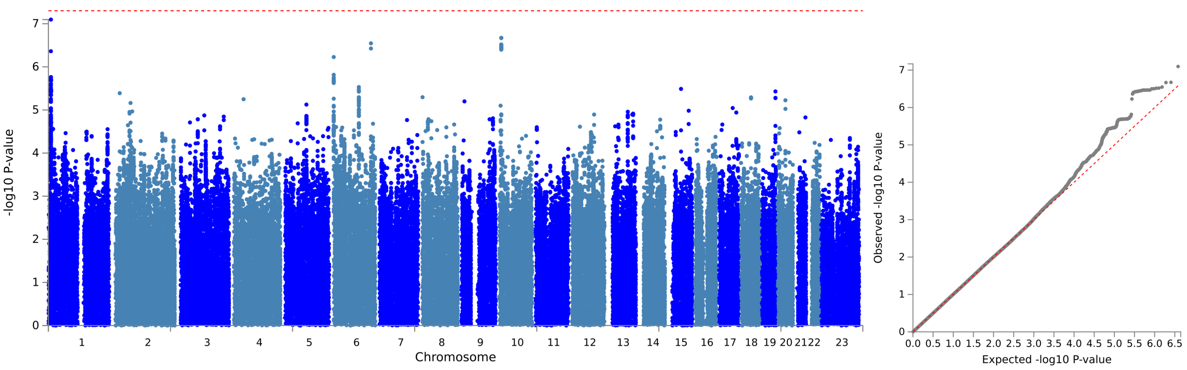


(a) Manhattan plot (b) Q-Q plot

Figure 23: Visualisation of the GWAS results with the cross-sectional data for the cognitive phenotype immediate memory

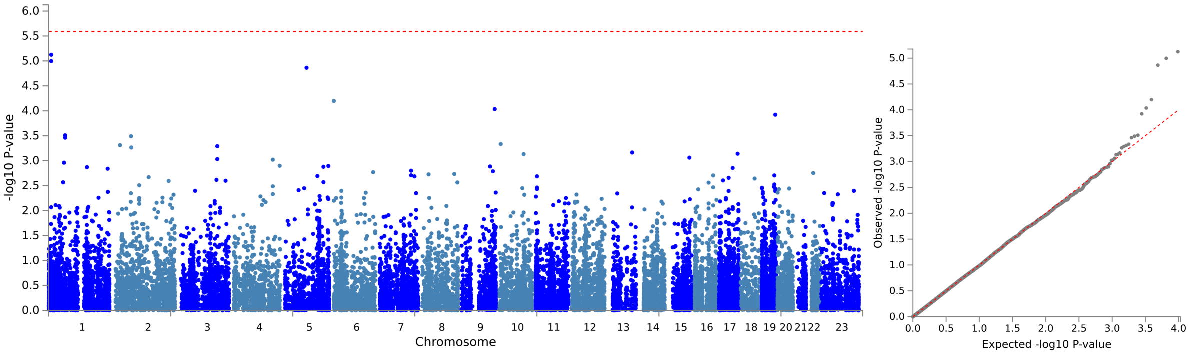


(a) Manhattan plot (b) Q-Q plot

Figure 24: Visualisation of the results of the gene-based analyses with the cross-sectional data for the cognitive phenotype immediate memory

##### 1.7.2 Longitudinal

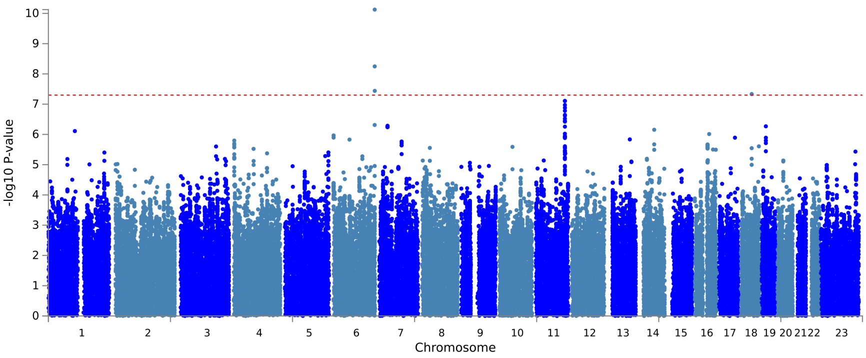


(a) Manhattan plot

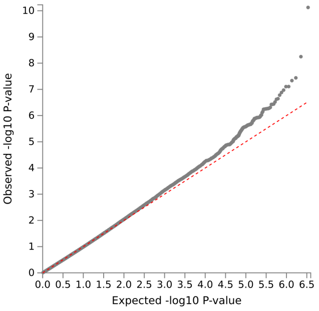


(b) Q-Q plot

Figure 25: Visualisation of the GWAS results with the longitudinal data for the cognitive phenotype immediate

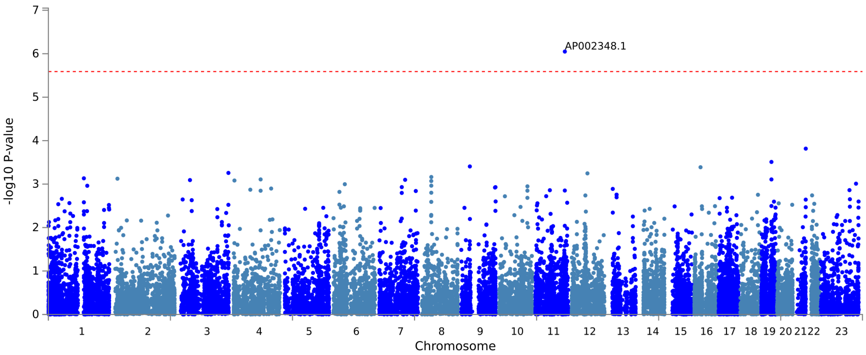


(a) Manhattan plot

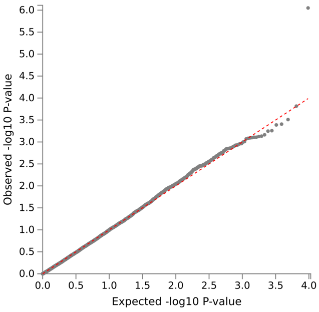


(b) Q-Q plot

Figure 26: Visualisation of the GWAS results with the longitudinal data for the cognitive phenotype immediate

#### 1.8 Visuoconstruction

##### 1.8.1 Cross-sectional

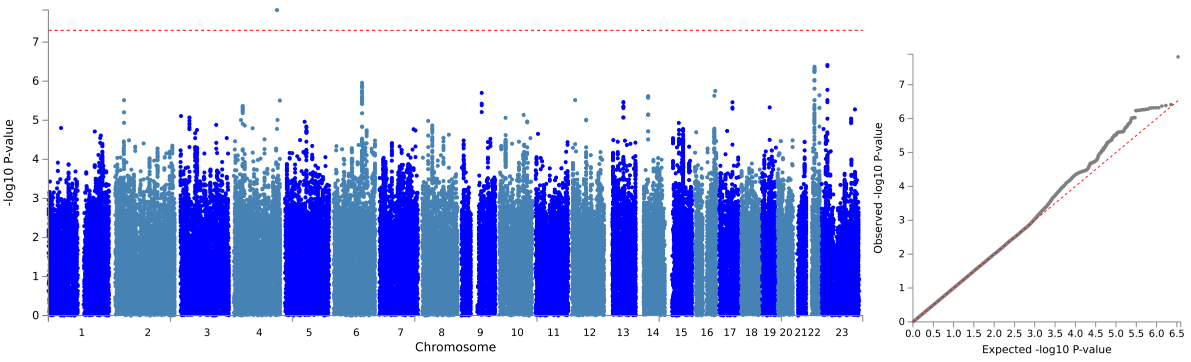


(a) Manhattan plot (b) Q-Q plot

Figure 27: Visualisation of the GWAS results with the cross-sectional data for the cognitive phenotype for the cognitive domain visuoconstruction

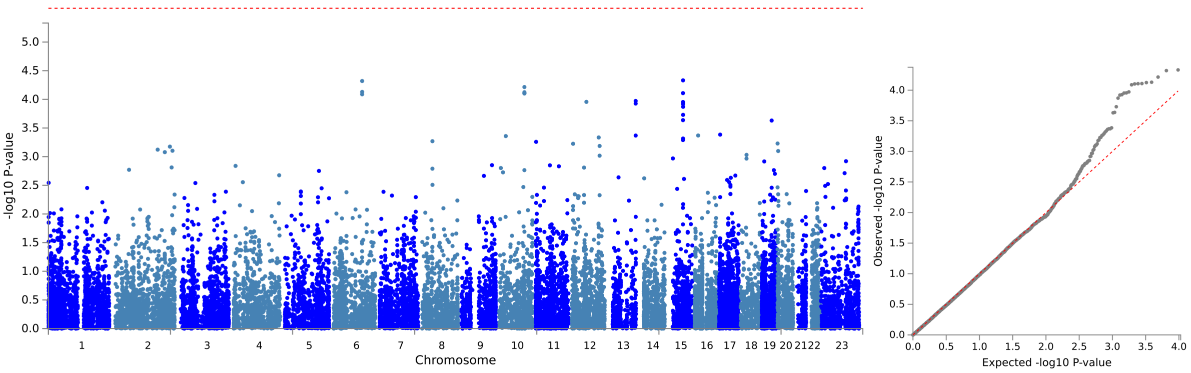


(a) Manhattan plot (b) Q-Q plot

Figure 28: Visualisation of the results of the gene-based analyses with the cross-sectional data for the cognitive phenotype visuoconstruction

##### 1.8.2 Longitudinal

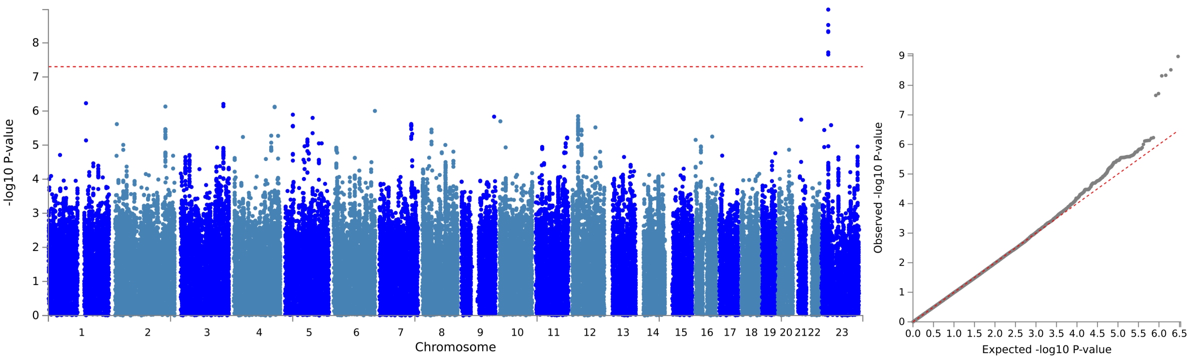


(a) Manhattan plot (b) Q-Q plot

Figure 29: Visualisation of the GWAS results with the longitudinal data for the cognitive phenotype visuoconstruction

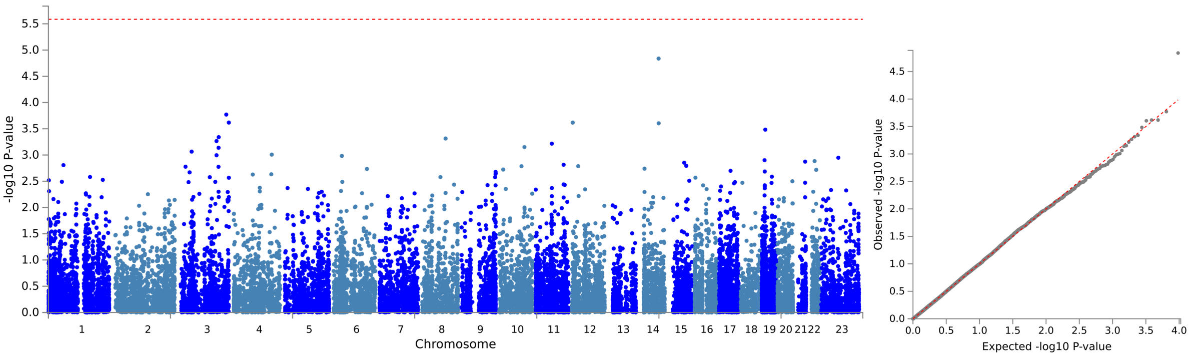


(a) Manhattan plot (b) Q-Q plot

Figure 30: Visualisation of the results of the gene-based analyses with the longitudinal data for the cognitive phenotype visuoconstruction

#### 2 GWAS - MRI phenotypes

##### 2.1 Fazekas

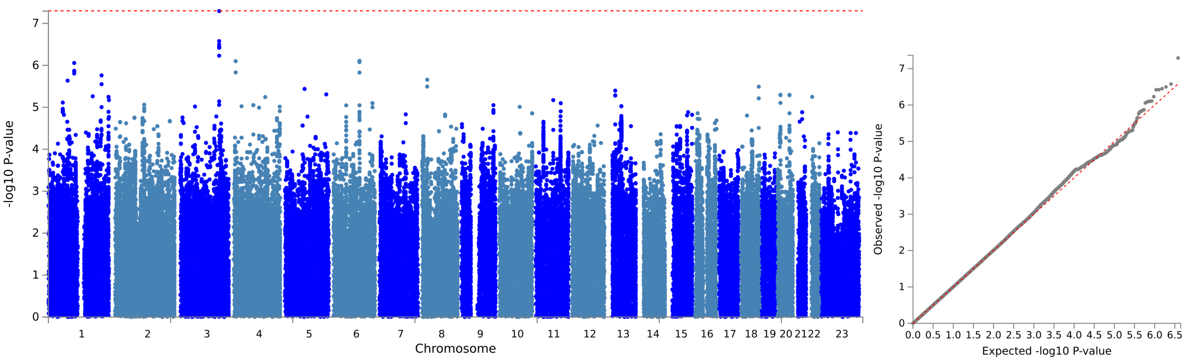


(a) Manhattan plot (b) Q-Q plot

Figure 31: Visualisation of the GWAS results with the cross-sectional data for the MRI phenotype Fazekas

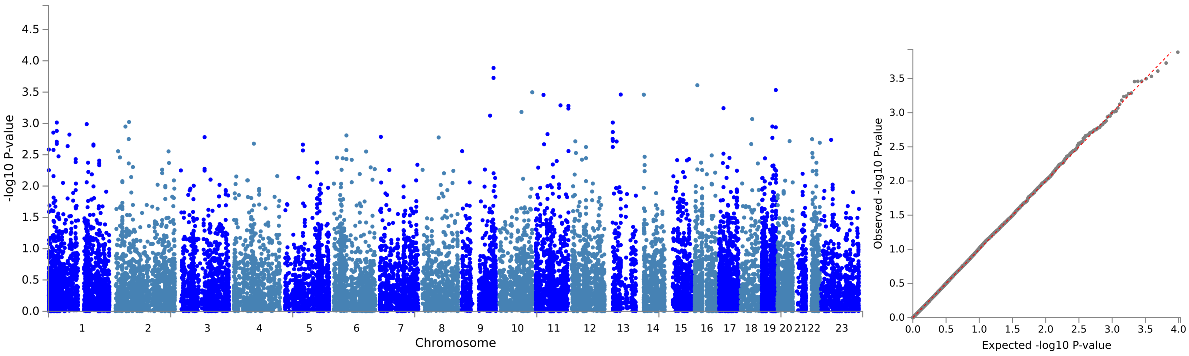


(a) Manhattan plot (b) Q-Q plot

Figure 32: Visualisation of the results of the gene-based analyses with the cross-sectional data for the MRI phenotype Fazekas

#### 2.2 Hippocampus volume sum

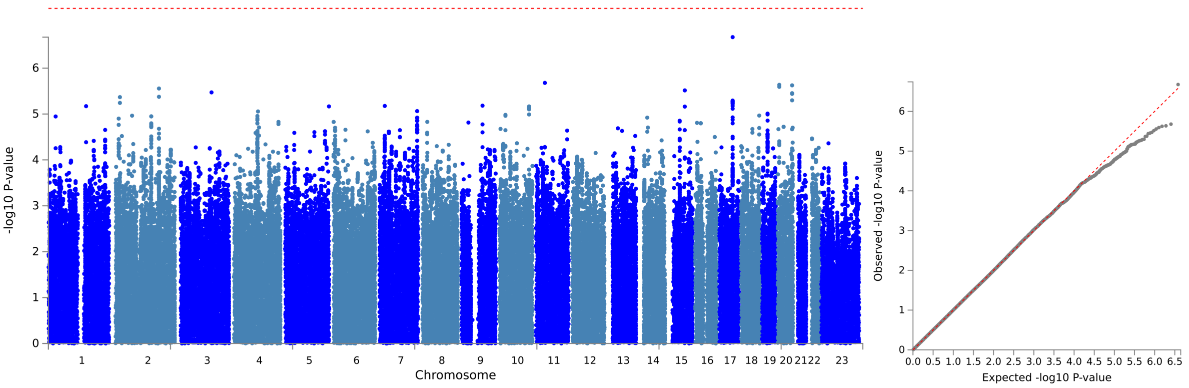


(a) Manhattan plot (b) Q-Q plot

Figure 33: Visualisation of the GWAS results with the cross-sectional data for the MRI phenotype hippocampus volume sum

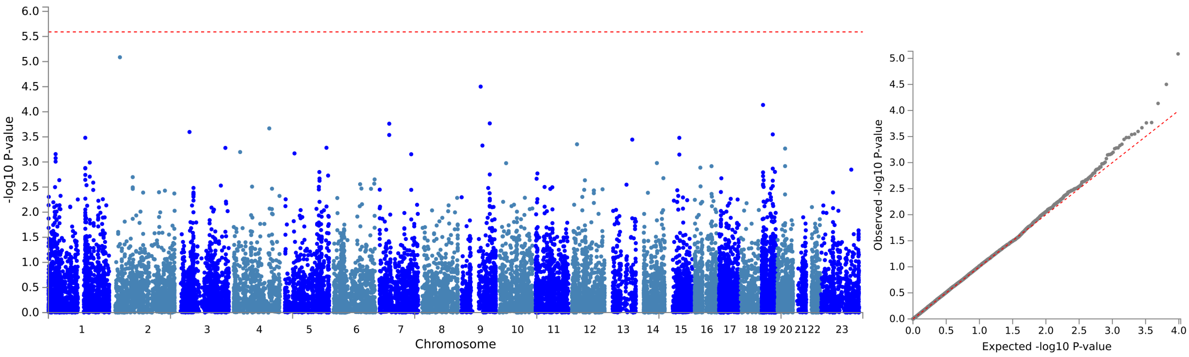


(a) Manhattan plot (b) Q-Q plot

Figure 34: Visualisation of the results of the gene-based analyses with the cross-sectional data for the MRI phenotype hippocampus volume sum

#### 2.3 Hippocampus volume left

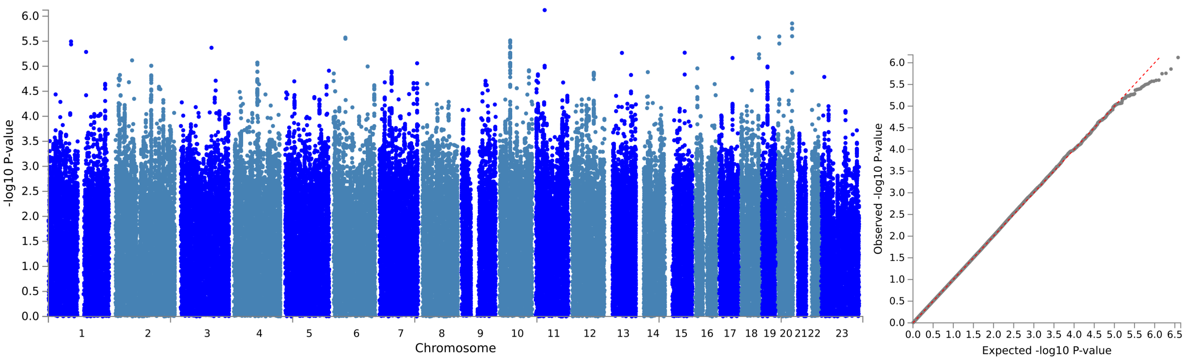


(a) Manhattan plot (b) Q-Q plot

Figure 35: Visualisation of the GWAS results with the cross-sectional data for the MRI phenotype hippocampus volume left

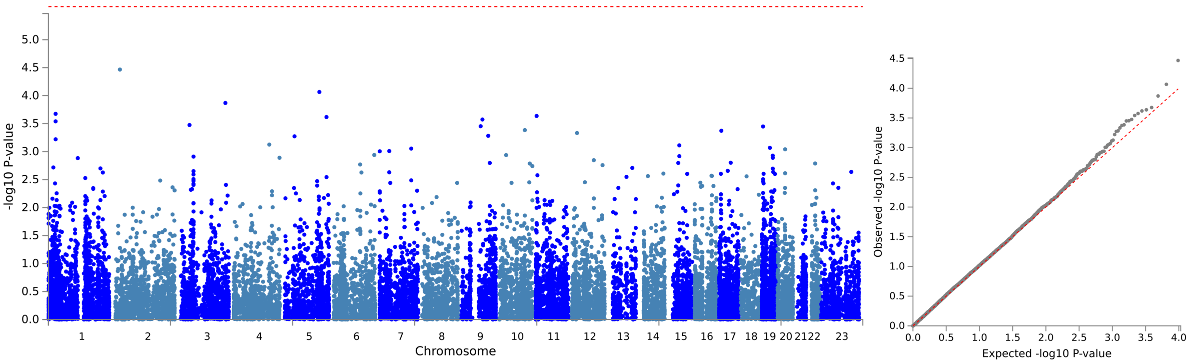


(a) Manhattan plot (b) Q-Q plot

Figure 36: Visualisation of the results of the gene-based analyses with the cross-sectional data for the MRI phenotype hippocampus volume left

#### 2.4 Hippocampus volume right

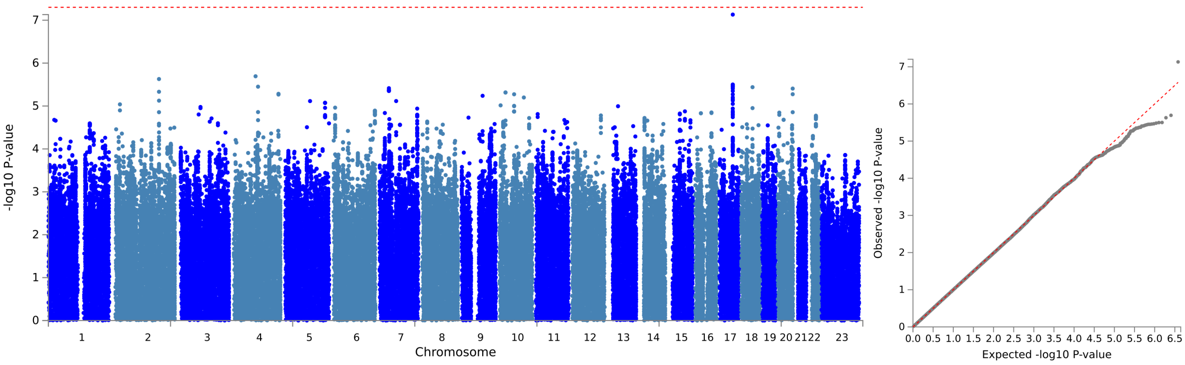


(a) Manhattan plot (b) Q-Q plot

Figure 37: Visualisation of the GWAS results with the cross-sectional data for the MRI phenotype hippocampus volume right

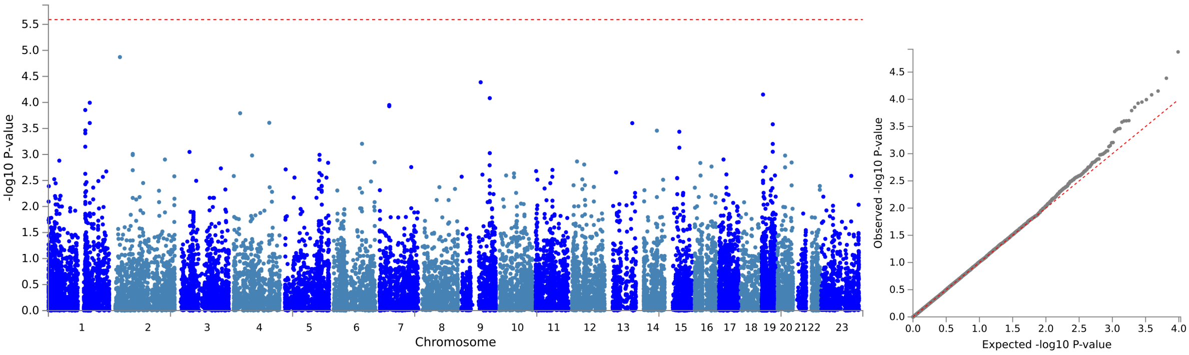


(a) Manhattan plot (b) Q-Q plot

Figure 38: Visualisation of the results of the gene-based analyses with the cross-sectional data for the MRI phenotype hippocampus volume right

#### 2.5 Thickness

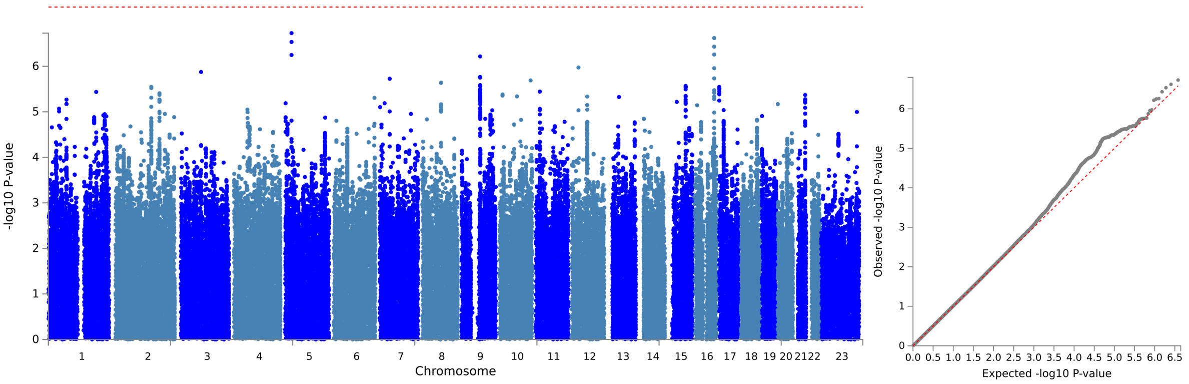


(a) Manhattan plot (b) Q-Q plot

Figure 39: Visualisation of the GWAS results with the cross-sectional data for the MRI phenotype thickness

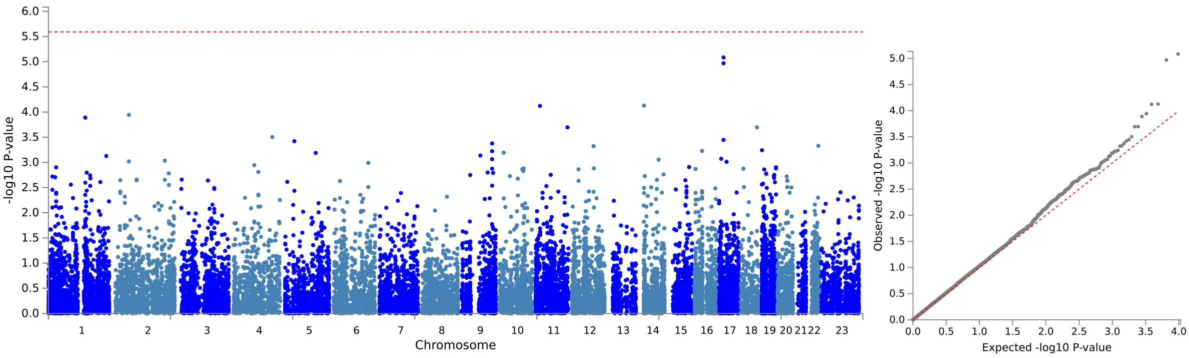


(a) Manhattan plot (b) Q-Q plot

Figure 40: Visualisation of the results of the gene-based analyses with the cross-sectional data for the MRI phenotype thickness

### 3 PGS - neuropsychological phenotypes

#### 3.1 PGS with Jansen GWAS

##### 3.1.1 MMSE

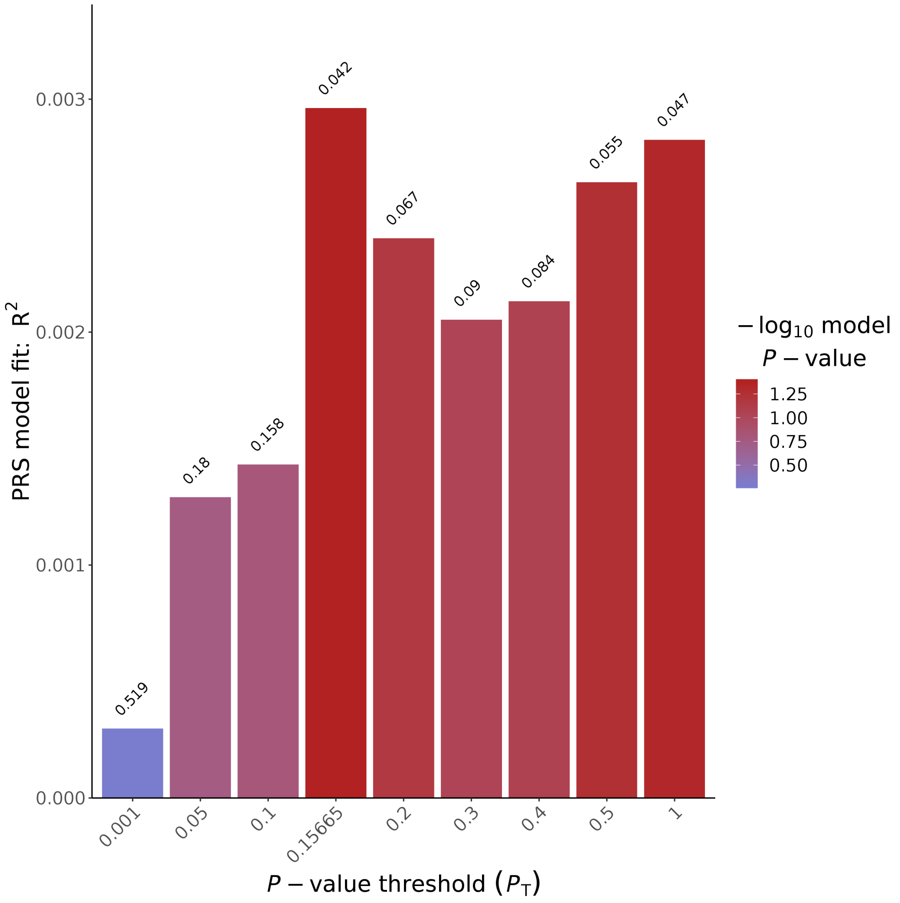


Figure 41: PGS for MMSE (cross-sectional) with SNPs from the Jansen GWAS

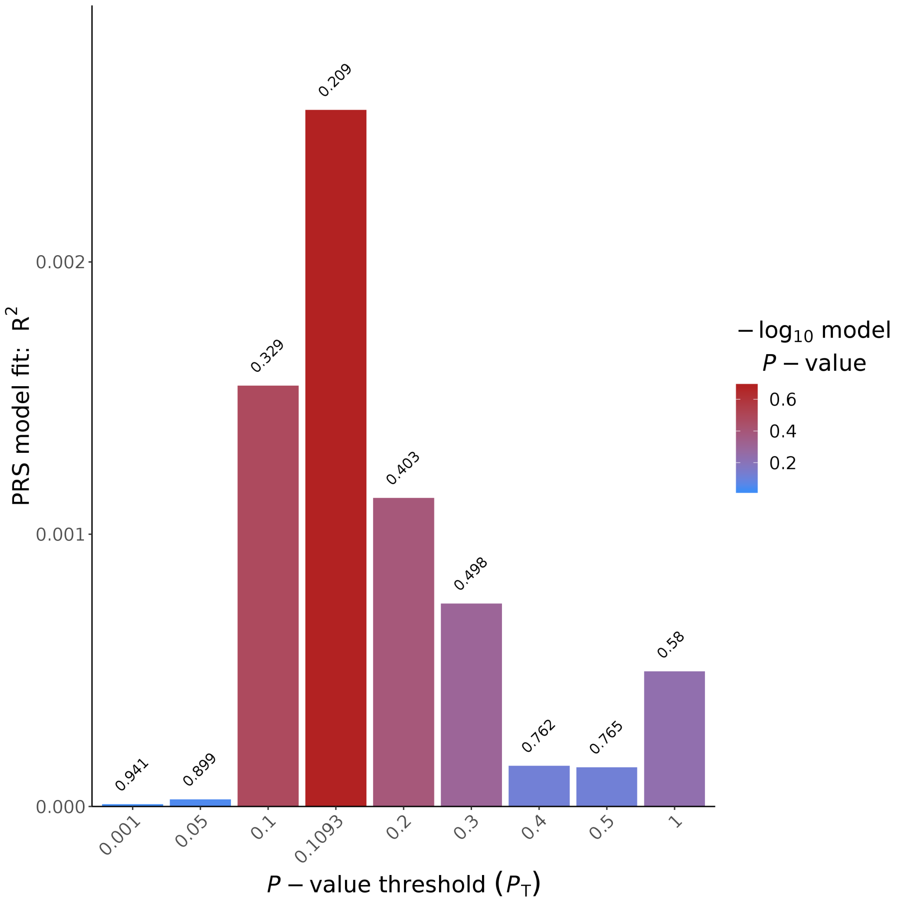


Figure 42: PGS for MMSE (longitudinal) with SNPs from the Jansen GWAS

##### 3.1.2 Attention

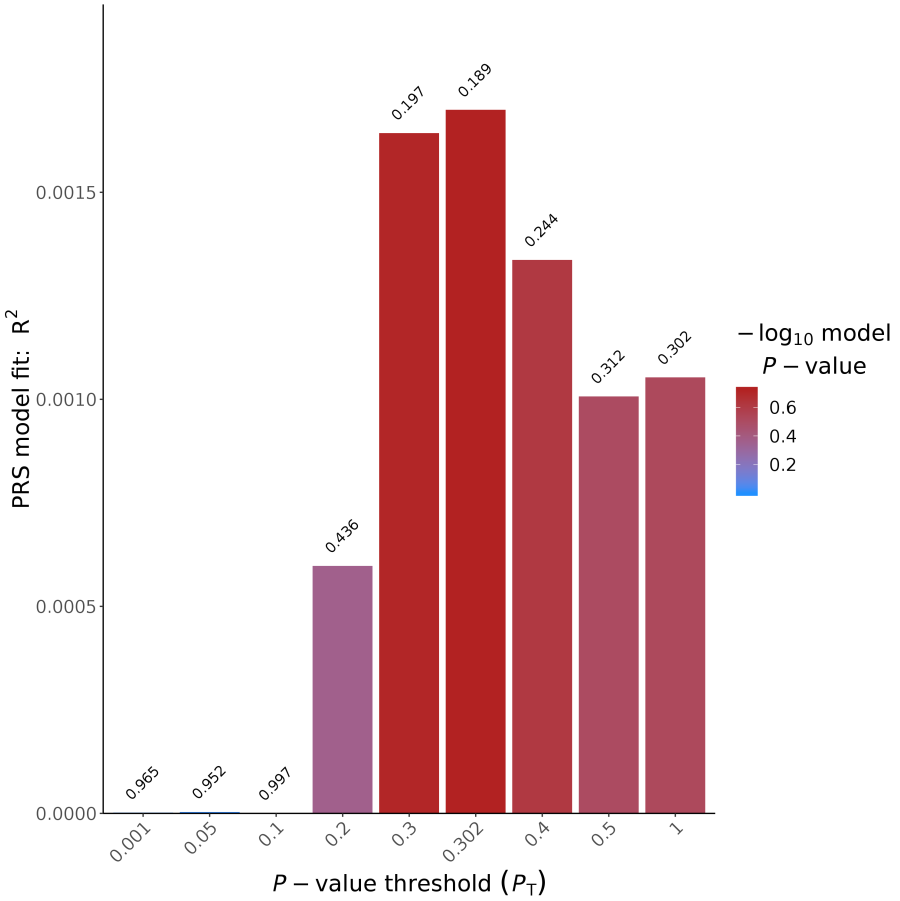


Figure 43: PGS for the cognitive domain attention (cross-sectional) with SNPs from the Jansen GWAS

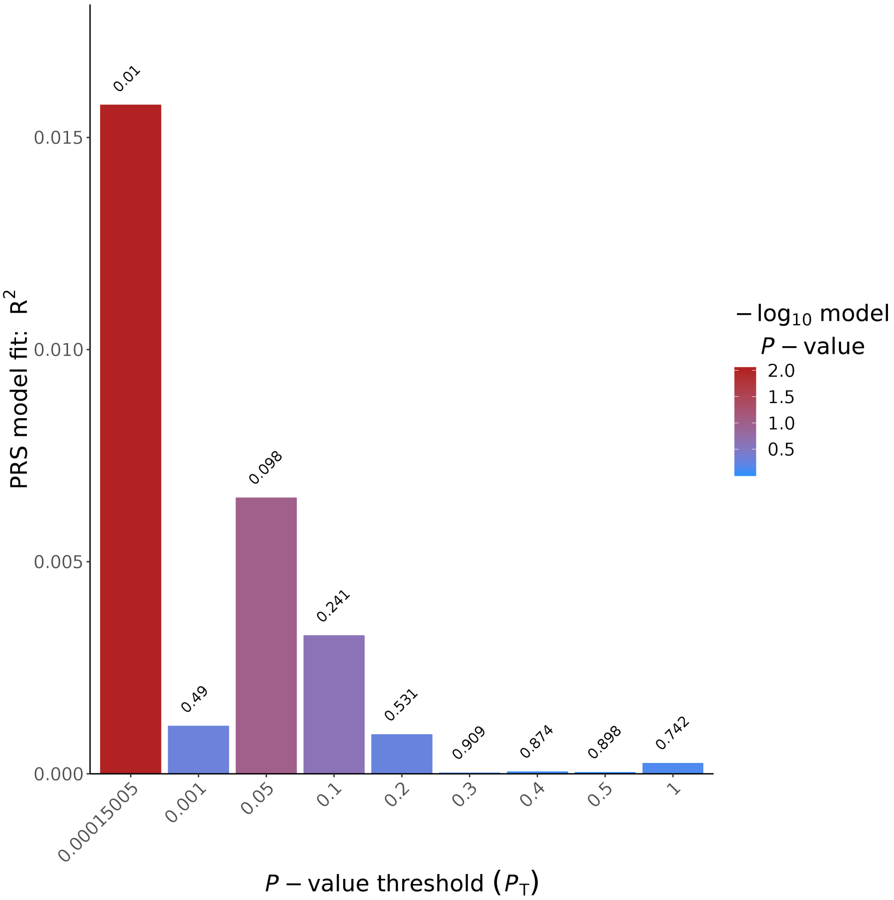


Figure 44: PGS for the cognitive domain attention (longitudinal) with SNPs from the Jansen GWAS

##### 3.1.3 Executive functioning

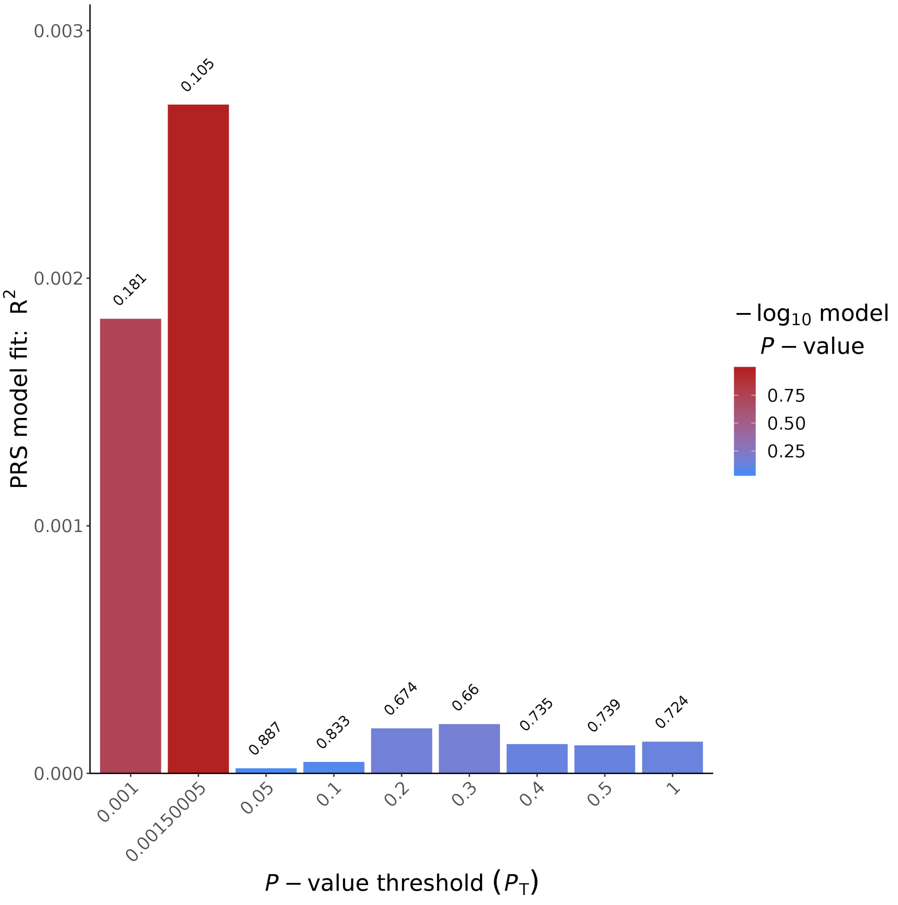


Figure 45: PGS for the cognitive domain executive functioning (cross-sectional) with SNPs from the Jansen GWAS

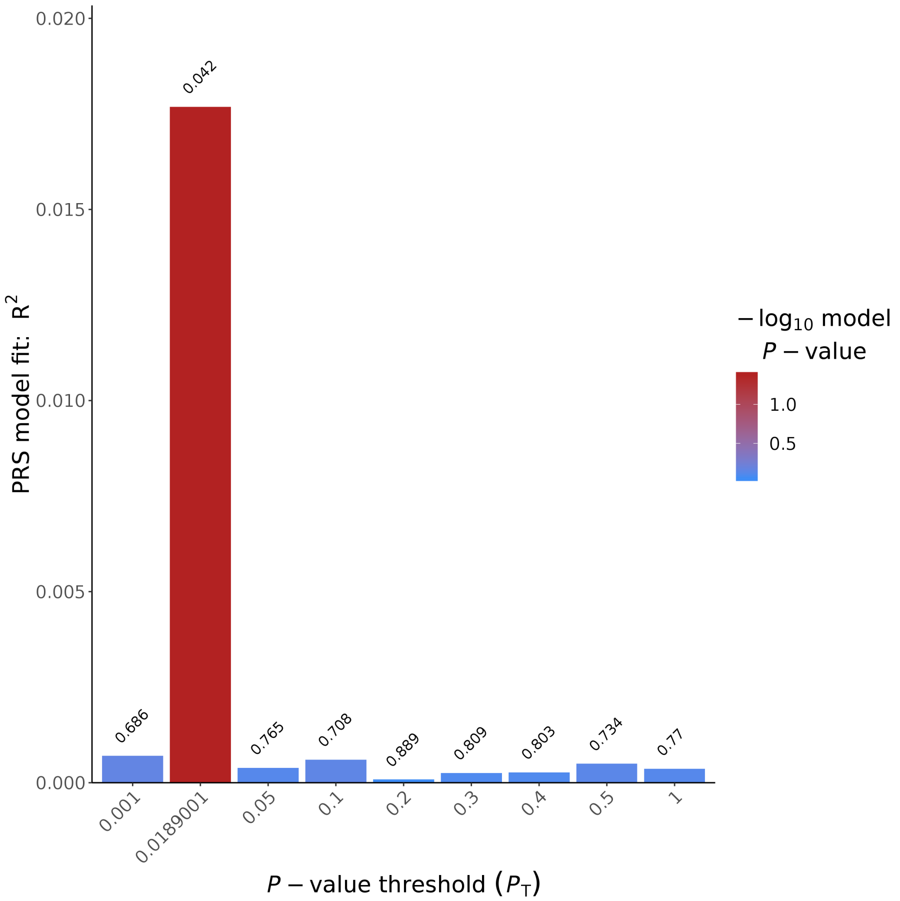


Figure 46: PGS for the cognitive domain executive functioning (longitudinal) with SNPs from the Jansen GWAS

##### 3.1.4 Language

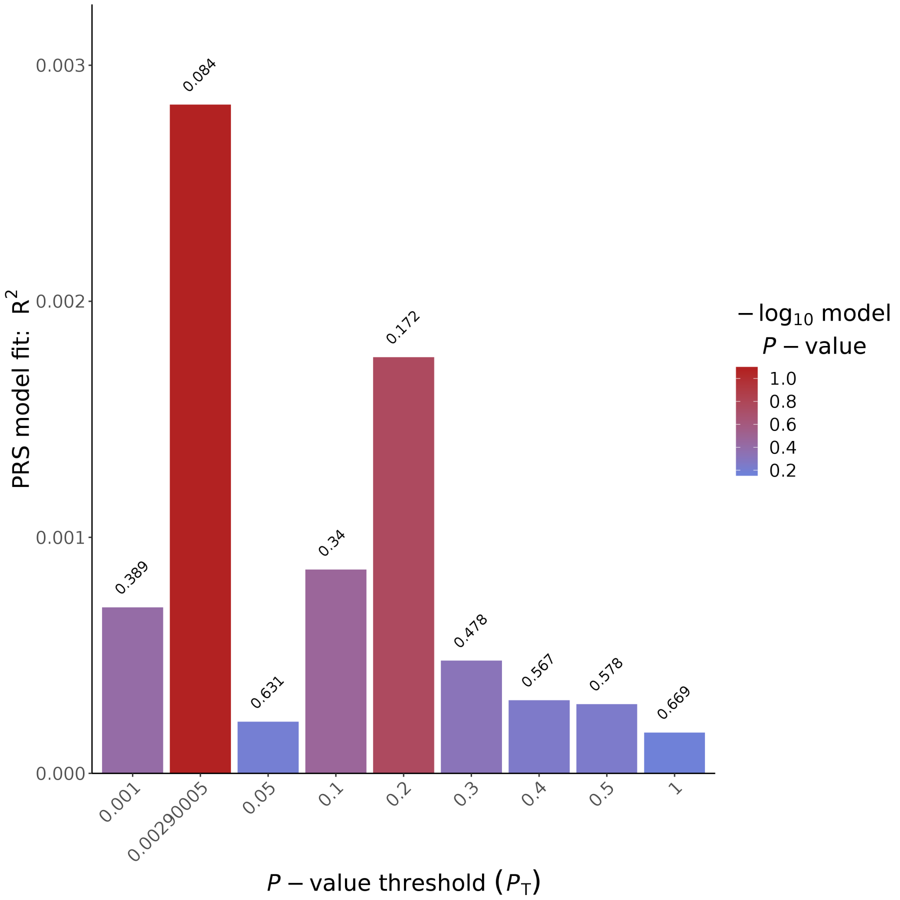


Figure 47: PGS for the cognitive domain language (cross-sectional) with SNPs from the Jansen GWAS

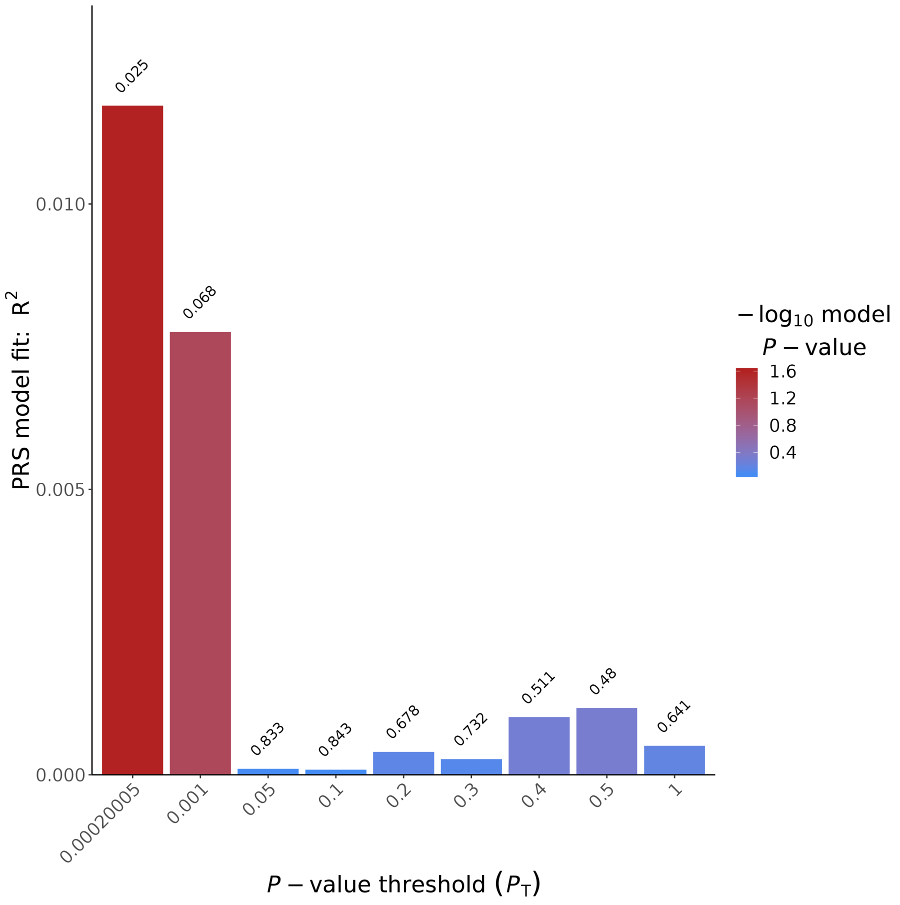


Figure 48: PGS for the cognitive domain language (longitudinal) with SNPs from the Jansen GWAS

##### 3.1.5 Delayed memory

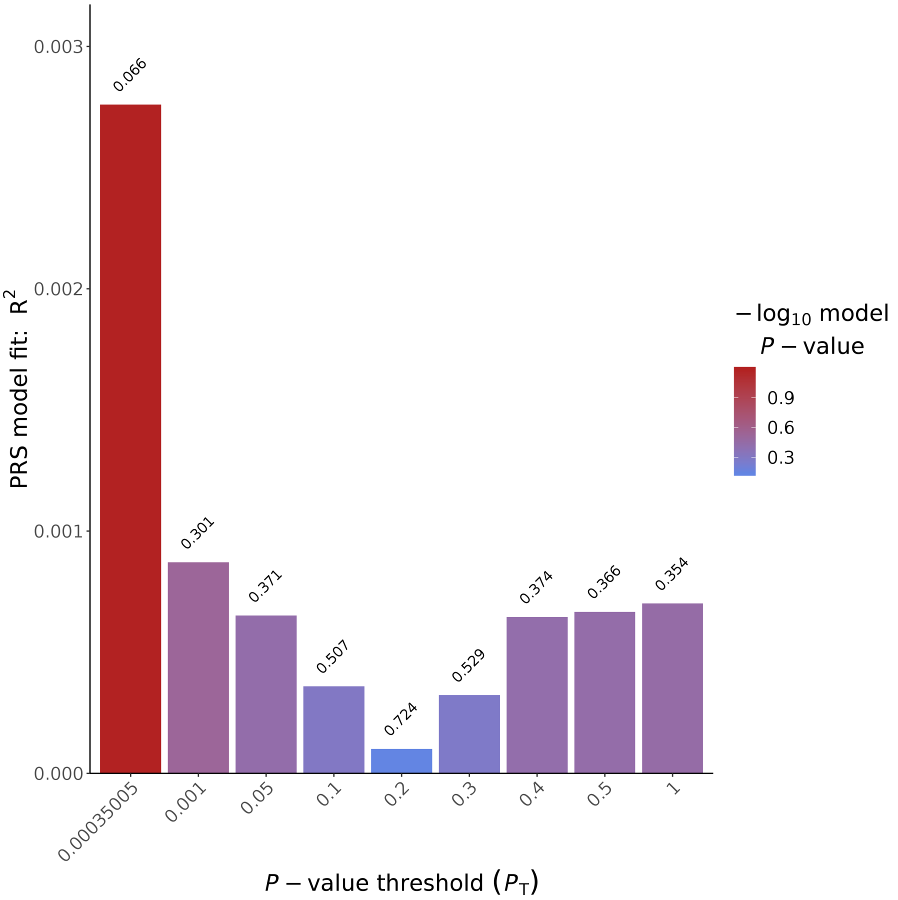


Figure 49: PGS for the cognitive domain delayed memory (cross-sectional) with SNPs from the Jansen GWAS

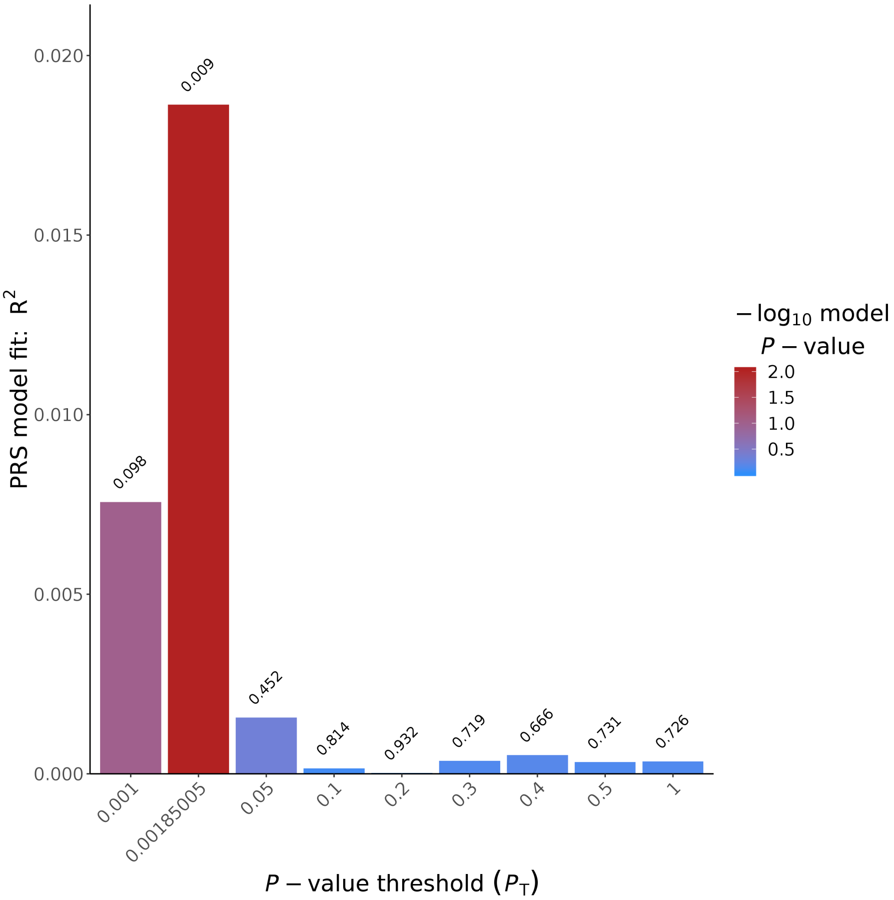


Figure 50: PGS for the cognitive domain delayed memory (longitudinal) with SNPs from the Jansen GWAS

##### 3.1.6 Immediate memory

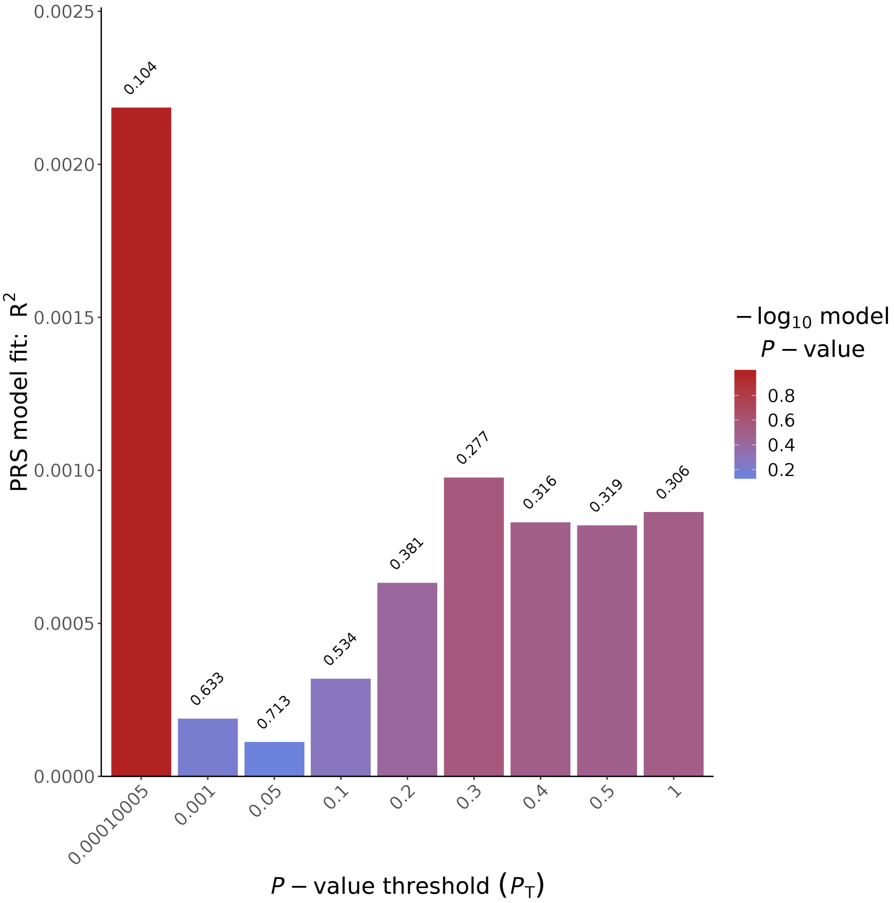


Figure 51: PGS for the cognitive domain immediate memory(cross-sectional) with SNPs from the Jansen GWAS

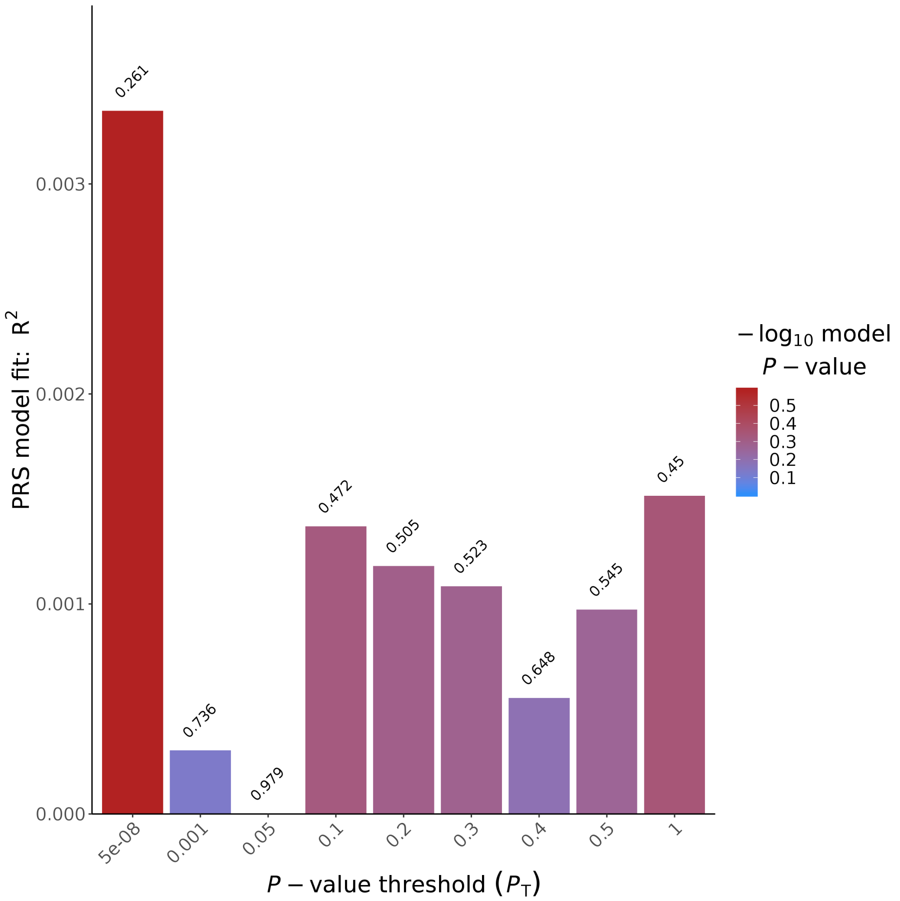


Figure 52: PGS for the cognitive domain immediate memory(longitudinal) with SNPs from the Jansen GWAS

##### 3.1.7 Visuoconstruction

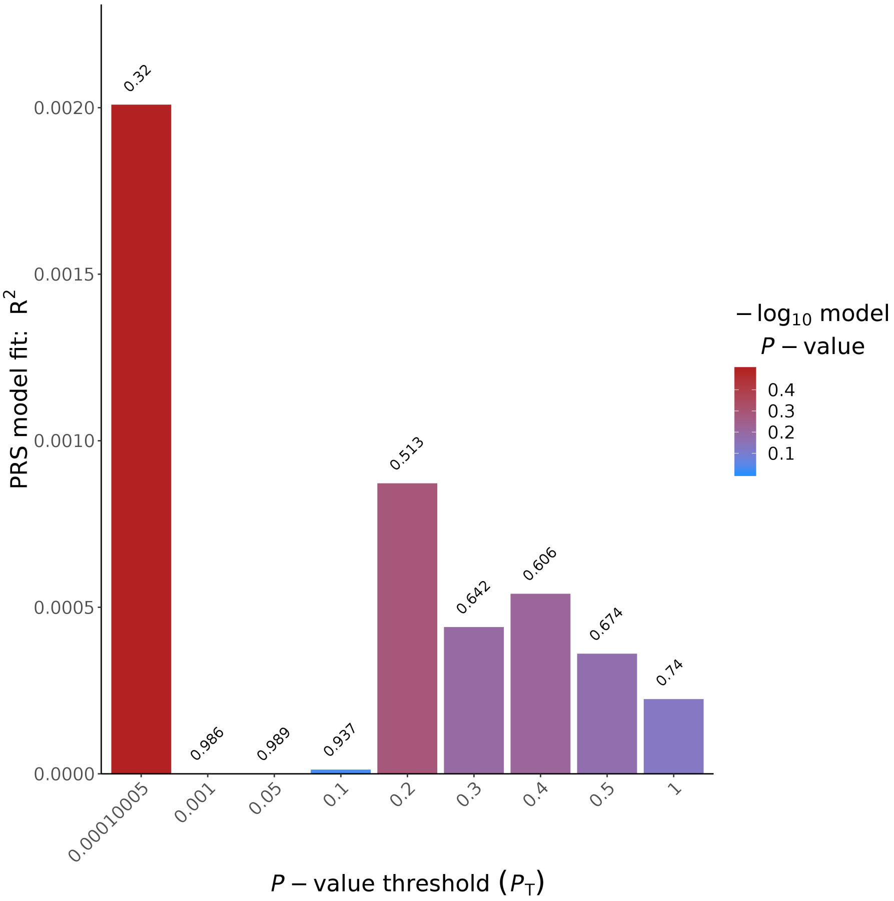


Figure 53: PGS for the cognitive domain visuoconstruction (cross-sectional) with SNPs from the Jansen GWAS

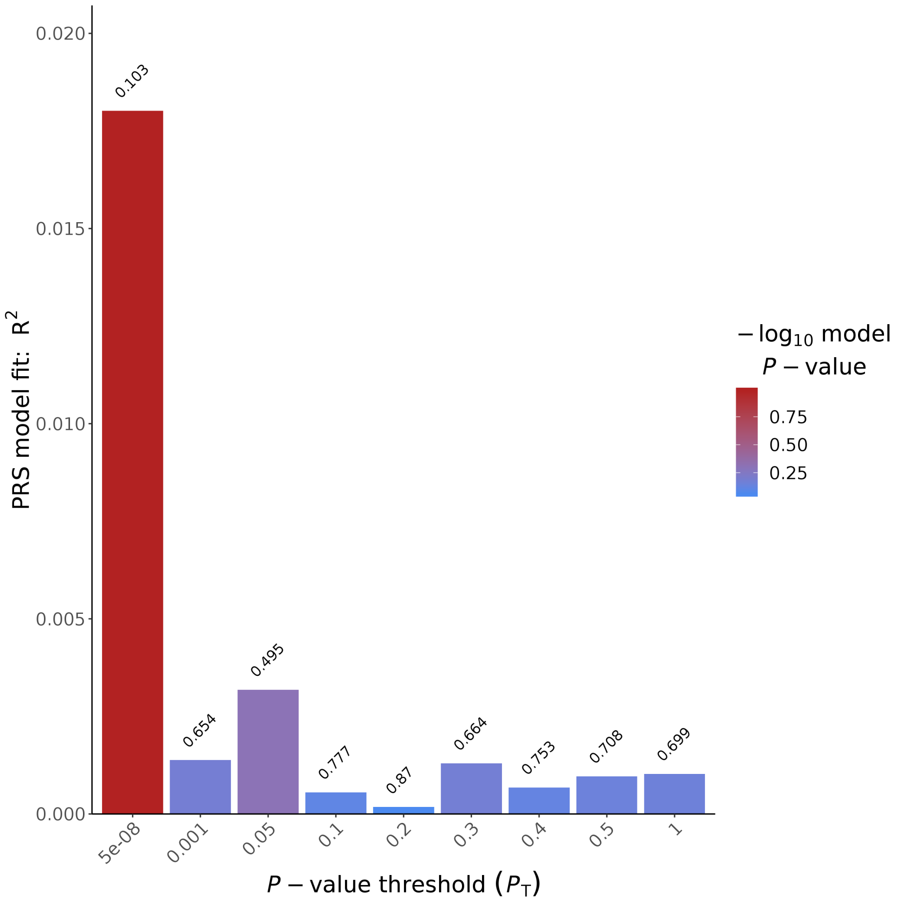


Figure 54: PGS for the cognitive domain visuoconstruction (longitudinal) with SNPs from the Jansen GWAS

#### 3.2 PGS with Davies GWAS

##### 3.2.1 MMSE

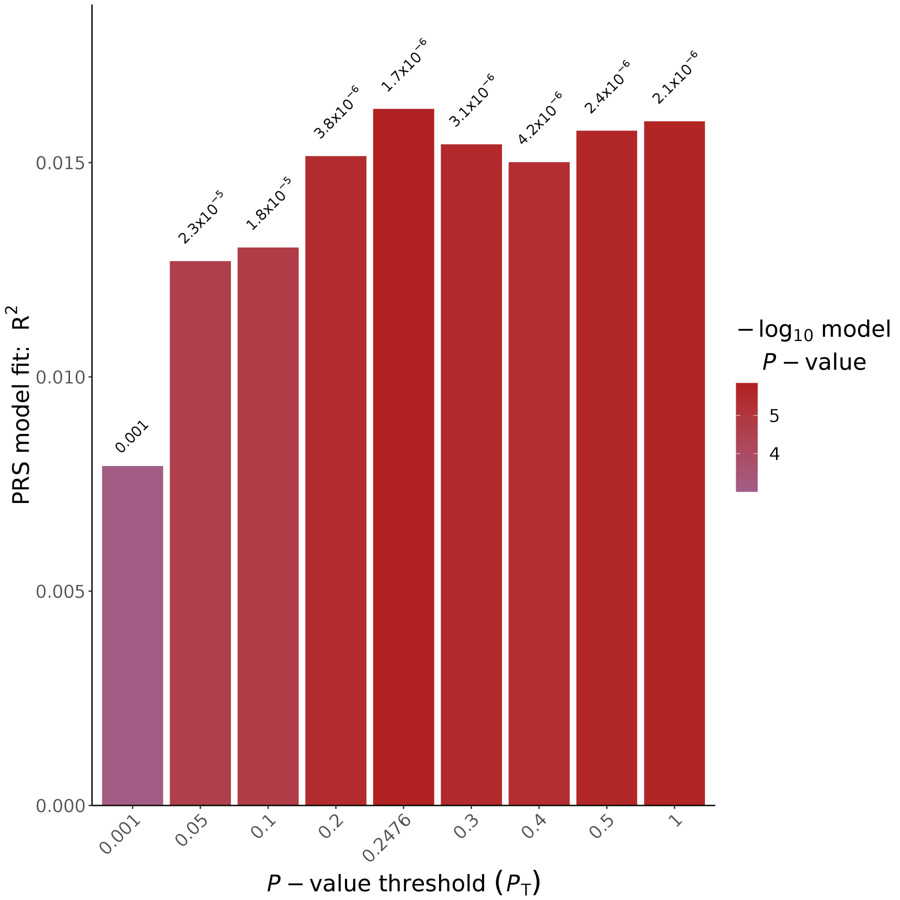


Figure 55: PGS for den Test MMSE (cross-sectional) with SNPs from the Davies GWAS

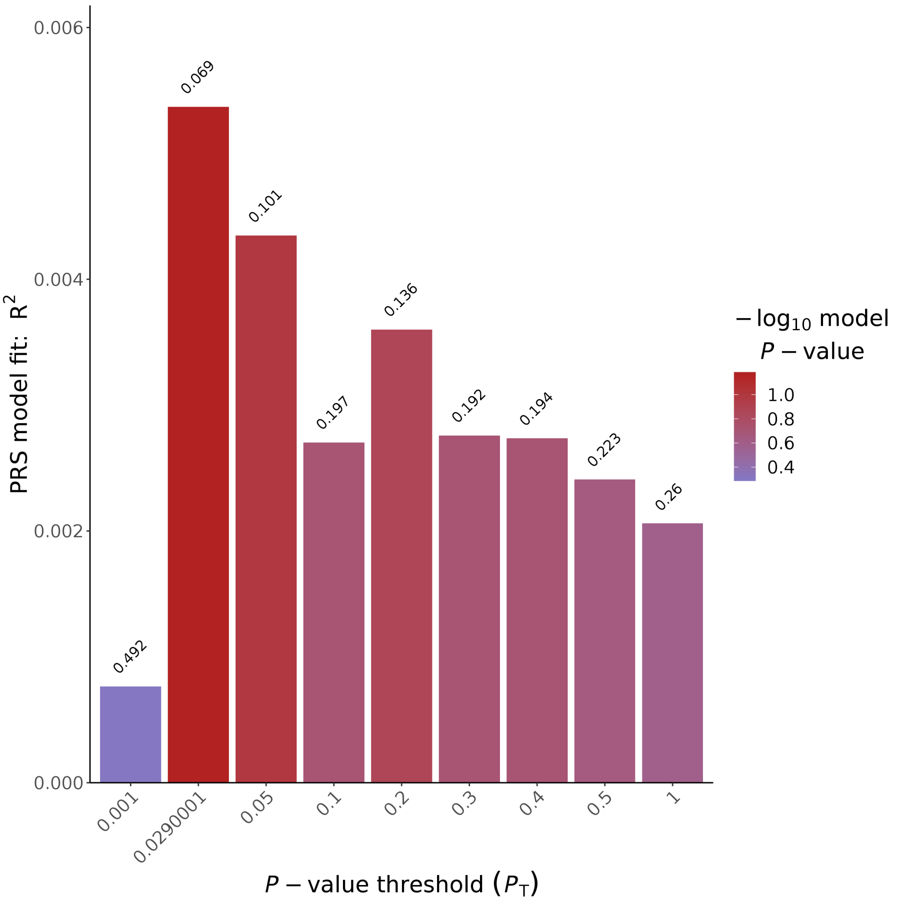


Figure 56: PGS for den Test MMSE (longitudinal) with SNPs from the Davies GWAS

##### 3.2.2 Attention

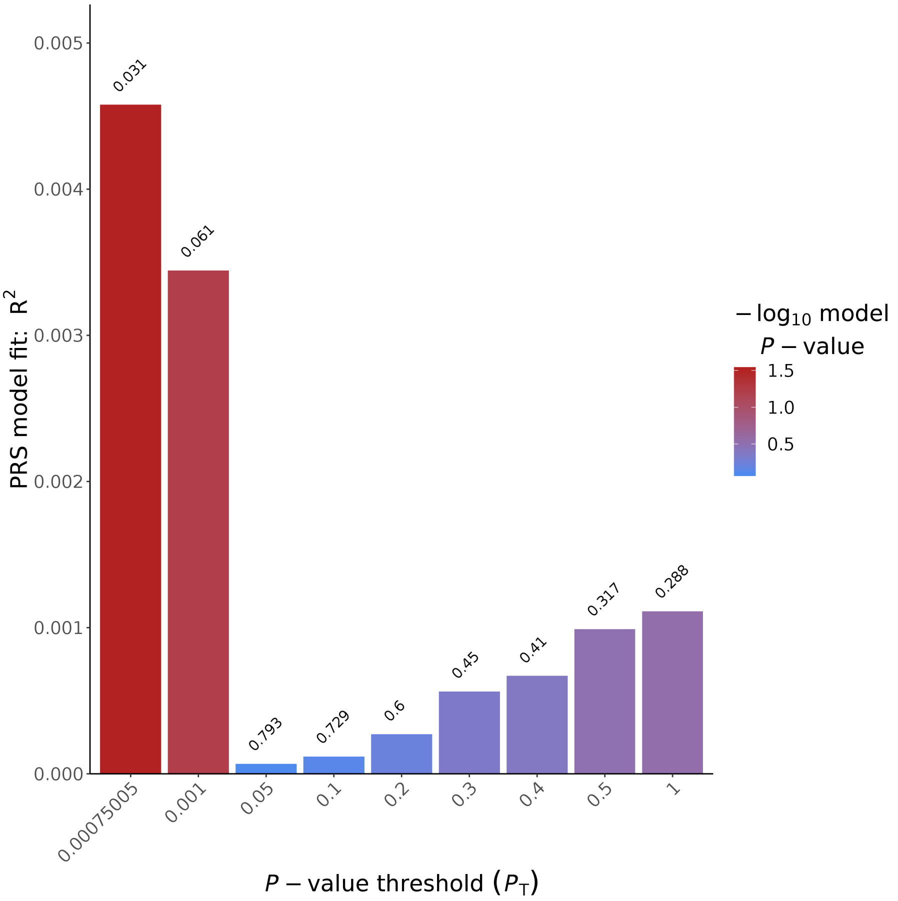


Figure 57: PGS for the cognitive domain attention (cross-sectional) with SNPs from the Davies GWAS

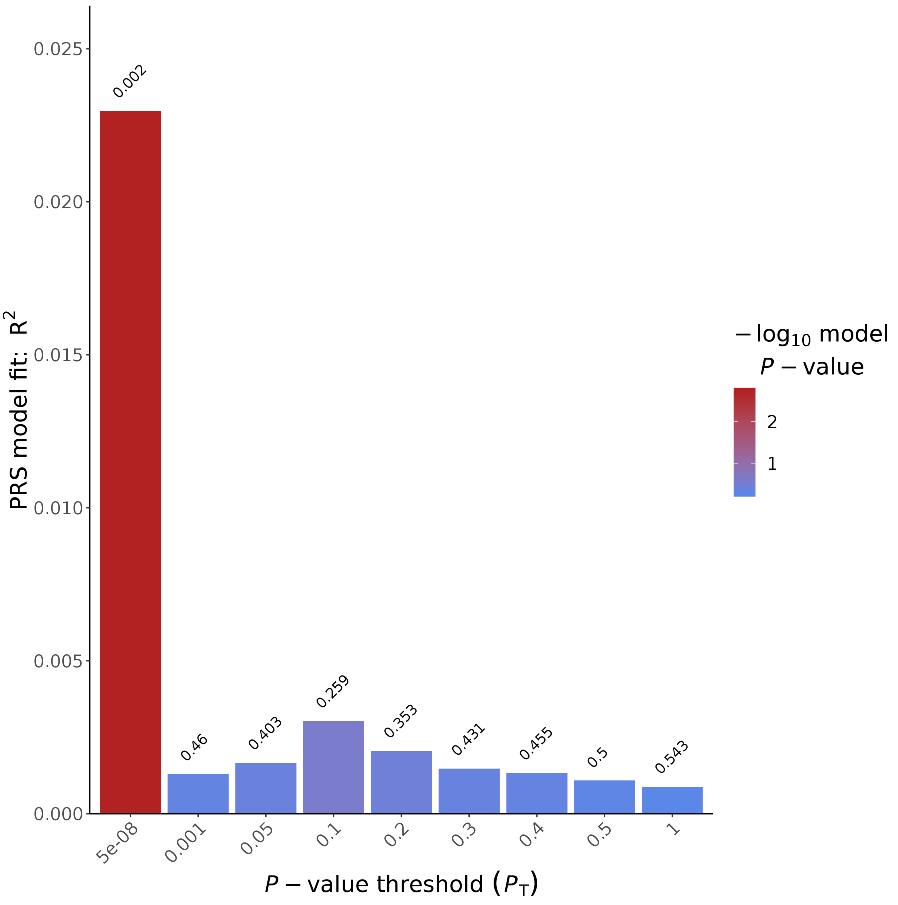


Figure 58: PGS for the cognitive domain attention (longitudinal) with SNPs from the Davies GWAS

##### 3.2.3 Executive functioning

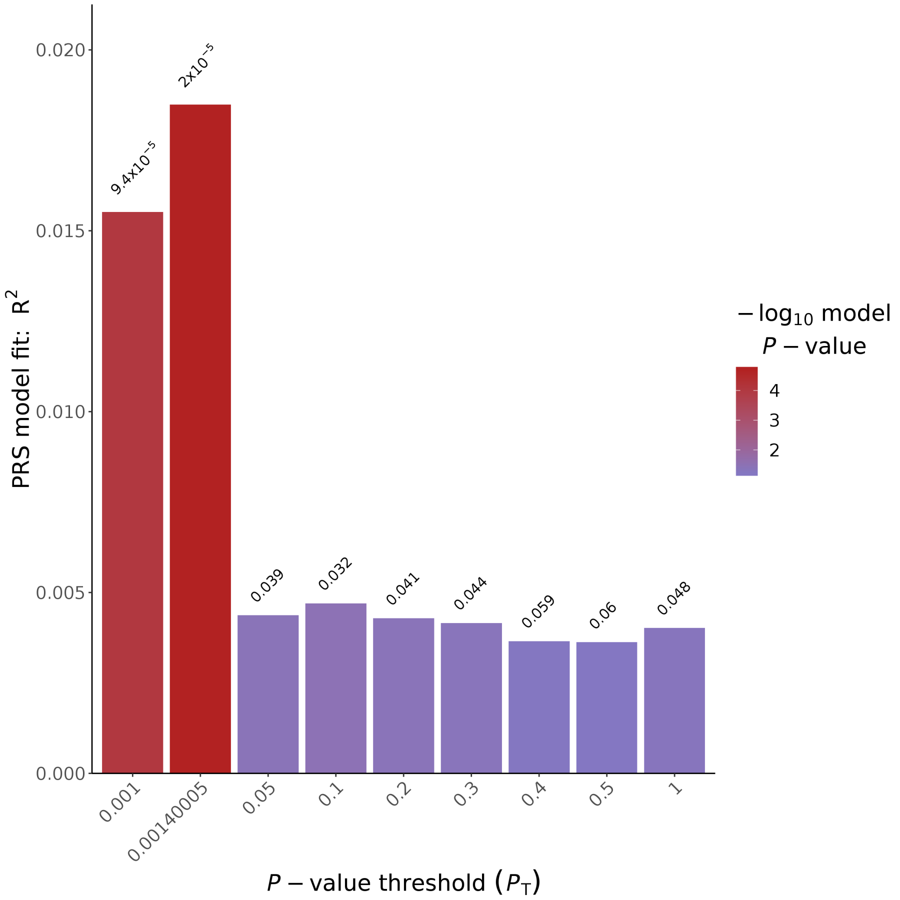


Figure 59: PGS for the cognitive domain executive functioning (cross-sectional) with SNPs from the Davies GWAS

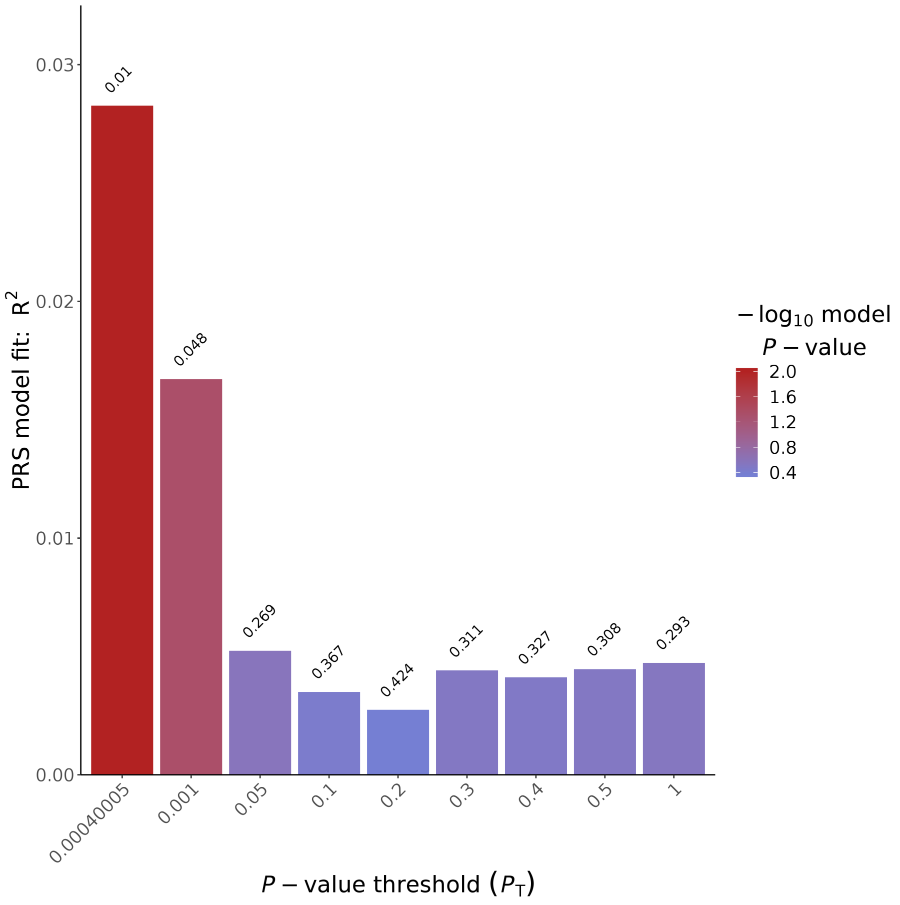


Figure 60: PGS for the cognitive domain executive functioning (longitudinal) with SNPs from the Davies GWAS

##### 3.2.4 Language

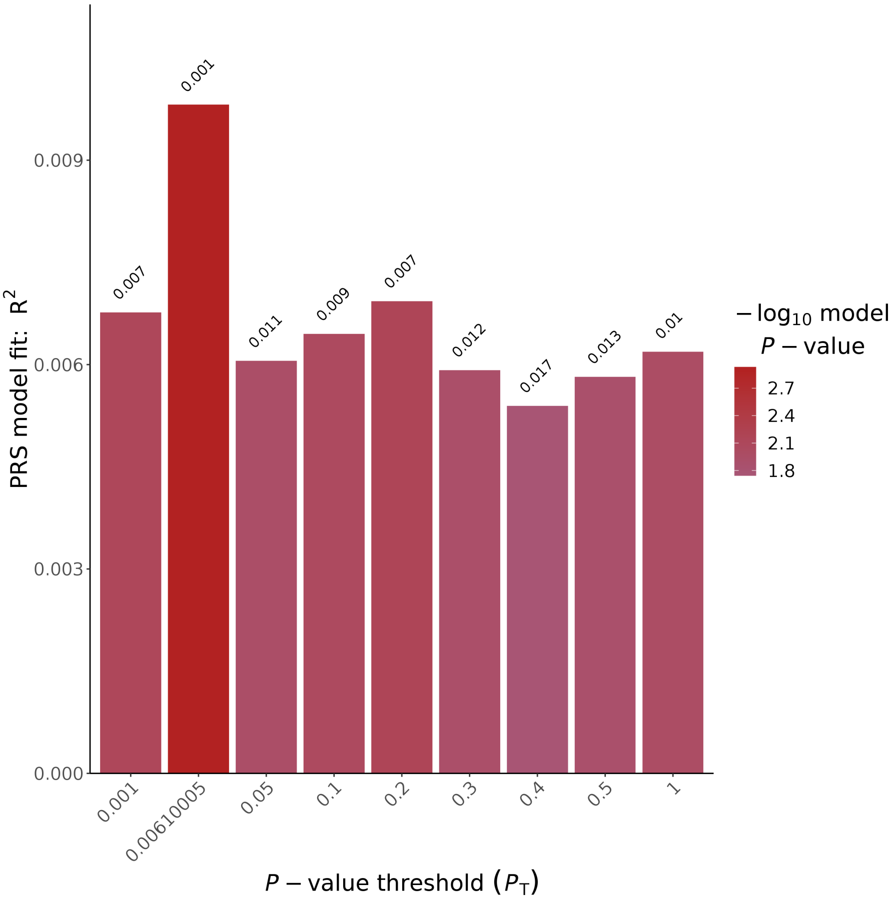


Figure 61: PGS for the cognitive domain language (cross-sectional) with SNPs from the Davies GWAS

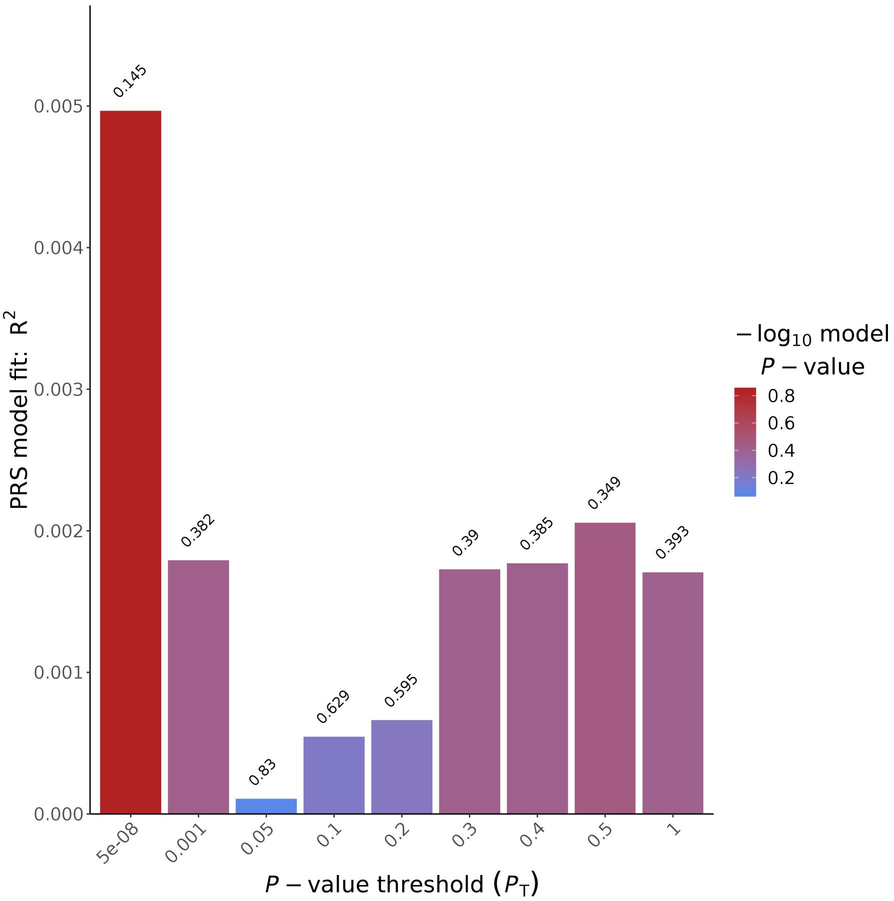


Figure 62: PGS for the cognitive domain language (longitudinal) with SNPs from the Davies GWAS

##### 3.2.5 Delayed memory

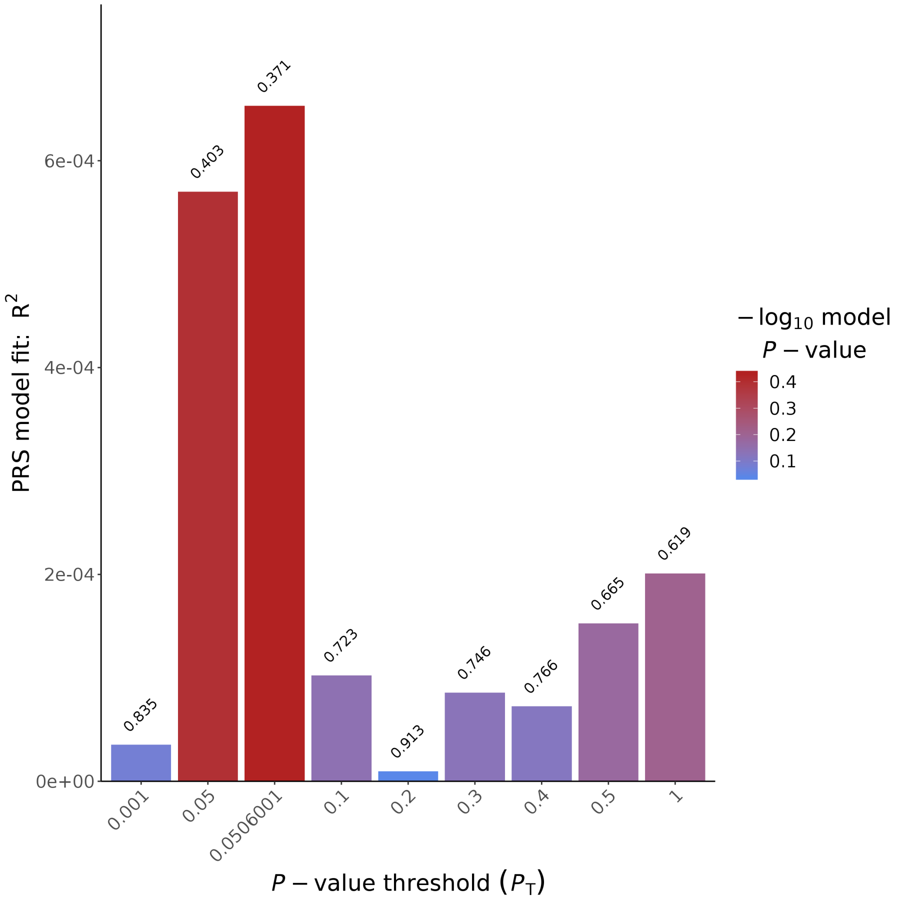


Figure 63: PGS for the cognitive domain delayed memory (cross-sectional) with SNPs from the Davies GWAS

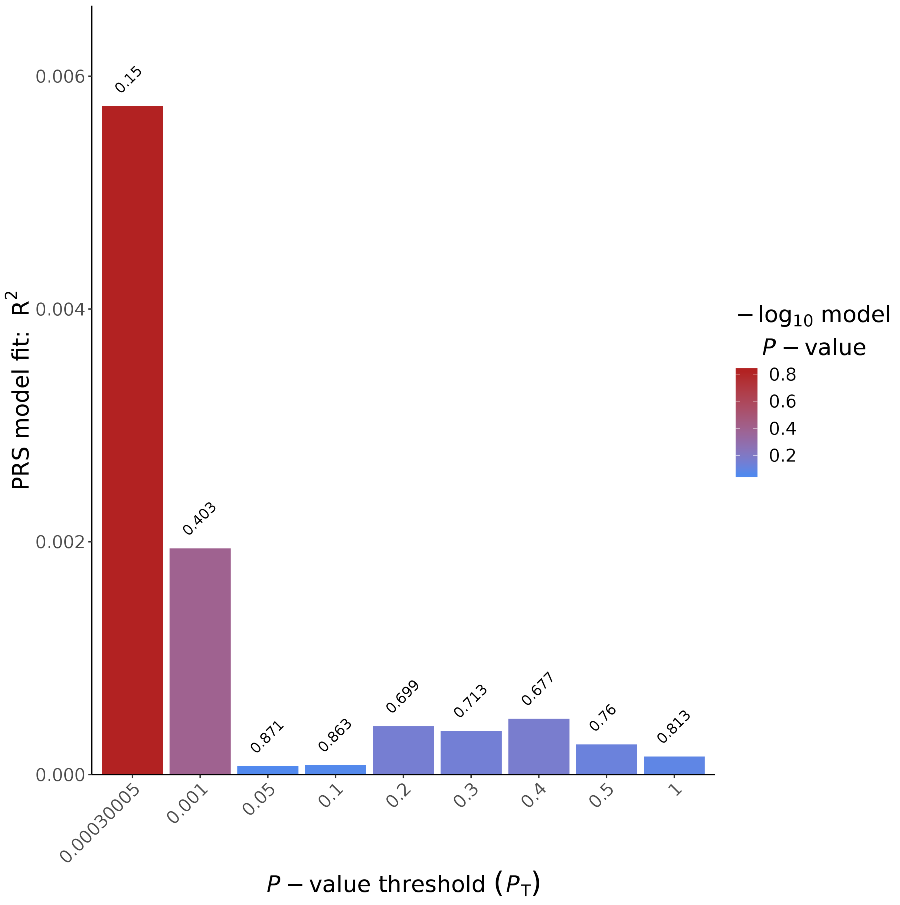


Figure 64: PGS for the cognitive domain delayed memory (longitudinal) with SNPs from the Davies GWAS

##### 3.2.6 Immediate memory

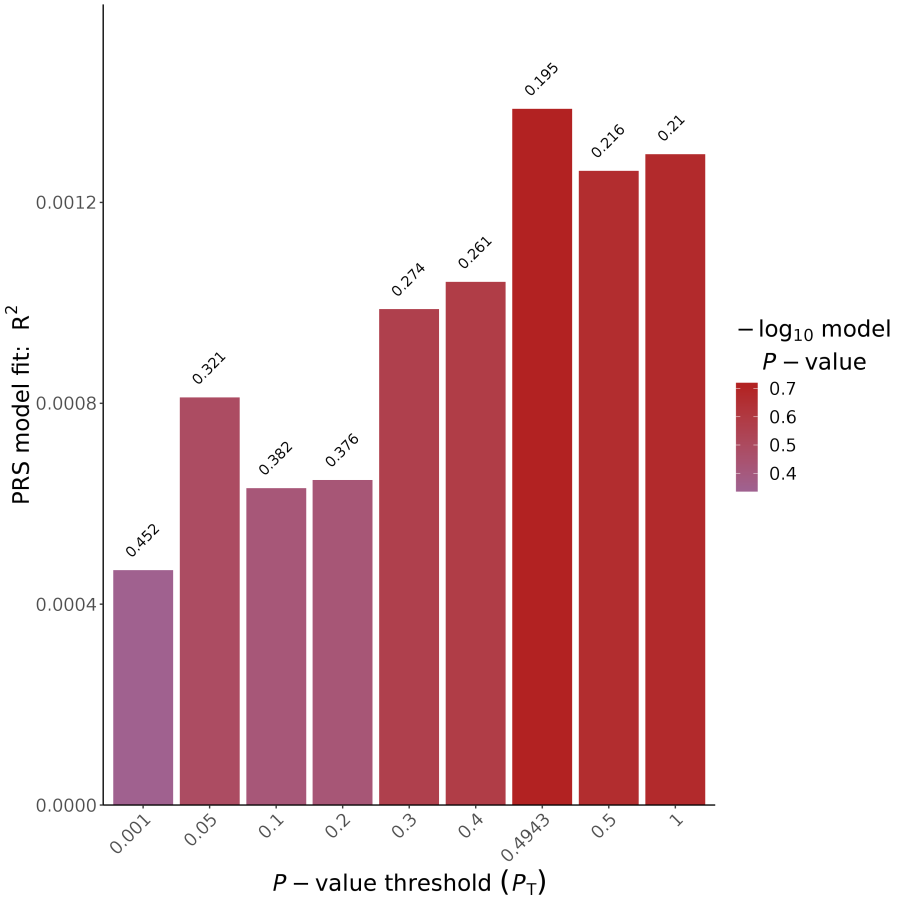


Figure 65: PGS for the cognitive domain immediate (cross-sectional) with SNPs from the Davies GWAS

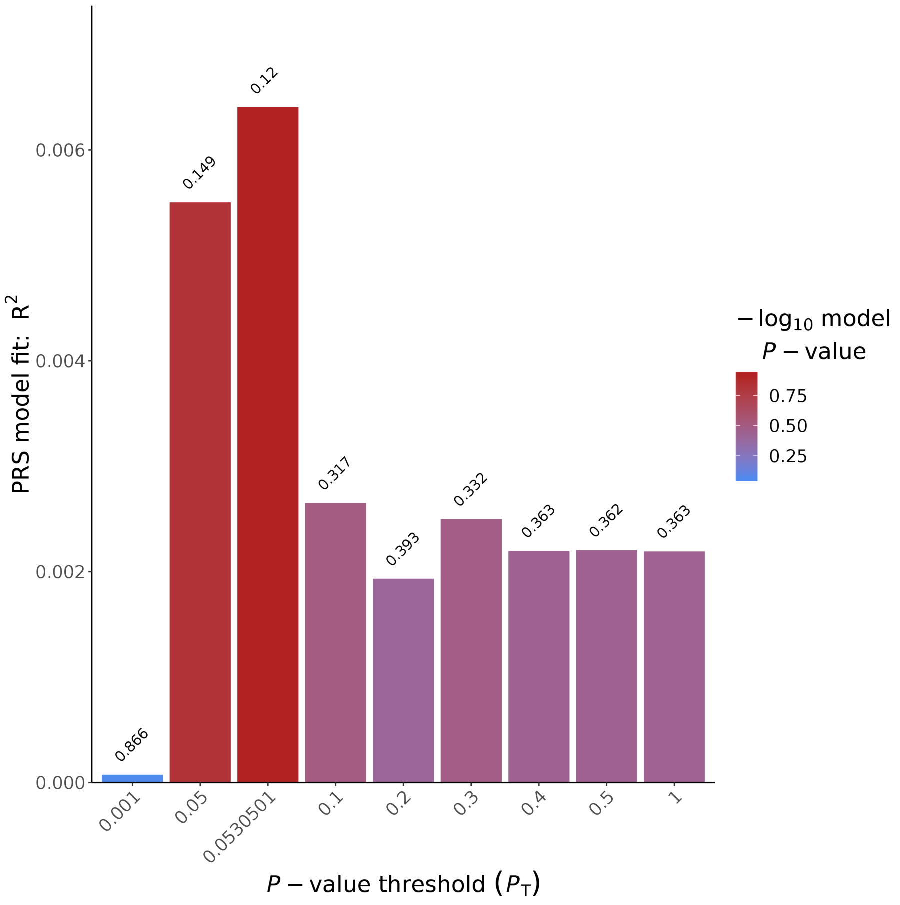


Figure 66: PGS for the cognitive domain immediate memory (longitudinal) with SNPs from the Davies GWAS

##### 3.2.7 Visuoconstruction

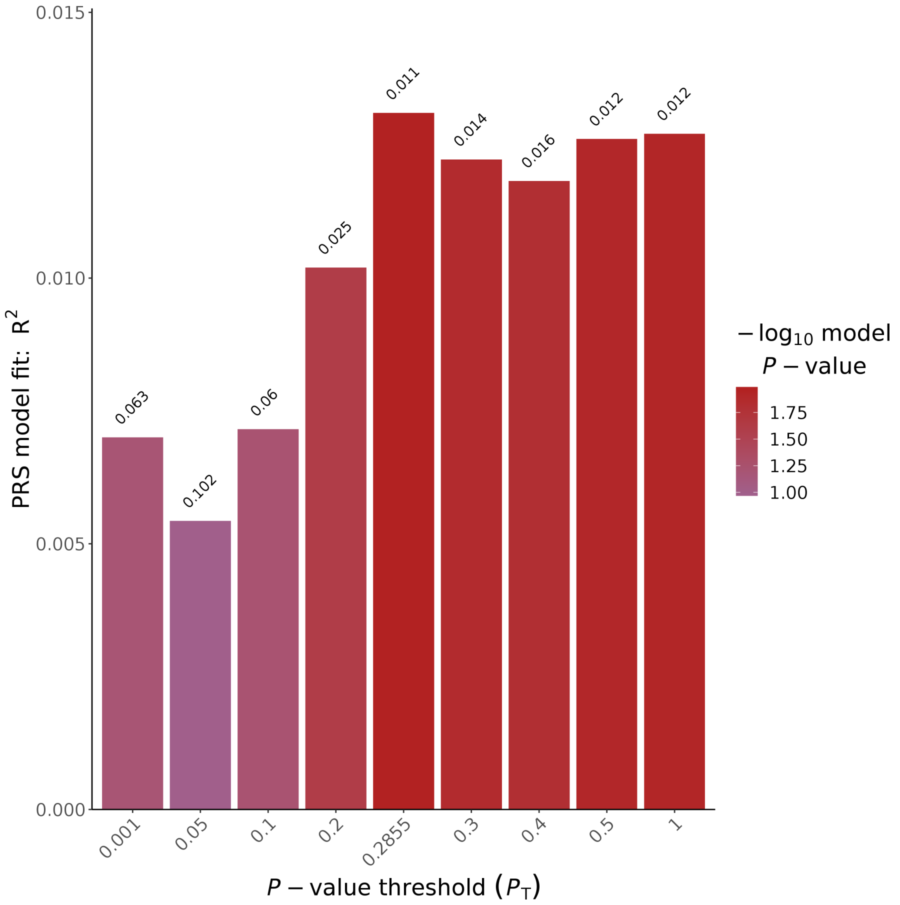


Figure 67: PGS for the cognitive domain visuoconstruction (cross-sectional) with SNPs from the Davies GWAS

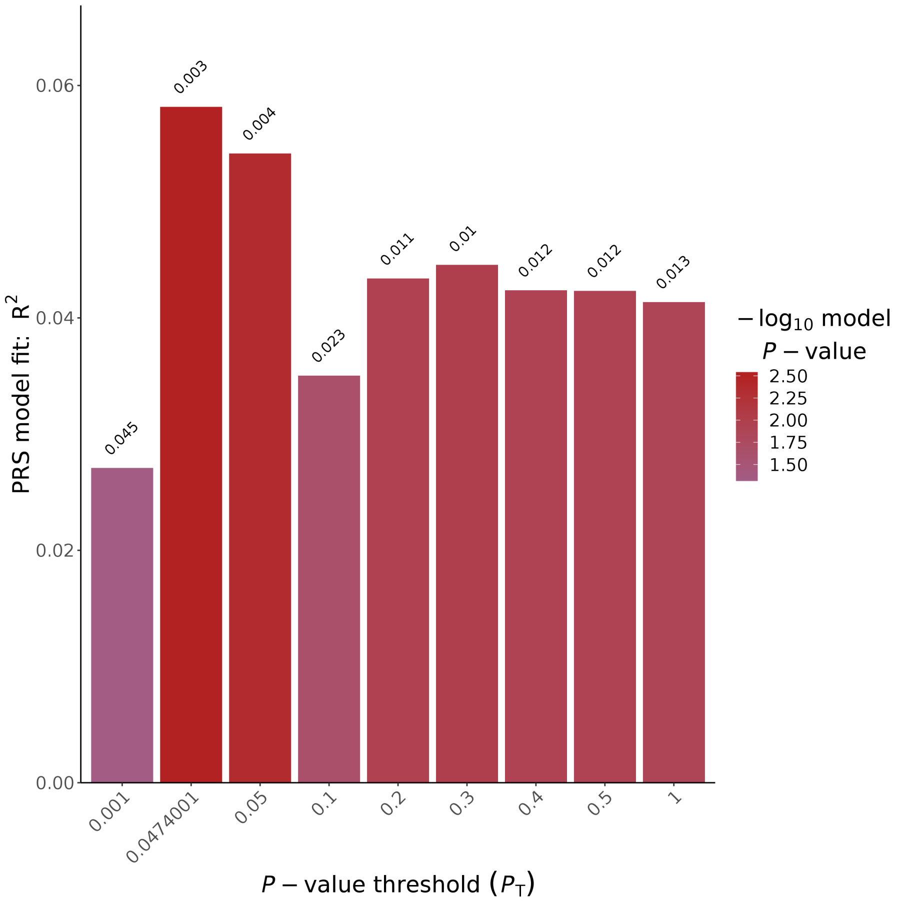


Figure 68: PGS for the cognitive domain visuoconstruction (longitudinal) with SNPs from the Davies GWAS

### 4 PGS - MRI phenotypes

#### 4.1 PGS with Jansen GWAS

##### 4.1.1 Fazekas

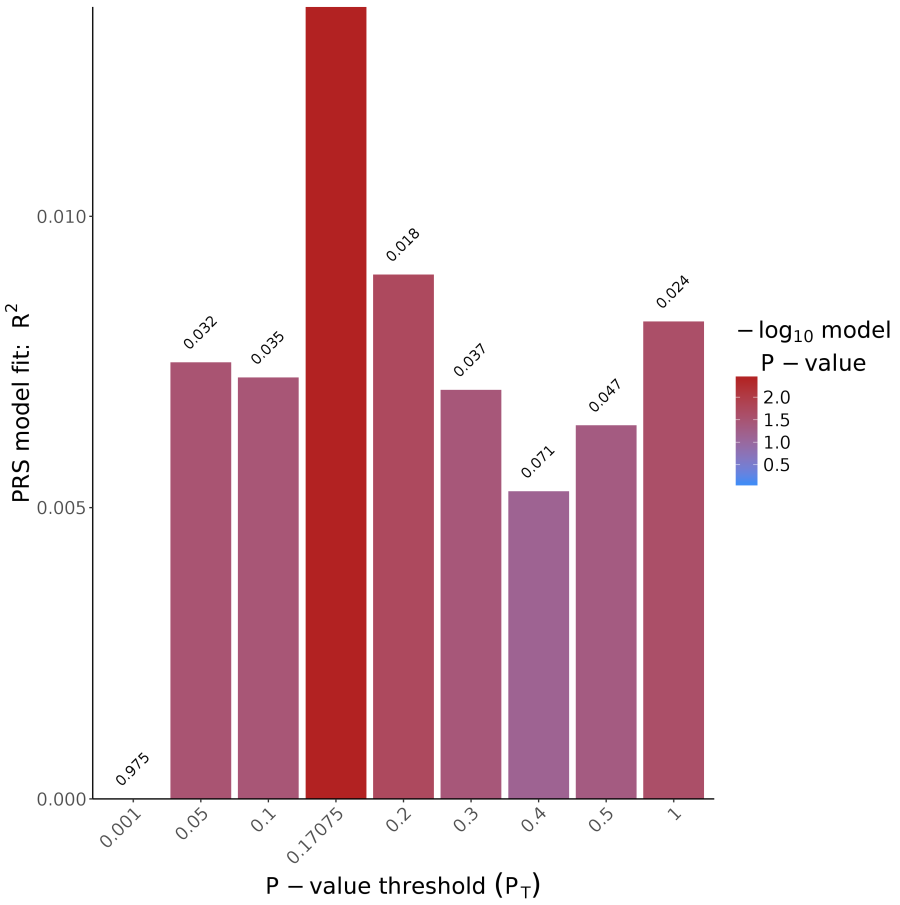


Figure 69: PGS for the MRI phenotype Fazekas with SNPs from the Jansen GWAS

##### 4.1.2 Hippocampus volume sum

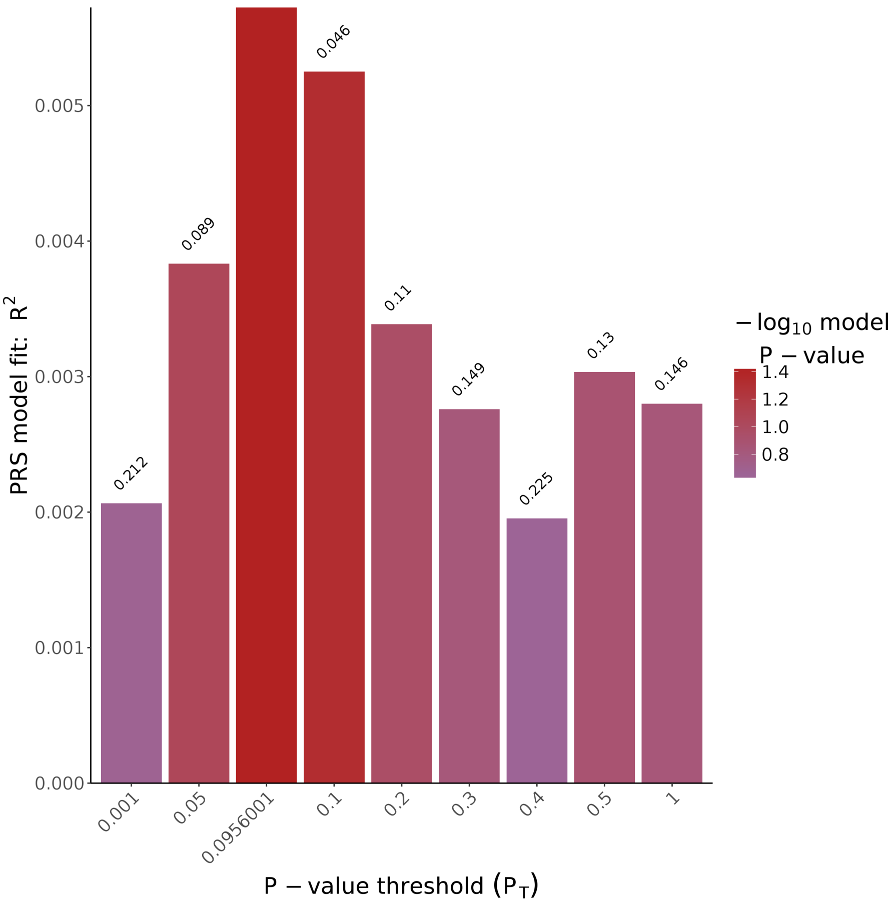


Figure 70: PGS for the MRI phenotype hippocampus volume sum with SNPs from the Jansen GWAS

##### 4.1.3 Hippocampus volume left

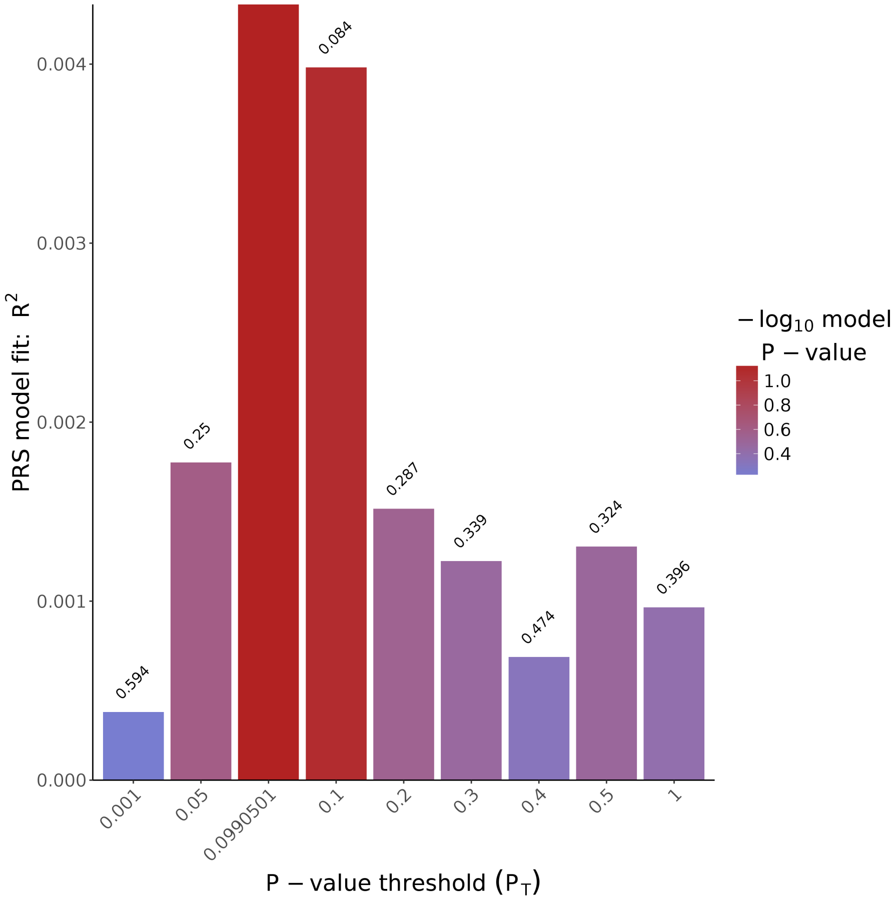


Figure 71: PGS for the MRI phenotype hippocampus volume left with SNPs from the Jansen GWAS

##### 4.1.4 Hippocampus volume right

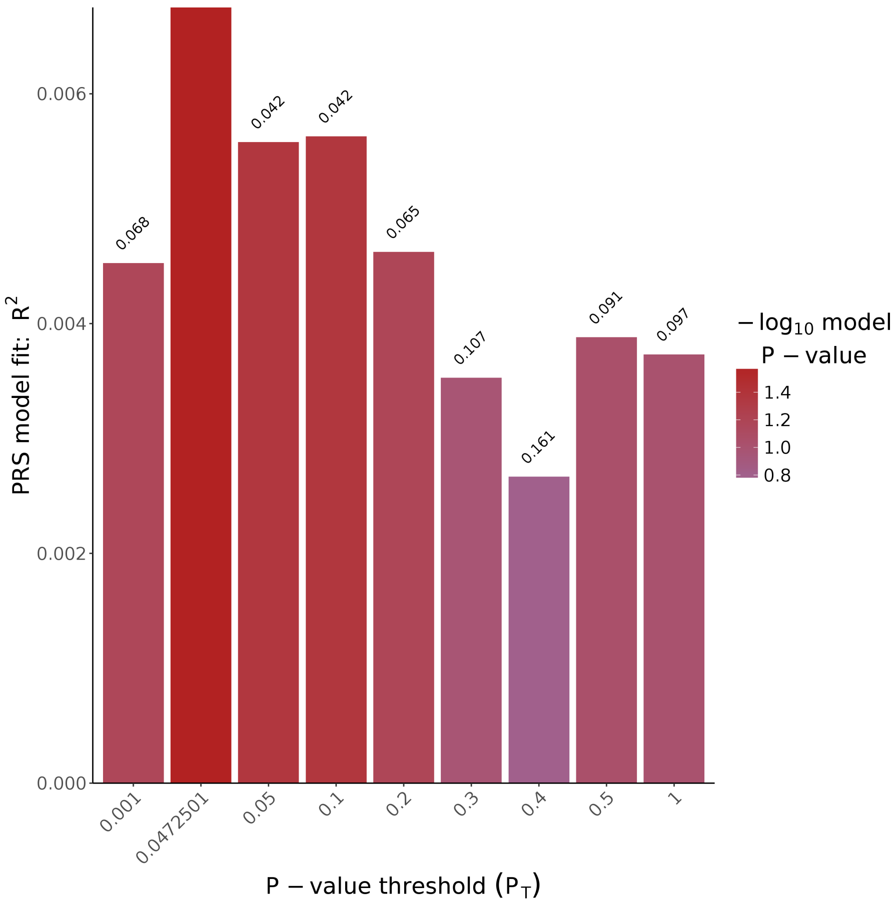


Figure 72: PGS for the MRI phenotype hippocampus volume right with SNPs from the Jansen GWAS

##### 4.1.5 Thickness

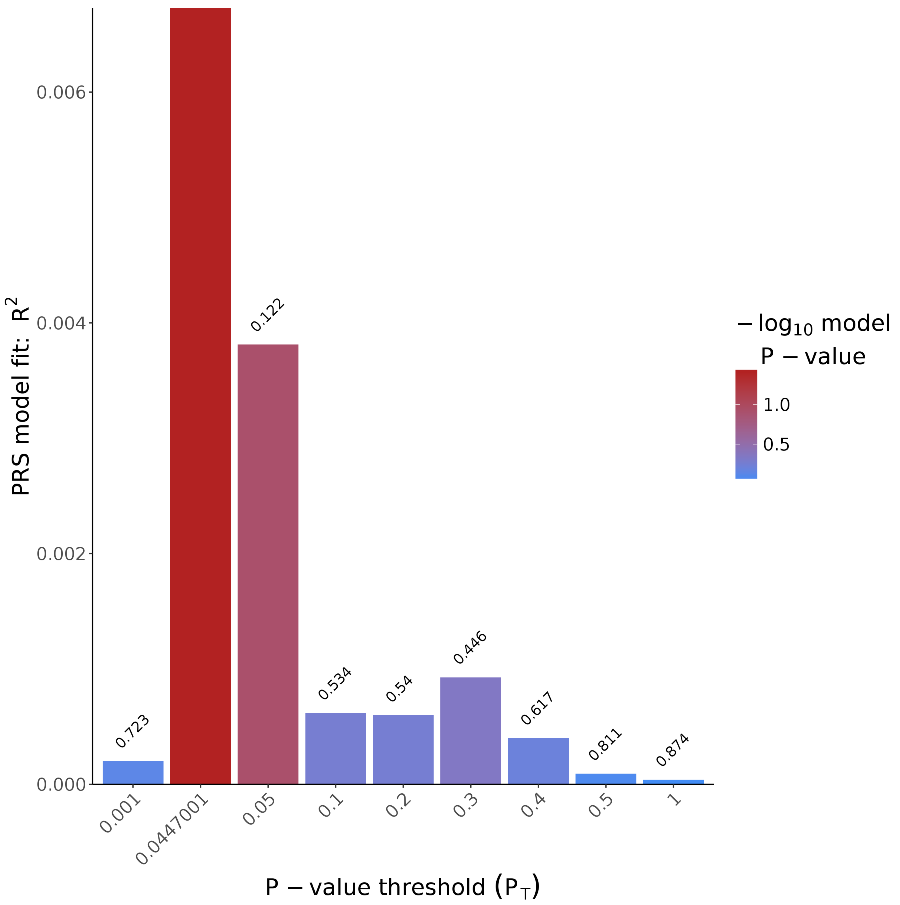


Figure 73: PGS for the MRI phenotype thickness with SNPs from the Jansen GWAS

#### 4.2 PGS with Hibar GWAS

##### 4.2.1 Fazekas

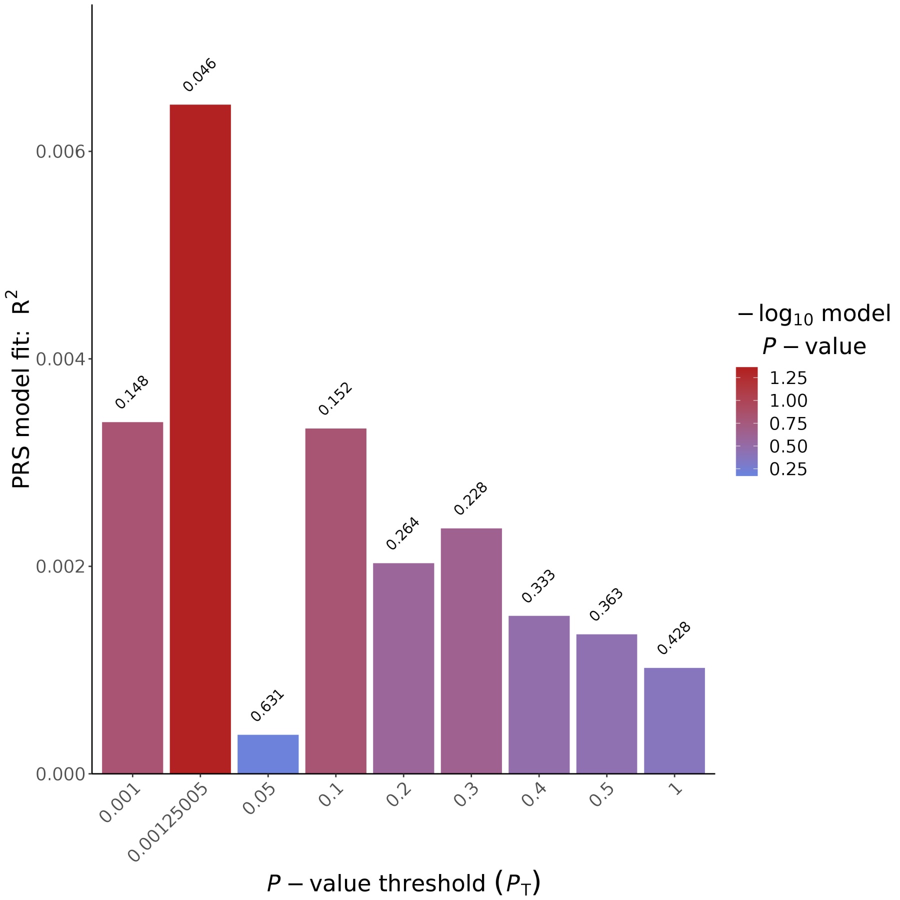


Figure 74: PGS for the MRI phenotype Fazekas with SNPs from the Hibar GWAS

##### 4.2.2 Hippocampus volume sum

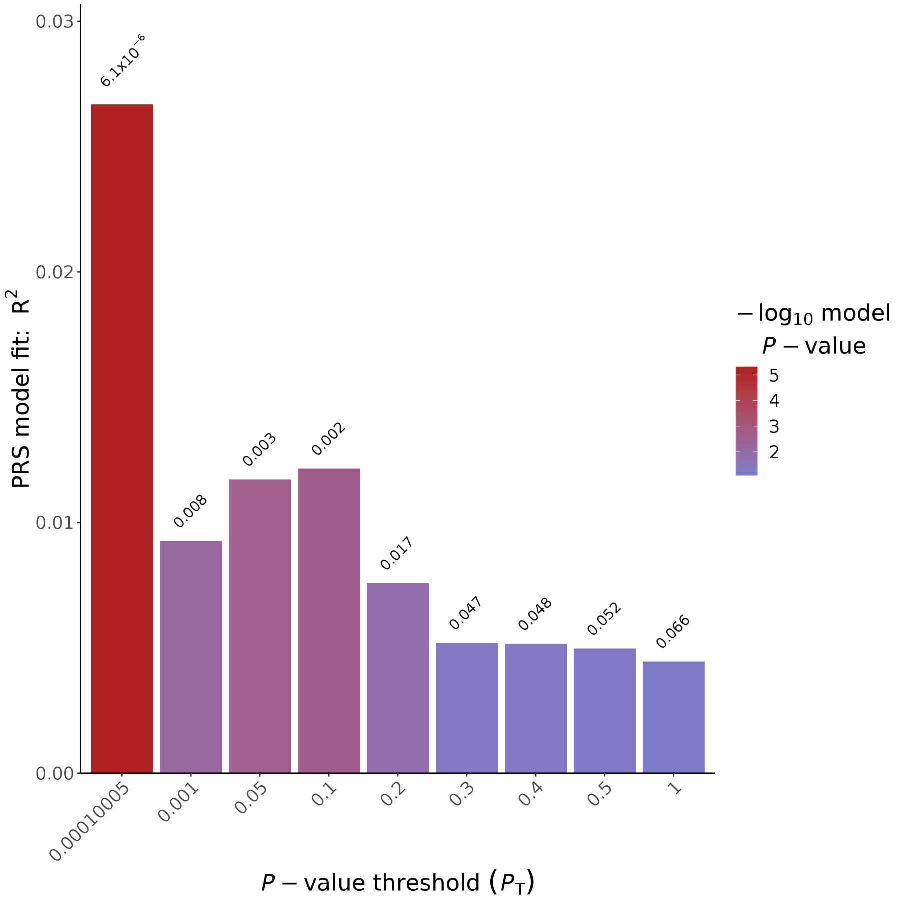


Figure 75: PGS for the MRI phenotype hippocampus volume sum with SNPs from the Hibar GWAS

##### 4.2.3 Hippocampus volume left

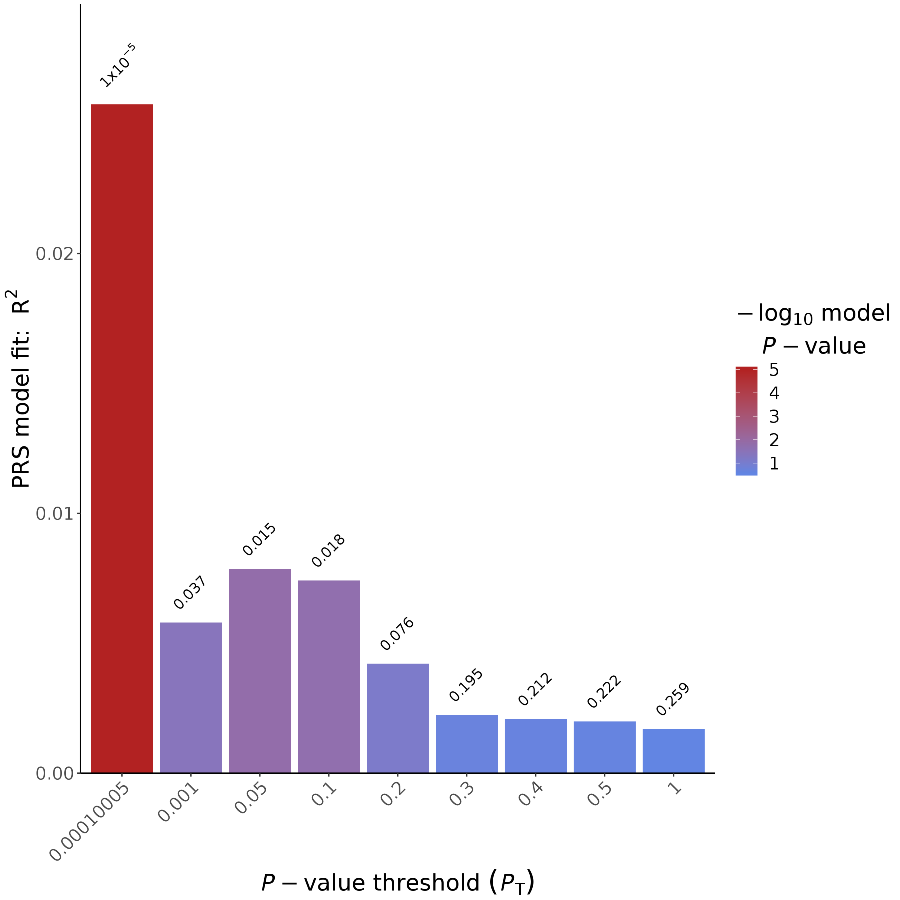


Figure 76: PGS for the MRI phenotype hippocampus volume left with SNPs from the Hibar GWAS

##### 4.2.4 Hippocampus volume right

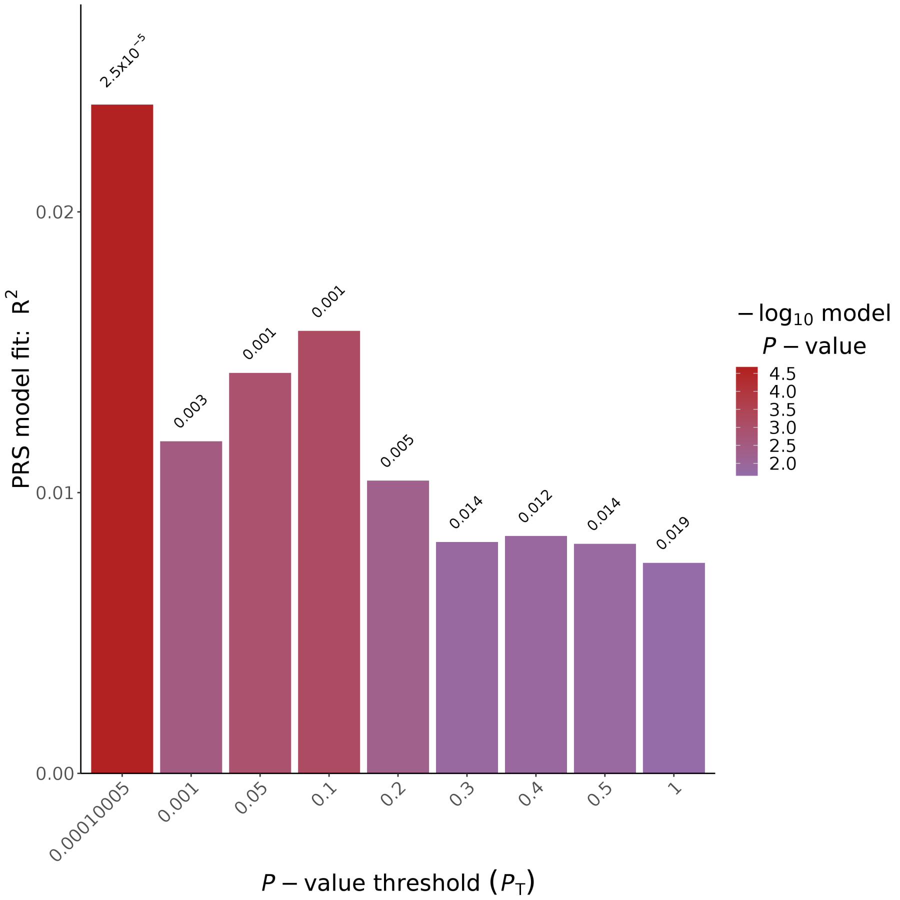


Figure 77: PGS for the MRI phenotype hippocampus volume right with SNPs from the Hibar GWAS

##### 4.2.5 Thickness

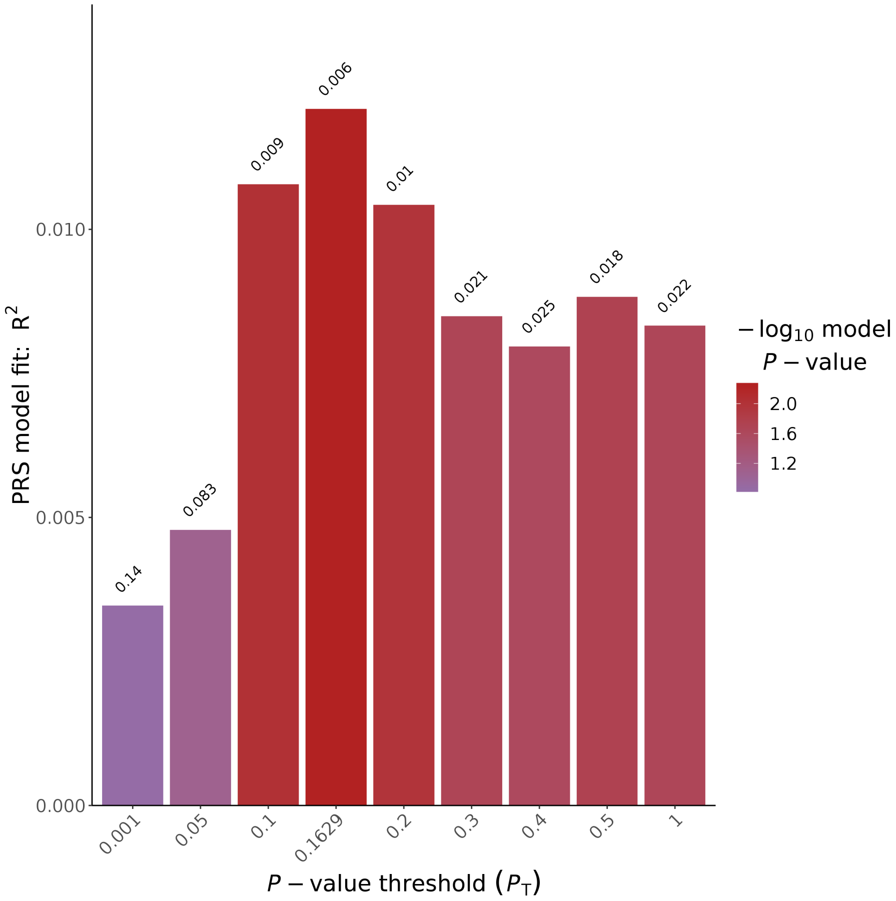


Figure 78: PGS for the MRI phenotype thickness with SNPs from the Hibar GWAS
